# A Large-Scale Genome-Wide Study of Gene-Sleep Duration Interactions for Blood Pressure in 811,405 Individuals from Diverse Populations

**DOI:** 10.1101/2024.03.07.24303870

**Authors:** Pavithra Nagarajan, Thomas W Winkler, Amy R Bentley, Clint L Miller, Aldi T Kraja, Karen Schwander, Songmi Lee, Wenyi Wang, Michael R Brown, John L Morrison, Ayush Giri, Jeffrey R O’Connell, Traci M Bartz, Lisa de las Fuentes, Valborg Gudmundsdottir, Xiuqing Guo, Sarah E Harris, Zhijie Huang, Mart Kals, Minjung Kho, Christophe Lefevre, Jian’an Luan, Leo-Pekka Lyytikäinen, Massimo Mangino, Yuri Milaneschi, Nicholette D Palmer, Varun Rao, Rainer Rauramaa, Botong Shen, Stefan Stadler, Quan Sun, Jingxian Tang, Sébastien Thériault, Adriaan van der Graaf, Peter J van der Most, Yujie Wang, Stefan Weiss, Kenneth E Westerman, Qian Yang, Tabara Yasuharu, Wei Zhao, Wanying Zhu, Drew Altschul, Md Abu Yusuf Ansari, Pramod Anugu, Anna D Argoty-Pantoja, Michael Arzt, Hugues Aschard, John R Attia, Lydia Bazzanno, Max A Breyer, Jennifer A Brody, Brian E Cade, Hung-hsin Chen, Yii-Der Ida Chen, Zekai Chen, Paul S de Vries, Latchezar M Dimitrov, Anh Do, Jiawen Du, Charles T Dupont, Todd L Edwards, Michele K Evans, Tariq Faquih, Stephan B Felix, Susan P Fisher-Hoch, James S Floyd, Mariaelisa Graff, Charles Gu, Dongfeng Gu, Kristen G Hairston, Anthony J Hanley, Iris M Heid, Sami Heikkinen, Heather M Highland, Michelle M Hood, Mika Kähönen, Carrie A Karvonen-Gutierrez, Takahisa Kawaguchi, Setoh Kazuya, Tanika N Kelly, Pirjo Komulainen, Daniel Levy, Henry J Lin, Peter Y Liu, Pedro Marques-Vidal, Joseph B McCormick, Hao Mei, James B Meigs, Cristina Menni, Kisung Nam, Ilja M Nolte, Natasha L Pacheco, Lauren E Petty, Hannah G Polikowsky, Michael A Province, Bruce M Psaty, Laura M Raffield, Olli T Raitakari, Stephen S Rich, Renata L Riha, Lorenz Risch, Martin Risch, Edward A Ruiz-Narvaez, Rodney J Scott, Colleen M Sitlani, Jennifer A Smith, Tamar Sofer, Maris Teder-Laving, Uwe Völker, Peter Vollenweider, Guanchao Wang, Ko Willems van Dijk, Otis D Wilson, Rui Xia, Jie Yao, Kristin L Young, Ruiyuan Zhang, Xiaofeng Zhu, Jennifer E Below, Carsten A Böger, David Conen, Simon R Cox, Marcus Dörr, Mary F Feitosa, Ervin R Fox, Nora Franceschini, Sina A Gharib, Vilmundur Gudnason, Sioban D Harlow, Jiang He, Elizabeth G Holliday, Zoltan Kutalik, Timo A Lakka, Deborah A Lawlor, Seunggeun Lee, Terho Lehtimäki, Changwei Li, Ching-Ti Liu, Reedik Mägi, Fumihiko Matsuda, Alanna C Morrison, Brenda WJH Penninx, Patricia A Peyser, Jerome I Rotter, Harold Snieder, Tim D Spector, Lynne E Wagenknecht, Nicholas J Wareham, Alan B Zonderman, Kari E North, Myriam Fornage, Million Veteran Program, Adriana M Hung, Alisa K Manning, James Gauderman, Han Chen, Patricia B Munroe, Dabeeru C Rao, Diana van Heemst, Susan Redline, Raymond Noordam, Heming Wang

**Author notes:** Corresponding Author: Heming Wang, PhD, Division of Sleep and Circadian Disorders, Brigham and Women’s Hospital, Harvard Medical School, 221 Longwood Ave BLI 252, Boston, MA 02115. These authors contributed equally (Pavithra Nagarajan, Thomas W Winkler, Amy R Bentley, Clint L Miller, Aldi T Kraja, Karen Schwander). These authors jointly directed this work (Patricia B Munroe, Dabeeru C Rao, Diana van Heemst, Susan Redline, Raymond Noordam, Heming Wang). Author Contributions: P.N. and H.W. conducted centralized project data analyses, data consolidation, meta-analyses, quality control, bioinformatics analysis, and contextual interpretation. P.N., H.W., A.R.B., A.T.K., C.L.M. K.S., T.W.W., P.B.M., R.N., D.C.R., S.R., and D.v.H. were part of the writing group and participated in project workflow design, interpretation of results, and drafting the manuscript. L.d.l.F., A.D., C.G., and D.C.R. participated in centralized study coordination. A.T.K., J.L.M., J.R.O., T.W.W., H.A., J.B.Meigs, X.Z., Han Chen, J.G., A.K.M., C.L.M., P.B.M., and P.A.P. acted as collaborators facilitating project design, specific code scripts, or specialized analyses. All other co-authors participated in final result interpretation, cohort-level study concept and design, cohort-level phenotype data acquisition and/or quality control, cohort-level genotype data acquisition and/or quality control, and/or cohort-level data-analysis and interpretation. All authors approved the final version of the paper that was submitted to the journal.

## Abstract

Although both short and long sleep duration are associated with elevated hypertension risk, our understanding of their interplay with biological pathways governing blood pressure remains limited. To address this, we carried out genome-wide cross-population gene-by-short-sleep and long-sleep duration interaction analyses for three blood pressure traits (systolic, diastolic, and pulse pressure) in 811,405 individuals from diverse population groups. We discover 22 novel gene-sleep duration interaction loci for blood pressure, mapped to genes involved in neurological, thyroidal, bone metabolism, and hematopoietic pathways. Non-overlap between short sleep (12) and long sleep (10) interactions underscores the plausibility of distinct influences of both sleep duration extremes in cardiovascular health. With several of our loci reflecting specificity towards population background or sex, our discovery sheds light on the importance of embracing granularity when addressing heterogeneity entangled in gene-environment interactions, and in therapeutic design approaches for blood pressure management.

## INTRODUCTION

Abnormal sleep duration is detrimental to cardiovascular health – increasing the risk of incident cardiovascular disease (CVD) and mortality – and inherently complex, with suspected heterogeneous effects according to sex and race/ethnicity^1,2^. Deviation from healthy sleep can impact diurnal rhythms, hormone levels (e.g. ghrelin, cortisol), autonomous nervous system balance, and even remodel vascular structure - resulting in adverse consequences, such as reduced nocturnal blood pressure (BP) dipping and sustained daytime hypertension^1,3^.

Yet the mechanistic pathways underlying the biomolecular connection between short and long sleep with cardiovascular health remain unclear. Evidence implicates heightened sympathetic tone and metabolic dysfunction in the mechanism of short sleep, but there remains a gap in clarity with the added complexity of interwoven pathways like oxidative stress and endothelial dysfunction^1,4^. The role of long sleep is more elusive, with recent work highlighting the pertinence of inflammatory markers, underlying comorbidity burden (i.e. dyslipidemia, depression) and arterial stiffness metrics^5,6^. This incomplete understanding of the intersection between habitual sleep duration and cardiovascular health necessitates further investigation.

Hypertension is a major risk factor for CVD, with blood pressure traits known to have a strong genetic background. Recent genome-wide association analyses (GWASs) have discovered more than two thousand loci explaining ∼40% of systolic or diastolic BP heritability among European descent individuals^7^. It is important to investigate the role of sleep health in such a polygenic landscape. This may both explain additional heritability of BP traits, as well as bring to the forefront novel genomic loci that inform perspective on sleep’s influence on biomolecular pathways underlying BP. Moreover, incorporating diverse population groups is essential – as this can reveal novel gene targets specific to particular subgroups or shared across – improving downstream therapeutic designs, and offering tangible insight to counter disparities in health.

Our prior work in the Cohorts for Heart and Aging Research in Genomic Epidemiology (CHARGE) Gene-Lifestyle Interactions Working Group highlighted novel non-overlapping gene-sleep interactions for BP, suggesting distinct roles of influence for short and long sleep duration^8^. Our current analysis advances the field by including a 12-fold larger sample size and additional sex-stratified analyses, yielding enhanced statistical power and granularity.

Here we report findings from genome-wide gene-by-sleep duration interaction analyses for BP traits, across 811,405 individuals of diverse population backgrounds (African [AFR], East Asian [EAS], European [EUR], Hispanic/Latino [HIS], and South Asian [SAS]). Cognizant of subliminal disparities that can shape sleep and cardiovascular health, and to ensure heterogeneity in interaction effects is addressed, we report findings stratified by both population group and sex parallel to our primary cross-population meta-analysis.

## RESULTS

### Overview

From an initial source of 37 studies, 59 population-group specific cohorts (derived from self-reported ancestry) resulted in a pooled sample size of 811,405 individuals comprising of 5.9% AFR (12 cohorts), 6.0% EAS (5 cohorts), 83.4% EUR (34 cohorts), 3.7% HIS (7 cohorts), and 0.9% SAS (1 cohort) [Supplementary Tables S1-S2]. Phenotypic and genetic data from each cohort were harmonized following a centralized protocol (see Methods). Population- and sex-stratified interaction models (M1) were analyzed for three BP traits (systolic BP [SBP], diastolic BP [DBP], pulse pressure [PP]) and two dichotomous sleep duration exposure (E) interaction variables (long total sleep time [LTST], short total sleep time [STST]). Similar to prior work, STST and LTST were defined by cohort-specific 20% and 80% quantiles of age- and sex-adjusted residuals of total sleep duration^8^. Marginal effect models for BP traits (M2) without interaction terms were also analyzed to screen for novel interactions. Covariate (C_1_ and C_2_) adjustment is described in the Methods. Genetic variants (G) were restricted to autosomal chromosomes with minor allele frequency:≥20.1%.

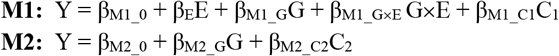

After quality-control of summary statistics, meta-analyses were performed both across population groups as cross-population meta-analysis (CPMA), and within specific population groups (AFR, EAS, EUR, HIS, SAS), stratified according to sex (combined sex, females only, males only) [Figure 1, Supplementary Figures S1-S2]. From these results, we identified evidence of gene-sleep duration interactions from the M1 model using the 1 degree of freedom (df) test of interaction effect (β_M1_G×E_), and the 2df joint test that simultaneously assesses the main effect (β_M1_G_) and the interaction effect (β_M1_G×E_)^9^. The marginal genetic effect in M2 (β_M2_G_) was utilized in a two-step protocol for detecting interactions.

**Figure 1.**
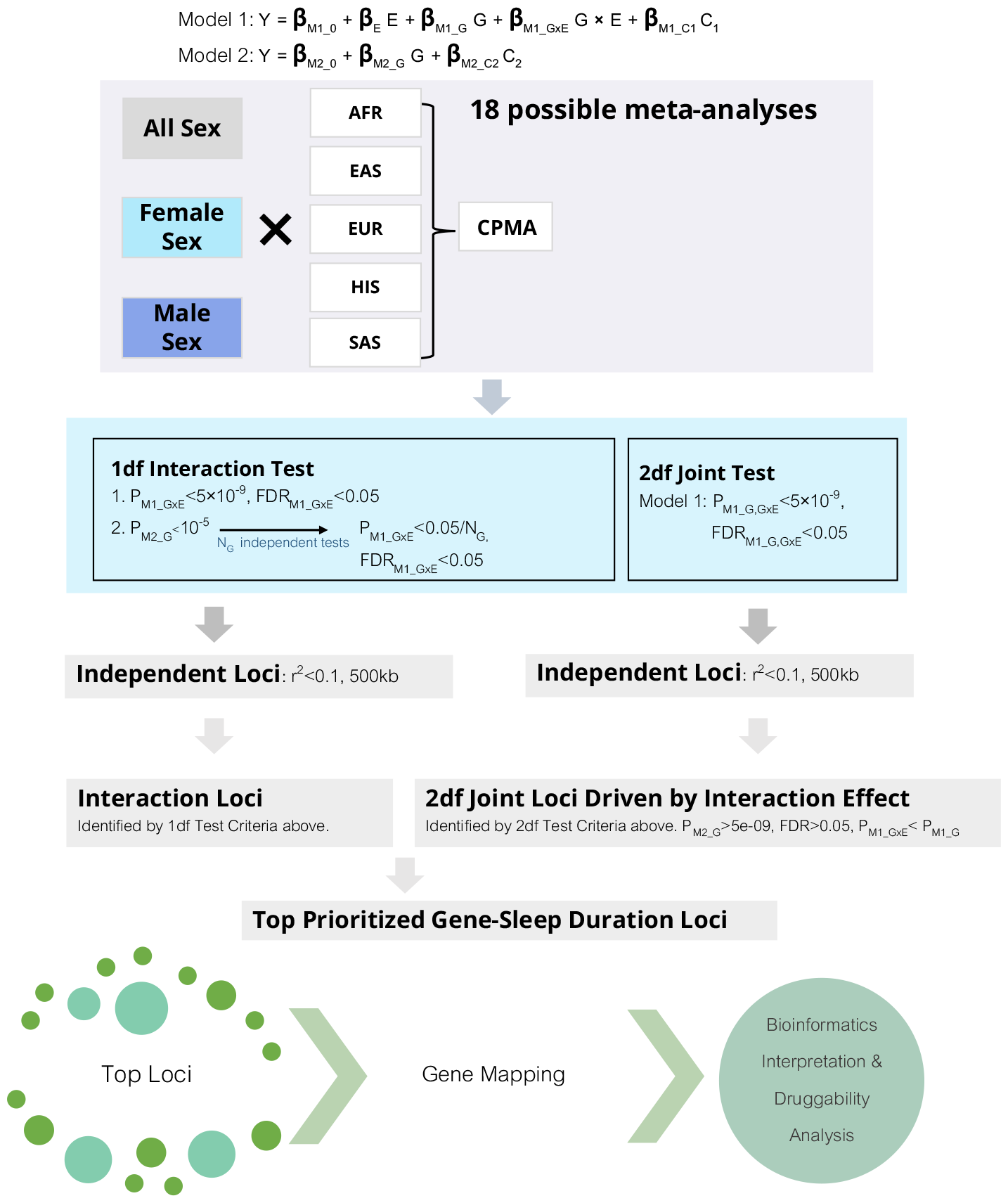
Analysis Workflow.

Variants were prioritized if: (i) significant in the 1df interaction test (*P*_M1_G×E_<5×10^−9^, FDR_M1_G×E_<0.05); (ii) significant in the two-step method where *z* independent variants prioritized based on marginal genetic effect (*P*_M2_G_<10^−5^) are evaluated using the 1df interaction test (*P*_M1_G×E<_0.05/*z*); or (iii) novel for BP, significant in the 2df joint test (*P*_M1_G,G×E_<5×10^−9^, FDR_M1_G,G×E_<0.05), insignificant in the marginal effect (*P*_M2_G_>5×10^−9^, FDR_M2_G_>0.05), and driven by the interaction effect (*P*_M1_G×E_< *P*_M1_G_) (Figure 1). Secondarily, variants novel for BP, significant in the 2df joint test, insignificant in the marginal effect, but not driven by the interaction effect (*P*_M1_G<_*P*_M1_G×E_) were noted. Novelty with respect to BP was defined as non-overlap with 1 Mb regions of prior reported BP GWAS associations (Supplementary Table S3). A full description of variant prioritization can be found in Methods.

This resulted in the 1df test revealing seven loci and the 2-step method revealing 1 locus. The 2df joint test first identified 3629 significant loci, from which 18 were novel for BP with insignificant marginal effect – revealing 14 loci driven by the interaction effect, and four not driven.

Thus in total we discovered 22 gene-sleep duration interaction loci, and 4 secondary loci – a total of 26 genetic variants of which 21 are novel for BP traits (Supplementary Table S4). Among the 22 prioritized interaction loci, four loci exhibited cross-population effects – one locus identified in combined sex and three in female sex-stratified analyses (Table 1); and 18 interaction loci were identified specific to either one of the AFR, HIS, or EUR population groups (Table 2).

**Table 1.**
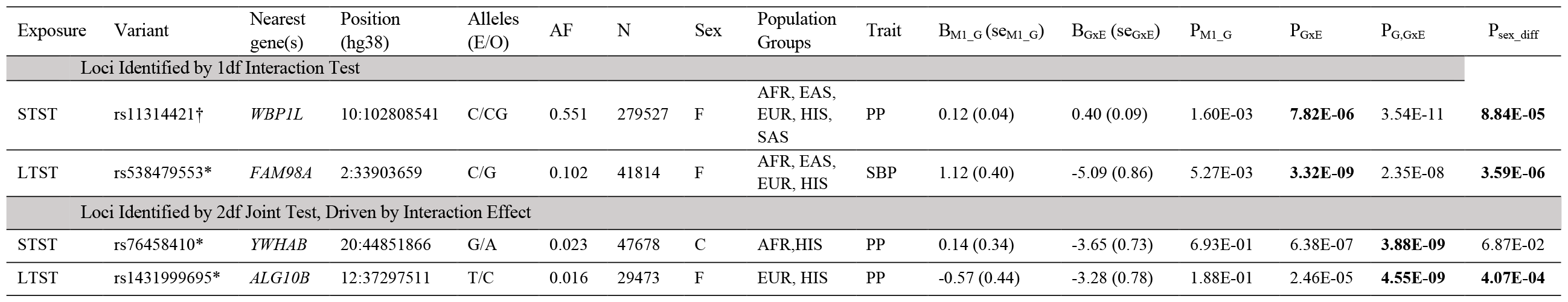
Novel Gene-Sleep Duration Interaction Loci Identified in Cross-Population Meta-Analysis All results herein are from the M1 model. Supplementary Table S21 provides summary statistics according to each population group identified in cross-population results. For the sex column, C denotes combined sex, and F denotes female sex-stratified meta-analysis. For the alleles column, E denotes effect allele, and O denotes other allele used as reference. Bold denotes significance. ^†^ denotes the variant was identified using the two-step approach. ^*^ denotes variants novel for BP.

**Table 2.** Novel Gene-Sleep Duration Interaction Loci Identified Specific to Certain Population Groups. All results herein are from the M1 model. Supplementary Table S22 provides summary statistics according to each population group-specific cohort identified in these population-group specific results. For the sex column, C denotes combined sex meta-analysis, and M denotes male sex-stratified meta-analysis. For the alleles column, E denotes effect allele, and O denotes other allele used as reference. Bold denotes significance. ^‪^ denotes variants novel for BP. ^†^ Empty cell in P_sex_diff_ indicates the variant, after quality control, was not found in both sex-stratified meta-analyses

| Exposure | Variant | Nearest gene(s) | Position (hg38) | Alleles (E/O) | Sex | AF | N | Population Groups | Trait | B<sub>M1_G</sub> (se<sub>M1_G</sub>) | B<sub>GxE</sub> (se<sub>GxE</sub>) | P<sub>M1_G</sub> | P<sub>GxE</sub> | P<sub>G,GxE</sub> | P<sub>sex_diff</sub><sup>†</sup> |
| --- | --- | --- | --- | --- | --- | --- | --- | --- | --- | --- | --- | --- | --- | --- | --- |
| Loci Identified by 1df Interaction Test |  |  |  |  |  |  |  |  |  |  |  |  |  |  |  |
| STST | rs114831731 | AHCYL1 | 1:109970740 | A/T | C | 0.009 | 23897 | HIS | PP | 0.85 (0.64) | -6.27 (1.04) | 1.87E-01 | **1.57E-09** | 7.34E-11 | **5.67E-04** |
|  | rs144229676 | ZNF521 | 18:25303461 | A/C | C | 0.019 | 27119 | HIS | DBP | 2.32 (0.70) | -7.23 (1.18) | 8.70E-04 | **8.10E-10** | 1.80E-09 | 5.00e-01 |
|  | rs1035064 | ZNF682 | 19:19997730 | T/C | M | 0.019 | 392939 | EUR | SBP | -0.27 (0.18) | 2.10 (0.36) | 1.62e-01 | **4.82E-09** | 5.76E-08 | **1.52E-04** |
|  | rs112958007* | PAK5 | 20:9805888 | C/T | C | 0.005 | 23897 | HIS | PP | 2.09 (0.97) | -10.83 (1.78) | 3.03E-02 | **1.28E-09** | 1.23E-08 |  |
|  | rs752086677* | KRTAP13-2 | 21:30362234 | C/G | C | 0.002 | 30009 | EUR | DBP | 2.12 (0.87) | -8.71 (1.23) | 1.47E-02 | **1.46E-12** | 5.68E-13 |  |
| LTST | rs372262693 | TG, SLA | 8:133069499 | T/C | C | 0.023 | 23897 | HIS | PP | 0.62 (0.41) | -4.91 (0.82) | 1.34E-01 | **2.33E-09** | 7.11E-09 | 2.43E-01 |
| Loci Identified by 2df Joint Test, Driven by Interaction Effect |  |  |  |  |  |  |  |  |  |  |  |  |  |  |  |
| STST | rs143863772* | MROH7 | 1:54707218 | T/G | C | 0.006 | 23902 | HIS | SBP | -1.56 (1.06) | -6.99 (1.74) | 1.47E-01 | 5.79E-05 | **1.84E-10** |  |
|  | rs141117715* | KCNJ3 | 2:155446948 | C/T | C | 0.021 | 23897 | HIS | PP | 0.51 (0.44) | -4.29 (0.76) | 2.53E-01 | 1.74E-08 | **2.45E-09** | 7.86E-02 |
|  | rs17011282* | ZNF385D | 3:22386254 | C/G | C | 0.006 | 23897 | HIS | PP | -0.01 (0.79) | -6.00 (1.20) | 9.93E-01 | 5.86E-07 | **4.13E-10** | 1.97E-01 |
|  | rs764985249* | EFNA5 | 5:105713137 | T/C | C | 0.002 | 26230 | EUR | PP | 0.97 (1.48) | -11.78 (2.00) | 5.09E-01 | 4.06E-09 | **6.45E-12** |  |
|  | rs542745170* | WWOX | 16:78323227 | A/C | C | 0.009 | 23897 | HIS | PP | -0.36 (0.72) | -4.96 (1.19) | 6.20E-01 | 3.14E-05 | **5.00E-09** | **2.05E-03** |
| LTST | rs533724062* | BRINP3 | 1:190792851 | TA/T | C | 0.011 | 34442 | AFR | PP | 1.13 (0.60) | -4.40 (1.18) | 6.03E-02 | 1.92E-04 | **3.69E-09** | **3.56E-04** |
|  | rs138288695* | CRBN | 3:3295965 | G/A | C | 0.006 | 23897 | HIS | DBP | 0.57 (0.68) | -5.50 (1.08) | 4.07E-01 | 3.46E-07 | **1.31E-09** |  |
|  | rs142966182* | ALCAM | 3:104791665 | T/C | C | 0.006 | 26230 | EUR | DBP | -0.20 (0.62) | -5.37 (1.01) | 7.50E-01 | 1.01E-07 | **1.21E-09** |  |
|  | rs540041583* | PAM | 5:103002447 | A/G | C | 0.003 | 26230 | EUR | PP | -1.54 (1.20) | -6.50 (1.66) | 1.99E-01 | 8.93E-05 | **6.93E-10** |  |
|  | rs113952142* | SDK1 | 7:3917121 | A/C | C | 0.005 | 23902 | HIS | SBP | -0.28 (1.20) | -9.07 (1.90) | 8.18E-01 | 1.81E-06 | **5.49E-10** |  |
|  | rs111392401* | JMJD1C | 10:65029197 | T/G | C | 0.007 | 23897 | HIS | DBP | 0.49 (0.63) | -5.71 (1.00) | 4.32E-01 | 1.27E-08 | **2.28E-10** |  |
|  | rs772862932* | ATP8A2 | 13:25395875 | T/C | C | 0.002 | 30009 | EUR | DBP | -0.11 (0.70) | -4.85 (1.00) | 8.70E-01 | 1.31E-06 | **1.28E-09** |  |

**Table 3.**
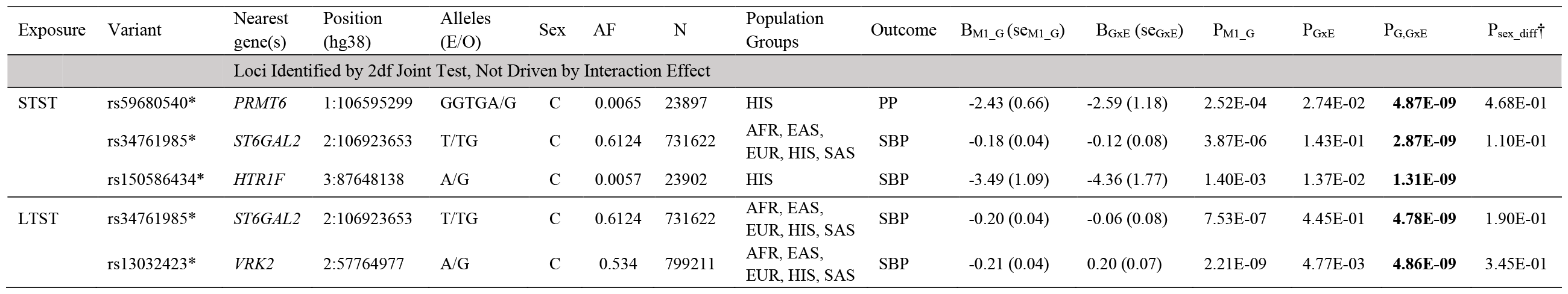
Novel BP Loci Identified by the 2df Joint Test, Not Driven by the Interaction Effect All results herein are from the M1 model. Supplementary Tables S21-S22 provides summary statistics according to each population group-specific cohort for rs59680540 and rs150586434, and according to each population group for rs34761985 and rs13032423 below. For the sex column, C denotes combined sex meta-analysis. For the alleles column, E denotes effect allele, and O denotes other allele used as reference. Bold denotes significance. ^‪^ denotes variants novel for BP. ^†^ Empty cell in P_sex_diff_ indicates the variant, after quality control, was not found in both sex-stratified meta-analyses

### Cross-Population Gene-Sleep Duration Interactions

A total of four cross-population gene-sleep duration interaction loci were identified (Table 1). In combined sex, the 2df joint test revealed a locus driven by STST interaction for PP at rs76458410 (*YWHAB*; *P*_GxE_=6.4×10^−7^; *P*_G,GxE_ =3.9×10^−9^) (Supplementary Figure S3). In female sex-specific analysis, the 2-step protocol revealed rs11314421 (*WBP1L*; *P*_GxE_=7.8×10^−6^; *P*_G,GxE_= 3.5×10^−11^) interacting with STST for PP, the 1df test identified rs538479553 (*FAM98A*; *P*_GxE_=3.3×10^−9^) interacting with LTST for SBP, and the 2df test revealed rs1431999695 (*ALG10B*; *P*_GxE_=2.5×10^−5^; *P*_G,GxE_ =4.5×10^−9^) driven by LTST interaction for PP (Figure 2, Supplementary Figure S3).

**Figure 2.**
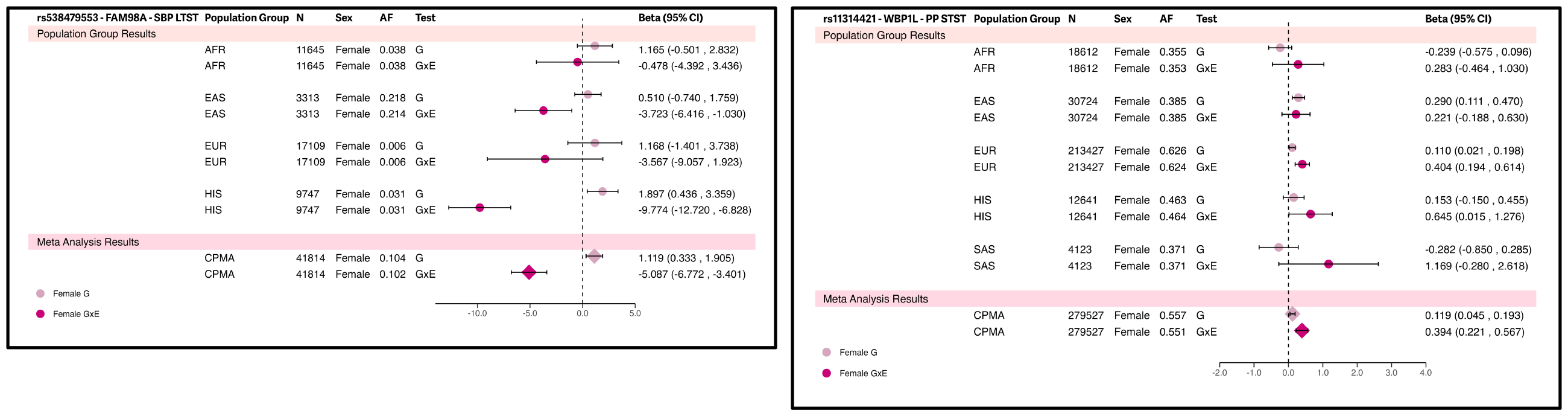
Forest Plots of Gene-Sleep Duration Loci Identified by the 1df Interaction Test in Female-Specific Cross-Population Meta-Analyses. These are the prioritized 1df Interaction Test results from female-specific cross-population meta-analyses. Contributing female-specific population groups’ (if this variant is found in the particular population group, after quality control) summary statistics are depicted here that were pooled.

### AFR-Specific Gene-Sleep Duration Interactions

Specific to AFR, the 2df joint test identified a locus driven by LTST interaction at rs533724062 (*BRINP3*; *P*_GxE_=1.9×10^−4^; *P*_G,GxE_ =3.7×10^−9^) for PP (Table 2, Supplementary Figure S3).

### HIS-Specific Gene-Sleep Duration Interactions

Specific to HIS, a total of 11 gene-sleep duration interaction loci were identified (Table 2). The 1df interaction test identified four interaction loci: three loci exhibiting interaction with STST at rs114831731 (*AHCYL1*; *P*_GxE_=1.6×10^−9^) for PP, rs144229676 (*ZNF521*; *P*_G_×_E_=8.1×10^−10^) for DBP, and rs112958007 (*PAK5*; *P*_GxE_=1.3×10^−9^) for PP; and one locus exhibiting interaction with LTST at rs372262693 (*TG,SLA*; *P*_GxE_=2.3×10^−9^) for PP (Figure 3). The 2df joint test identified four loci driven by STST interactions at rs143863772 (*MROH7*; *P*_GxE_=5.8×10^−5^; *P*_G,GxE_=1.8×10^−10^) for SBP, rs141117715 (*KCNJ3*; *P*_GxE_=1.7×10^−8^; *P*_G,GxE_ =2.4×10^−9^) for PP, rs17011282 (*ZNF385D*; *P*_GxE_=5.9×10^−7^; *P*_G,GxE_ =4.1×10^−10^) for PP, and rs542745170 (*WWOX*; *P*_GxE_=3.1×10^−5^; *P*_G,GxE_ =5.0×10^−9^) for PP; and three loci driven by LTST interactions at rs138288695 (*CRBN*; *P*_GxE_=3.5×10^−7^; *P*_G,GxE_ =1.3×10^−9^) for DBP, rs113952142 (*SDK1*; *P*_GxE_=1.8×10^−6^; *P*_G,GxE_ =5.5×10^−10^) for SBP, and rs111392401 (*JMJD1C*; *P*_GxE_=1.3×10^−8^; *P*_G,GxE_ =2.3×10^−10^) for DBP (Supplementary Figure S3).

**Figure 3.**
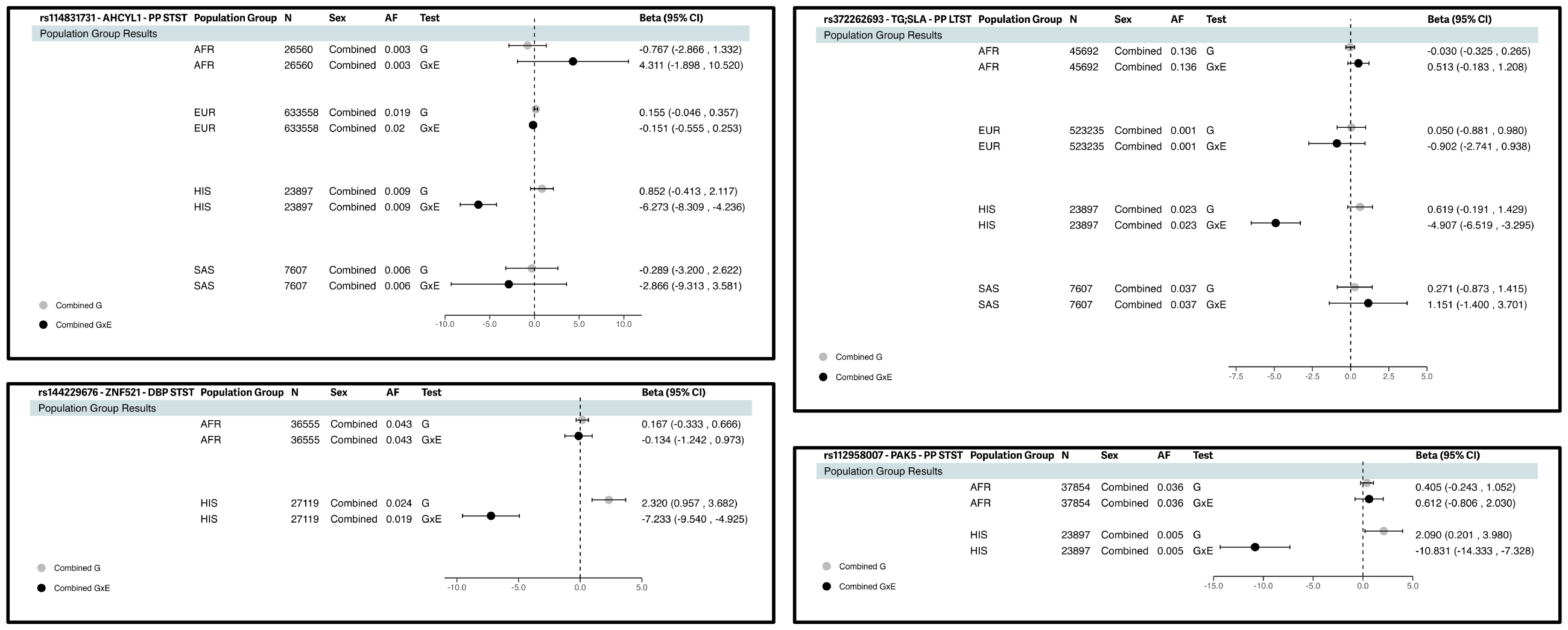
Forest Plots of Gene-Sleep Duration Loci Identified by the 1df Interaction Test in Combined Sex HIS-Specific Meta-Analyses These are the prioritized 1df Interaction Test results from HIS-specific meta-analyses. Other population group data (if this variant is found in the particular population group, after quality control) are shown here to emphasize that these gene-sleep duration interaction loci were significant only in the HIS population group.

### EUR-Specific Gene-Sleep Duration Interactions

Specific to EUR, six gene-sleep duration loci were identified – five loci in combined sex and one locus in male sex-stratified analysis (Table 2). The 1df interaction test identified rs752086677 (*KRTAP13-2*; *P*_GxE_=1.5×10^−12^) showing interaction with STST for DBP (Supplementary Figure S3). The 2df joint test identified one locus driven by STST interaction for PP at rs764985249 (*EFNA5*; *P*_GxE_=4.1×10^−9^; *P*_G,GxE_= 6.4×10^−12^); and three loci driven by LTST interactions at rs142966182 (*ALCAM*; *P*_GxE_=1.0×10^−7^; *P*_G,GxE_ = 1.2×10^−9^) for DBP, rs540041583 (*PAM*; *P*_GxE_=8.9×10^−5^; *P*_G,GxE_ =6.9×10^−10^) for PP, and rs772862932 (*ATP8A2*; *P*_GxE_=1.3×10^−6^; *P*_G,GxE_ = 1.3×10^−9^) for DBP (Supplementary Figure S3). The 1df interaction test in male-specific EUR meta-analysis uniquely revealed rs1035064 (*ZNF682*; *P*_GxE_=4.8×10^−9^) harboring interaction with STST for its effect on SBP (Table 2, Supplementary Figure S3).

### Sex Differences in Interactions

Testing for sex differences (*P*_sex_diff_<0.002) revealed variants identified in combined sex meta-analyses – rs114831731 (*AHCYL1*), rs542745170 (*WWOX*), and rs533724062 (*BRINP3*) – showing evidence of heterogeneous effect by sex (Table 2).

### Prior Reported Gene-Sleep Interactions

Our previous work identified two STST interaction loci (rs73493041 for DBP, rs10406644 for PP) and one LTST interaction locus (rs7955964 for mean arterial pressure) utilizing a smaller subset of cross-population samples (N=62,969)^8^. In our current cross-population meta-analysis, rs10406644 showed interaction evidence with STST (*P*_G×E_=2.0×10^−4^ for PP), and rs7955964 with LTST (*P*_G×E_=6.6×10^−3^ for SBP, *P*_G×E_=1.7×10^−2^ for DBP) with direction of association agreeing with prior work, and non-significance (*P*_GxE_>0.05) for the opposite sleep duration exposure (Supplementary Table S5).

### Functional Potential of Variants

Variants were assessed using FAVOR and RegulomeDB to annotate deleteriousness or functionality scores^10,11^ (Supplementary Table S6). Four variants (rs11483173, rs372262693, rs113952142, rs11314421) were marked by high transcription activity chromatin states and eight variants (rs114831731, rs372262693, rs143863772, rs533724062, rs11314421, rs1035064, rs538479553, rs13032423) were marked by accessible chromatin in heart tissue or blood. Six variants (rs114831731, rs542745170, rs533724062, rs11314421, rs1035064, rs538479553) reflected marked regulatory potential with RegulomeDB scores ≥22c.

### Mapped Protein-Coding Genes

All 26 variants were either intronic or intergenic, and mapped to a primary set of 27 protein coding genes as identified by: (1) direct overlap with genes (i.e. intronic), (2) shortest distance to transcription start site, or (3) shortest distance to gene start/end site (Supplementary Table S7). Cognizant of the complexity of genomic variation, a secondary extended list of genes was annotated for each locus by positional overlap, chromatin interaction (CI), or eQTL evidence accounting for linkage disequilibrium (LD) structure using FUMA SNP2GENE and RegulomeDB^10,12^ (Methods, Supplementary Tables S7-S9). This extended mapping revealed 292 genes highlighted for the 12 STST interaction loci, 67 genes for the 10 LTST interaction loci, and 35 genes for the four joint 2df loci not driven by interaction.

### Expression Quantitative Trait Loci (eQTL)

Tissue-specific (GTEXv8) eQTL associations were observed at rs11314421, rs1035064, and rs34761985 in tissues of the heart, vasculature, or blood (Supplementary Table S8)^10^. *WBP1L*-rs11314421 and *ZNF682*-rs1035064 gene mappings were corroborated by eQTL evidence identified in venous blood or the tibial artery. Beyond these primary mapped genes, *MFSD13A, BORCS7, CALHM2, RPARP-AS1, AS3MT*, and *SFXN2* expression were mapped to rs11314421 by eQTL evidence in the ascending aorta, coronary artery, tibial artery, left ventricle myocardium, right atrium auricular region, venous blood, or lymphoblast. Similarly, *ZNF56, ZNF253, ZNF93, ZNF90*, and *ZNF486* were mapped by coronary artery or venous blood eQTL evidence to rs1035064. *UXS1* was mapped by rs34761985 eQTL data identified in the right atrium auricular region.

### Variant-Level Cross-Trait Associations

Several variants show associations (*P*<5×10^−8^) with other traits (Supplementary Table S10). Querying Open Target Genetics (https://genetics.opentargets.org) identified rs11314421 (*WBP1L)* to be associated with hypertension and testosterone levels, and rs13032423 *(VRK2)* with sleep duration and feeling miserable (Supplementary Table S11). Common Metabolic Diseases Knowledge Portal (https://hugeamp.org/) further identified rs13032423 *(VRK2)* to be associated with BMI and sleep duration (Supplementary Figure S4). Querying brain imaging phenotypes through the Oxford Brain Imaging Genetics Server (BIG40) revealed rs13032423’s (*VRK2)* connection to brain functional connectivity by its association with rfMRI connectivity (ICA100 edge 965) (Supplementary Table S12, Supplementary Figure S5)^13^.

### Gene Functional Implications

Reported evidence from gene knockout studies and genetic association analyses were synthesized to holistically understand functionality of the primary mapped genes (Supplementary Table S10). Mice knockout evidence from the International Mouse Phenotyping Consortium highlighted genes important for heart morphology (*BRINP3, CRBN, ALG10B, PRMT6*), and cardiac rhythm (*TG, WBP1L*).^14^ (Supplementary Table S13). Open Target Genetics, PheWeb, and PheGenI revealed genes implicated in genetic associations (p<5×10^−8^), with OMIM (https://omim.org/) identifying any linked Mendelian disorders (Supplementary Tables S14-S17)^15-17^. Specifically, eight genes harbored links to the cardiovascular domain through association with traits identifying by genetic studies: *WBP1L, EFNA5, ZNF521, WWOX, ZNF385D, FAM98A, PAM*, and *JMJD1C. ALG10B* was identified to be implicated in the Mendelian disorder long QT syndrome. In the realm of sleep and circadian health, reported genetic associations corroborated the relevance of *EFNA5, ZNF52*1, *WWOX, ALG10B, PAM* and *SDK1* with insomnia, daytime napping, or chronotype traits. Genetic associations to other pertinent domains including kidney function, neurological health, liver function, thyroid function, metabolism, lifestyle choice, and inflammation, were also synthesized (Supplementary Table S10).

### Gene Set Enrichment Analysis

To formally investigate distinction between STST and LTST pathways, we performed gene set enrichment analyses on the aforementioned extended gene sets in the FUMA GENE2FUNC platform and STRING database (Supplementary Tables S18-S19)^12,18^. STST-mapped genes highlighted pathways in antioxidant defense and neuron excitation, along with phenotypic connection to lipid levels, neurological health, cardiovascular health, metabolism and immune defense. LTST-mapped genes implicated traits involving inflammation, neurological health, and metabolism. A clearly distinctive pattern differentiating short and long sleep duration interaction loci was thus not observed.

### Druggability

We investigated druggability of the primary mapped genes using an integrative approach to highlight drug repurposing potential (Supplementary Table S20)^19^. First, genes were queried in the Drug-Gene Interaction database revealing those marked as clinically actionable or members of the druggable genome^20^. These identified gene candidates revealed connections to serotonergic response (*HTR1F, KCNJ3*), proteasome-mediated ubiquitination (*CRBN*), thyroid hormone synthesis (*TG*), and axon guidance (*PAK5, ALCAM*) pathways. Of these, *KCNJ3, CRBN, HTR1F, TG, PAK5*, and *ALCAM* harbored links to reported drug interactions and active ligand interactions in the ChEMBL database. Drug-gene interactions with FDA-approved drugs were queried for involvement in late-stage clinical trials using DrugBank and ClinicalTrials.gov (https://clinicaltrials.gov/)^21,22^. This identified the following genes displaying evidence of pharmacological targeting: *KCNJ3* (by small molecule inhibitors Atomoxetine and Dronedarone); *CRBN (*by thalidomide analogs Pomalidomide and Lenalidomide); *HTR1F* (by selective serotonin receptor agonists like Lasmiditan); and *ALCAM* (by chemotherapy agent Fluorouracil).

## DISCUSSION

In this large-scale effort investigating the biomolecular mechanisms underpinning the intersecting roles of sleep health and blood pressure traits, we conducted genome-wide gene-by-sleep duration (short and long sleep) interaction analyses in 811,405 individuals of diverse population backgrounds (AFR, EAS, EUR, HIS, SAS) for systolic blood pressure, diastolic blood pressure, and pulse pressure. We report novel discovery of 22 gene-sleep duration interaction loci for BP traits – 12 for short sleep, and 10 for long sleep. Several of the identified variants are rare with allele frequency <=1%, with four variants identified in sex-stratified meta-analyses, and 18 variants specific to either the AFR(1), EUR(6), or HIS (11) population groups. In line with our previous research, the identified genomic loci exhibiting interactions with short and long sleep are non-overlapping (with non-significance in the opposing sleep duration exposure), suggesting distinct mechanisms influencing cardiovascular health. Nonetheless, we did not observe a clear differentiating pattern in the biological pathways implicated when comparing short sleep and long sleep.

The functional annotation investigations of our prioritized genes point towards cardiovascular and neurological connections, along with revealing links to circadian rhythm, thyroid function, bone health, and hematopoiesis mechanisms. Our findings highlight potential pharmacological candidates and suggest pertinent pathways to consider when designing holistic therapeutic regimens for improving blood pressure control.

Firstly, at a broad level, several identified genes are tied to neurological mechanisms. *KCNJ3* encodes Kir3.1 – the alpha subunit for the I_KACh_ potassium channel – and is interestingly implicated in bradyarrhythmia by its missense variant inducing a gain of function of I_KACh_, as activation of this channel is tied to the negative chronotropic effect on heart rate exerted by the parasympathetic nervous system^23^. *CRBN* is linked to cognitive function ^24^, *SDK1* promotes synaptic connectivity ^25^, *ZNF521* regulates neuron cell fate ^26^, and *ATP8A2* is involved in both neuron vesicle transport and cardiac conduction ^27^. Further, *KRTAP13-2, WWOX, EFNA5*, and *ALCAM* are linked to nervous system development with additional roles for *WWOX* in myelination ^28^ and *EFNA5* in vascular sympathetic innervation ^29^. These functional connections may suggest a potential nervous system-heart connection that could be influenced by sleep or circadian disturbances.

In fact neurological pathway connections to circadian rhythm reveal themselves through two enzymes - *PAK5* and *PAM*. Given that circadian rhythm and clock gene expression is intimately connected to blood pressure patterns, of note is *PAK5* – a serine/threonine kinase protective of adult neurons from injury and ischemic stress^30^. *PAK5* has both been shown to be targeted by clock gene-regulated miRNAs in the liver and identified to strongly bind to 14-3-3 proteins – a protein family connected to light-sensitive melatonin diurnal patterns and plausibly influential for sleep behavior^31,32^. This strong binding affinity to 14-3-3 proteins suggests an interesting connection, as *YWHAB* (one of this study’s primary genes mapped to a STST interaction locus), is part of this protein family. Another enzyme informing the neurological-sleep axis is *PAM*, encoding a copper-dependent enzyme important for synthesizing amidated neuropeptides like NPY – which regulates sleep through noradrenergic signals^33,34^.

Further, *TG* and *JMJD1C*, both encoding proteins intrinsically tied to thyroid hormone function (thyroglobulin and thyroid receptor-interacting protein 8 respectively) – present suggestive ties to the intersection between thyroid function and circadian rhythms. *TG* mRNA and protein expression levels have shown to increase in response to melatonin, along with its genetic variants associated with autoimmune thyroid diseases^35,36^. Gene silencing of *JMJD1C*’s paralog has shown arrhythmicity and prolonged sleep in Drosophila^37^. Given that circadian clock and thyroid function are increasingly suggested to be interconnected, and sleep deprivation can disrupt temporal hormone profiles (e.g. increased morning plasma thyroid-stimulating hormone (TSH) levels), it may be valuable to investigate further the overlapping pathways between thyroid function, healthy sleep duration, and cardiovascular morbidity^38^.

Beyond thyroidal pathways, hematopoiesis presents a possible comprehensive perspective on the interconnectedness between sleep health and nervous system response. *WBP1L*, one of the primary genes identified (mapped to a STST interaction locus identified in female-specific CPMA) has suggestive connection to regulating the *CXCL12*-*CXCR4* signaling pathway by its inhibitory role on *CXCR4*, the receptor for ligand *CXCL12*^39^. This pathway is both influential for inflammation and hematopoietic state, reflects circadian control, and directly implicates the sympathetic nervous system response – pertinent as stressors are suspected to induce a more exacerbated response in females^40,41^. If stress factors (e.g. sleep loss) induce noradrenaline, this can downregulate *CXCL12*, with resultant increased cell proliferation of pro-inflammatory cells from the bone marrow, incurring vascular damage^40^. For instance, fragmented sleep has shown to promote myelopoiesis and lower hypocretin release by the hypothalamus, in turn accelerating atherosclerosis progression^42^. Thus perhaps WBP1L can offer insight into the intersections between sympathetic activation, neurological control, and unhealthy sleep impacting cardiovascular health, especially in women.

On a similar note of addressing sex-specificity, of relevance is *FAM98A*, a gene identified in female-specific CPMA for interaction with long sleep. *FAM98A*, harboring multiple arginine demethylation sites, is a substrate of *PRMT1* - an enzyme which catalyzes the synthesis of asymmetric dimethylarginine (ADMA), a molecule associated with cardiovascular harm as it induces endothelial dysfunction^43^. Thus seeking to lower harmful ADMA levels to counter harmful effects of sleep loss may be relevant in preservation of vascular integrity^44^. *FAM98A*, encoding a microtubule-associated protein, is also functionally linked to osteoclast formation, which is key to bone resorption and involved in postmenopausal osteoporosis etiology^45^. Given that osteoporosis and CVD share pathology, the *FAM98A* locus may shed light on the importance of considering holistic treatment for hypertensive women approaching or after menopause – an example being Felodipine, an antihypertensive found to additionally discourage osteoclast differentiation^46,47^.

Beyond *FAM98A*, specific genes highlight pathway connections to offer possible avenues for enhancing treatment efficacy for hypertension. Addressing the role of inflammation, *SLA* may lend promise as an immunosuppressant, with cytoplasm-specific delivery of specific domains of *SLA* shown to inhibit the T cell receptor functional cascade^48^. *ALG10B* closely interacts with *KCNH2* to protect it from inhibition by pharmaceuticals and thus prevent acquired long QT syndrome - interesting, as past work has identified *KCNH2* genetic variation to associate with efficacy of specific antihypertensive drugs^49,50^. *PAK5* is the effector protein of *CDC42*, vital for endothelial integrity and involved in the mechanism of Nebivolol, a third generation beta-blocker^51,52^. *CRBN*, due to its intrinsic role in ubiquitination, is recruited as an E3 ligase ligand in protease-targeted chimeras (PROTACs), which hold promise in cardiovascular therapeutics – an example being P22A shown to reduce collateral damage of HMGCR upregulation caused by statins^53^. These findings point to the need for future preclinical and clinical studies to confirm the hypothesized mechanisms and test promising interventions.

Our druggability analysis specified genes acting as existing pharmacological targets of FDA-approved drugs, offering perspective for drug repurposing. *HTR1F* and *KCNJ3* are linked to the serotonergic pathway and are targets of approved ADHD and antiarrhythmic drugs Atomoxetine and Dronedarone, respectively. This is potentially relevant given that serotonin may impact blood pressure regulation, and serotonin receptor desensitization is implicated in chronic sleep restriction^54,55^. *HTR1F* encodes for 5-HT_1F_, shown to function in smooth muscle and trigeminal nerves, with its selective agonists (i.e. Lasmiditan) offering greater efficacy for migraine treatment without the collateral harm of vasoconstrictive effects induced by non-selective triptans^56^.

Noticeably, all 22 gene-sleep duration interaction loci we identified were specific to a particular population group, a subset of population groups, or a particular sex. This may be due to substantial heterogeneity in BP architecture and sleep lifestyle as a result of cultural differences, uniquely varying stressors due to socioeconomics, and genetic risk that are both shaped by and influence lifestyle choices. For example, admixed African and Hispanic populations are more likely to have poorly controlled hypertension and circadian abnormalities in BP regulation, as well as higher prevalence of both short and long sleep duration relative to individuals of European ancestry^57,58^. Females generally sleep longer, have higher prevalence of insomnia, and experience an increased proinflammatory response to sleep deprivation compared to males^59^. Such differential risk profiles are likely attributed to a myriad of social or environmental variables along with genetic and epigenetic susceptibility^8^. Therefore, it is likely that the same duration of self-reported sleep has different etiologies and physiological effects across sex and population background. Future research incorporating extensive phenotyping may help clarify whether gender-specific or population-specific findings are explained by differences in sleep-related or other lifestyle behaviors, mechanisms underlying response to sleep disturbance, or are spurious.

This study has several strengths, including its large-scale nature made possible by inclusion of several international biobanks and cohort studies, rigorous data harmonization and quality control protocols, and robust statistical analysis pipelines. Our findings are reinforced by multiple lines of evidence from bioinformatics analysis. Focused druggability analysis and interpretation of drug-gene interactions offer promising insight in drug repurposing and candidate targets for future pursuits.

Limitations of this study include the risk of unidentified misclassification of self-reported sleep duration (opposed to objective measurements from actigraphy or polysomnography) due to recall bias, sleep misperception, or other psychosocial factors. Sleep health is complex, with key dimensions beyond duration (e.g., timing, quality, satisfaction, and regularity)^60^. Abnormality in these other sleep dimensions were not tested here due to lack of readily available data. Adding to the complexity, sleep duration itself reflects heterogeneous health effects influenced by genetic determinants. For example, genetic variation conducive to naturally short sleepers may even lend neuroprotection against harmful brain pathology^61^. In addition, there may be residual confounding bias due to unadjusted comorbidities or environmental factors. Lastly, despite notable diversity of our sample, our data was dominated by individuals of European ancestry. It is striking that several of our loci are HIS-specific – which may be resultant of complex admixture present in this population group. Although we were able to delve into sex-specific interpretations for FAM98A, and WBP1L – future investigation is desired to understand the reasons behind heterogeneous effects by sex. Enrichment of sample sizes in minority populations is critical for future investigations.

In conclusion, this study advances our understanding of the interaction between sleep duration extremes and genetic risk factors shaping the genetic landscape of blood pressure. Our novel discovery of 22 gene-sleep duration interaction loci both accentuates the relevance of proper sleep duration in cardiovascular health and the need to be conscious of heterogeneity present in specific sex or population groups, providing valuable perspective for therapeutic intervention strategies to address cardiovascular disease burden.

## METHODS

This work was approved by the Institutional Review Board of Washington University in St. Louis and complies with all relevant ethical regulations. For each of the participating cohorts, the appropriate ethics review board approved the data collection and all participants provided informed consent.

### Data Harmonization

Data from each cohort were harmonized following this centralized protocol. Data were stratified by population group, based on self-reported ancestry and individual cohort definitions (AFR: African, EAS: East Asian, EUR: European, HIS: Hispanic/Latinos, SAS: South Asian), and sex (combined sex, female sex, male sex). Analyses considered 3 primary blood pressure (BP) traits as outcome variables (SBP: systolic, DBP: diastolic, PP: pulse pressure) and 2 dichotomous lifestyle exposures (LTST: long total sleep time, STST: short total sleep time). Genetic variants (G) were restricted to autosomal chromosomes 1-22 imputation quality≥20.3, and minor allele frequency≥20.1%. Age was restricted to ≥218 years, and reported total sleep time constrained within 3 and 14 hours. In scenarios of multiple visits, the single visit with largest sample size was utilized and in case-control study designs, cases and controls were required to be analyzed separately. For BP outcome measures, if multiple readings were taken in a single visit the mean was used. All BP values were winsorized at 6 standard deviations from the mean. BP values were adjusted for reported use of anti-hypertensive medications as follows: SBP (+15 mmHg) and DBP (+10 mmHg). PP was derived as SBP – DBP. In the case of studies with known between-sample relatedness, null model residuals (regressing BP traits on a kinship matrix/genetic covariance matrix) were denoted as the BP outcome. STST and LTST were derived from total sleep time (TST) by regressing TST on age, sex, age×sex and using the residuals’ 20^th^ and 80th percentiles as cutoffs (STST=1 if ≤ 20^th^ percentile, LTST=1 if ≥ 80^th^ percentile, STST=0 if > 20^th^ percentile, LTST=0 if < 80^th^ percentile). Covariates included population-group specific principal components, cohort-specific confounders (study center), age, age^2^, sex, age×S/LTST, age^2^×S/LTST, and sex×S/LTST. Samples with missing data were excluded.

### Data Analysis

After data harmonization, each population-group specific cohort ran 2 regression models (M1 and M2) for 18 phenotype-exposure-sex combinations (3 phenotypes x 2 exposures x 3 sex groups: combined sex, female sex, male sex). Below E denotes the lifestyle exposure (STST or LTST), Y denotes the BP outcome (SBP, DBP, or PP), C_1_ denotes the vector of covariates incorporating E (age, age^2^, S/LTST, age*S/LTST, age^2^*S/LTST, sex, sex*S/LTST), and C_2_ denotes the subset without incorporating E (age, age^2^, sex). Female-specific and male-specific analyses were not adjusted for sex. Specialized software choice included LinGxEScanR v1.0 (https://github.com/USCbiostats/LinGxEScanR), GEM v1.4.1 (https://github.com/large-scale-gxe-methods/GEM), and/or MMAP (latest version available) (https://github.com/MMAP/MMAP.github.io) with robust standard errors (SEs) enforced^62^. One degree of freedom (df) tests for the marginal effect (β_M2_G_), the main effect (β_M1_G_), and the interaction effect (β_M1_GxE_) were conducted; alongside the 2df joint test that simultaneously assesses the main effect and the interaction effect (β_M1_G_, β_M1_GxE_)^9^.

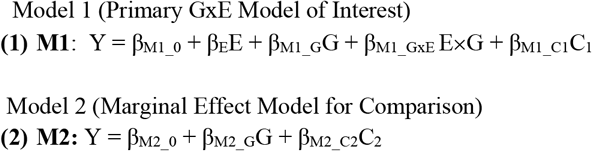

### Quality Control (Cohort-Level, Meta-Level)

Summary statistics were centrally processed after individual studies submitted results. EasyQC2 software (www.genepi-regensburg.de/easyqc2) was used to perform quality control (QC) on resultant data^63^. Data were filtered for degrees of freedom≥220 calculated as minor allele count * imputation quality (e.g. MACxR^2^ provided by each cohort) within the unexposed, the exposed, and the total sample. Missing or invalid/out of range values for statistics and duplicated or monomorphic variants were discarded. hg19 genomic coordinates were lifted over to hg38 genomic coordinates. Allele frequency discrepancies relative to TOPMed-imputed 1000G reference panels (Trans-Omics for Precision Medicine imputed 1000Genomes) were assessed for each specific population group, along with genomic control (GC) lambda inflation. Next, meta-level quality control was conducted within groups based on population group (AFR: 12 cohorts, EAS: 5 cohorts, EUR: 34 cohorts, HIS: 7 cohorts, SAS: 1 cohort), with evaluation of unwanted centering of the outcome variable, outlying cohorts highlighting unstable numerical computation, or alarming inflation.

### Meta-Analysis

Meta-analysis was designed as the following paradigm. Cross-population meta-analysis (CPMA) was designed to be combine all population group results, with additional focused population-group specific and sex-specific analyses. This resulted in 18 total meta-analyses to be run: 6 population groups (CPMA, EUR, HIS, EAS, AFR, SAS) and 3 sex groups (combined sex, female sex, male sex). To accomplish this, METAL software was first used to run all meta-analyses within each specific population group for the marginal effect (β_M2_G_), main effect (β_M1_G_), interaction effect (β_M1_GxE_), and joint effect (β_M1_G_, β_M1_GxE_) with GC correction for inflation^64^. Inverse-variance weights were used and Manning et. al’s method for the 2df joint test ^65^. CPMA was subsequently executed on the resultant population-group specific METAL output results with GC correction.

### Genome-wide Significant Loci Identification

EasyStrata2 software was used to prioritize top loci from significant results identified from the 1df interaction and 2df joint tests^66^. GC correction for population-group specific results was applied. Variants found within 1 Mb distance of the major histocompatibility complex (MHC) region were excluded. Either minimum sample size (N>20000) or multiple cohorts (≥23) was required as necessary criteria for processing results from a specific sex-stratified, and/or population group-stratified meta-analysis.

Significant variants were identified using the following threshold criteria. *i* variants with significant interaction effect (*P*_M1_GxE_<5e-9, FDR<0.05) and *j* variants with significant joint effect (*P*_M1_G,GxE_<5e-9, FDR<0.05) were filtered as top variants. Additionally, *k* top variants for the interaction effect were identified using a 2-step method: identifying first *z* variants by the marginal effect (*P*_M2_G_<1e-5) and then filtering these by the interaction effect (*P*_M1_GxE_<0.05/N_G,_ FDR_GxE_<0.05) where N_G_ is the number of independent tests calculated using principal components analysis on the *z* variants. This 2-step method was incorporated to increase power for detecting interactions^67^. This design was executed to maintain both stringent threshold criteria and incorporate false discovery correction implemented by the Benjamini-Hochberg method.

All such *i+j+k* significant variants were narrowed down to loci based on 500 kilobase (kb) regions. Finally, within these regions independent lead variants were identified as the top significant variant within the locus, subsequently defining variants in LD as those with linkage disequilibrium (LD) r^2^ threshold<0.1 using TOPMed-imputed 1000G reference panels. If variants were missing in the LD panels, then the most significant variant within each 500kb region was retained for combined sex meta-analyses results.

### Prioritizing Novel Sleep Duration Interaction Loci

Significant independent loci were subsequently filtered to prioritize gene-sleep duration interaction loci. From the 1df interaction test, *X* interaction loci were prioritized as those not found within 1Mb of previously identified gene-sleep duration loci for BP^8^. Loci were annotated as whether novel for BP genetic architecture, or not, by checking for overlap with 1Mb of previous GWAS variants (Supplementary Table S3).

For the 2df test, first loci were filtered to those variants not found within 1Mb of previous GWAS identified variants for BP traits, and with insignificant marginal effect (*P*_M2_G_ >5e-09, FDR_M2_G_ >0.05). From these variants, *Y* loci were prioritized as driven by interaction if they harbored a stronger interaction effect relative to the main effect (*P*_M1_GxE <_ *P*_M1_G_), and *Z* loci deemed as supported (but not driven) by interaction if this was not true.

Thus, collectively *X+Y* gene-sleep duration interaction loci were highlighted, alongside secondarily *Z* loci supported by interaction.

### Heterogeneity by Sex

To test for interaction effects showing evidence of heterogeneity by sex (p<0.05/*Q*), two-sample Z-tests assuming independence, were conducted for each of the top interaction loci and adjusted for multiple testing.

### Mapped Protein Coding Genes

Gene mapping prioritized protein-coding genes for downstream interpretation. Variants directly overlapping protein-coding gene regions were top priority criteria for gene assignment. For intergenic variants nearest distance to transcription start site (TSS) or gene start/end site was queried from Open Target Genetics v22.10^17^ or MyGene.Info using Python package *mygene* v3.2.2 (https://github.com/biothings/mygene.py). Variant mapping annotations were additionally noted from Open Target Genetics, Functional Annotation of Variants – Online Resource v2.0 (FAVOR)^11^, HaploReg v4.2 (https://pubs.broadinstitute.org/mammals/haploreg/haploreg.php), BRAVO variant browser (https://bravo.sph.umich.edu/freeze8/hg38/), Functional Mapping and Annotation of Genome-wide Association Studies v1.5.6 (FUMA)^12^, and MyGene.Info.

### Functional Annotations

At the variant level, FAVOR was queried to annotate deleteriousness or functionality scores^11^, and RegulomeDB v2.2 was used to extract aggregate regulatory function evidence scores, along with chromatin state, DNA accessibility, overlap with transcription factor (TF) binding sites or TF motifs, and expression quantitative trait loci (eQTL)^10^. At the genomic region level, FUMA’s SNP2GENE pipeline was used to annotate a comprehensive list of genes for each top locus, incorporating positional, chromatin interaction (FDR <=1e-6, 250bp upstream - 500 bp downstream of TSS), and GTEXv8 eQTL evidence (agreeing with RegulomeDB) with the top variant or its variants in LD (r2>0.1 within 500kb)^12^.

### Phenotypic Annotations

At the variant level, PheWeb, Open Target Genetics, Common Metabolic Diseases Knowledge Portal (https://hugeamp.org/), and Oxford Brain Imaging Genetics Server (BIG40) were queried for significant trait associations (p<5e-08) from past GWAS^13,15,17^. At the gene level, International Mouse Phenotyping Consortium release 19.1 (IMPC), Online Mendelian Inheritance in Man (OMIM; https://omim.org/), PheWeb, Phenotype-Genotype Integrator (PheGenI), and Open Target Genetics were queried for phenotypic annotations from mice knockout study results, involvement in Mendelian disorders, and significant trait associations (p<5e-08) ^14-17^. All STST and LTST mapped protein-coding genes were then queried using FUMA’s GENE2FUNC pipeline to identify significant (adjusted p-value<0.05) pathways and traits^12^. STRING v12.0 was additionally queried using medium confidence threshold (0.4) to note significantly (FDR<0.05) enriched traits or pathways to compare and contrast LTST and STST loci ^18^.

### Druggability Analysis

The Drug-Gene Interaction database (v4.2.0) was first utilized to identify druggability potential, with genes also annotated for implicated pathways and functions using the Kyoto Encyclopedia of Genes and Genomes database. Druggability target categories were annotated and all interacting drugs queried from reports across 43 databases (BaderLabGenes, CarisMolecularIntelligence, dGene, FoundationOneGenes, GO, HingoraniCasas, HopkinsGroom, HumanProteinAtlas, IDG, MskImpact, Oncomine, Pharos, RussLampel, Tempus, CGI, CIViC, COSMIC, CancerCommons, ChemblDrugs, ChemblInteractions, ClearityFoundationBiomarkers, ClearityFoundationClinicalTrial, DTC, DoCM, DrugBank, Ensembl, Entrez, FDA, GuideToPharmacology, JACX-CKB, MyCancerGenome, MyCancerGenomeClinicalTrial, NCI, OncoKB, PharmGKB, TALC, TEND, TTD, TdgClinicalTrial, Wikidata). Protein targets for available active ligands in ChEMBL were also noted. Gene targets were looked up in the druggable genome using the most recent druggable genome list established from the NIH Illuminating the Druggable Genome Project (https://github.com/druggablegenome/IDGTargets) available through the Pharos web platform. Lastly, FDA-approved drugs, late-stage clinical trials and disease indications were queried in the DrugBank, ChEMBL, ClinicalTrials.gov databases to provide results for the top MESH and DrugBank indications and clinical trials.

## Data Availability

All data produced in the present study are available upon reasonable request to the authors.

## ACKNOWLEDGMENTS

This project was largely supported by two grants from the U.S. National Heart, Lung, and Blood Institute (NHLBI), the National Institutes of Health, R01HL118305 and R01HL156991. Study-specific acknowledgements are included in the Supplementary Note. The research on the Million Veteran Program, was supported by award, CX001897 (PI A.M. Hung) from BLR&D, VA Office of R&D. This publication does not represent the views of the Department of Veteran Affairs or the United States Government. Acknowledgment of the Million Veteran Program leadership and staff contributions can be found in the Supplementary Note under MVP core acknowledgement.

## Notes

Conflict of Interest/Disclosures: C.L.M. has received funding from AstraZeneca not related to the current study. B.M.P. serves on the steering committee of the Yale Open Data Access Project funded by Johnson & Johnson. D.C. receives consultancy fees from Roche Diagnostics and Trimedics and speaker fees from Servier. D.A.L. has received support from Medtronic LTD and Roche Diagnostics for biomarker research not related to the current study. The remaining authors declare no competing interests.

### Competing Interest Statement

C.L.M. has received funding from AstraZeneca not related to the current study. B.M.P. serves on the steering committee of the Yale Open Data Access Project funded by Johnson & Johnson. D.C. receives consultancy fees from Roche Diagnostics and Trimedics and speaker fees from Servier. D.A.L. has received support from Medtronic LTD and Roche Diagnostics for biomarker research not related to the current study. The remaining authors declare no competing interests.

## REFERENCES

1. Makarem, N. et al. Sleep Duration and Blood Pressure: Recent Advances and Future Directions. Curr Hypertens Rep 21, 33 (2019).

2. Kanki, M. et al. Poor sleep and shift work associate with increased blood pressure and inflammation in UK Biobank participants. Nat Commun 14, 7096 (2023).

3. Kario, K. Sleep and nocturnal hypertension: Genes, environment, and individual profiles. J Clin Hypertens (Greenwich) 24, 1263–1265 (2022).

4. Bock, J.M., Vungarala, S., Covassin, N. & Somers, V.K. Sleep Duration and Hypertension: Epidemiological Evidence and Underlying Mechanisms. Am J Hypertens 35, 3–11 (2022).

5. Matsubayashi, H. et al. Long sleep duration and cardiovascular disease: Associations with arterial stiffness and blood pressure variability. J Clin Hypertens (Greenwich) 23, 496–503 (2021).

6. Cui, H. et al. Relationship of sleep duration with incident cardiovascular outcomes: a prospective study of 33,883 adults in a general population. BMC Public Health 23, 124 (2023).

7. Warren, H. et al. Genome-wide analysis in over 1 million individuals reveals over 2,000 independent genetic signals for blood pressure. (Research Square, 2022).

8. Wang, H. et al. Multi-ancestry genome-wide gene-sleep interactions identify novel loci for blood pressure. Mol Psychiatry 26, 6293–6304 (2021).

9. Kraft, P., Yen, Y.C., Stram, D.O., Morrison, J. & Gauderman, W.J. Exploiting gene-environment interaction to detect genetic associations. Hum Hered 63, 111–9 (2007).

10. Boyle, A.P. et al. Annotation of functional variation in personal genomes using RegulomeDB. Genome Res 22, 1790–7 (2012).

11. Zhou, H. et al. FAVOR: functional annotation of variants online resource and annotator for variation across the human genome. Nucleic Acids Res 51, D1300–D1311 (2023).

12. Watanabe, K., Taskesen, E., van Bochoven, A. & Posthuma, D. Functional mapping and annotation of genetic associations with FUMA. Nat Commun 8, 1826 (2017).

13. Smith, S.M. et al. An expanded set of genome-wide association studies of brain imaging phenotypes in UK Biobank. Nat Neurosci 24, 737–745 (2021).

14. Groza, T. et al. The International Mouse Phenotyping Consortium: comprehensive knockout phenotyping underpinning the study of human disease. Nucleic Acids Res 51, D1038–D1045 (2023).

15. Gagliano Taliun, S.A. et al. Exploring and visualizing large-scale genetic associations by using PheWeb. Nat Genet 52, 550–552 (2020).

16. Ramos, E.M. et al. Phenotype-Genotype Integrator (PheGenI): synthesizing genome-wide association study (GWAS) data with existing genomic resources. Eur J Hum Genet 22, 144–7 (2014).

17. Ghoussaini, M. et al. Open Targets Genetics: systematic identification of trait-associated genes using large-scale genetics and functional genomics. Nucleic Acids Res 49, D1311–D1320 (2021).

18. Szklarczyk, D. et al. STRING v11: protein-protein association networks with increased coverage, supporting functional discovery in genome-wide experimental datasets. Nucleic Acids Res 47, D607–D613 (2019).

19. Kavousi, M. et al. Multi-ancestry genome-wide study identifies effector genes and druggable pathways for coronary artery calcification. Nat Genet 55, 1651–1664 (2023).

20. Freshour, S.L. et al. Integration of the Drug-Gene Interaction Database (DGIdb 4.0) with open crowdsource efforts. Nucleic Acids Res 49, D1144–D1151 (2021).

21. Zdrazil, B. et al. The ChEMBL Database in 2023: a drug discovery platform spanning multiple bioactivity data types and time periods. Nucleic Acids Res (2023).

22. Wishart, D.S. et al. DrugBank 5.0: a major update to the DrugBank database for 2018. Nucleic Acids Res 46, D1074–D1082 (2018).

23. Yamada, N. et al. Mutant KCNJ3 and KCNJ5 Potassium Channels as Novel Molecular Targets in Bradyarrhythmias and Atrial Fibrillation. Circulation 139, 2157–2169 (2019).

24. Choi, T.Y. et al. Cereblon Maintains Synaptic and Cognitive Function by Regulating BK Channel. J Neurosci 38, 3571–3583 (2018).

25. de Wit, J. & Ghosh, A. Specification of synaptic connectivity by cell surface interactions. Nat Rev Neurosci 17, 22–35 (2016).

26. Bond, H.M. et al. Early hematopoietic zinc finger protein-zinc finger protein 521: a candidate regulator of diverse immature cells. Int J Biochem Cell Biol 40, 848–54 (2008).

27. Sakuragi, T. & Nagata, S. Publisher Correction: Regulation of phospholipid distribution in the lipid bilayer by flippases and scramblases. Nat Rev Mol Cell Biol 24, 597 (2023).

28. Aldaz, C.M. & Hussain, T. WWOX Loss of Function in Neurodevelopmental and Neurodegenerative Disorders. Int J Mol Sci 21(2020).

29. Damon, D.H., teRiele, J.A. & Marko, S.B. Eph/ephrin interactions modulate vascular sympathetic innervation. Auton Neurosci 158, 65–70 (2010).

30. Huang, N. et al. Reprogramming an energetic AKT-PAK5 axis boosts axon energy supply and facilitates neuron survival and regeneration after injury and ischemia. Curr Biol 31, 3098–3114 e7 (2021).

31. Tinti, M. et al. ANIA: ANnotation and Integrated Analysis of the 14-3-3 interactome. Database (Oxford) 2014, bat085 (2014).

32. Klein, D.C. et al. 14-3-3 Proteins and photoneuroendocrine transduction: role in controlling the daily rhythm in melatonin. Biochem Soc Trans 30, 365–73 (2002).

33. Bousquet-Moore, D., Mains, R.E. & Eipper, B.A. Peptidylgycine alpha-amidating monooxygenase and copper: a gene-nutrient interaction critical to nervous system function. J Neurosci Res 88, 2535–45 (2010).

34. Singh, C., Rihel, J. & Prober, D.A. Neuropeptide Y Regulates Sleep by Modulating Noradrenergic Signaling. Curr Biol 27, 3796–3811 e5 (2017).

35. Lee, H.J., Stefan-Lifshitz, M., Li, C.W. & Tomer, Y. Genetics and epigenetics of autoimmune thyroid diseases: Translational implications. Best Pract Res Clin Endocrinol Metab 37, 101661 (2023).

36. Garcia-Marin, R. et al. Melatonin in the thyroid gland: regulation by thyroid-stimulating hormone and role in thyroglobulin gene expression. J Physiol Pharmacol 66, 643–52 (2015).

37. Shalaby, N.A. et al. JmjC domain proteins modulate circadian behaviors and sleep in Drosophila. Sci Rep 8, 815 (2018).

38. Ikegami, K., Refetoff, S., van Cauter, E. & Yoshimura, T. Interconnection between circadian clocks and thyroid function. Nat Rev Endocrinol 15, 590–600 (2019).

39. Borna, S. et al. Transmembrane adaptor protein WBP1L regulates CXCR4 signalling and murine haematopoiesis. J Cell Mol Med 24, 1980–1992 (2020).

40. Poller, W.C., Nahrendorf, M. & Swirski, F.K. Hematopoiesis and Cardiovascular Disease. Circ Res 126, 1061–1085 (2020).

41. Greenlund, I.M. & Carter, J.R. Sympathetic neural responses to sleep disorders and insufficiencies. Am J Physiol Heart Circ Physiol 322, H337–H349 (2022).

42. McAlpine, C.S. et al. Sleep modulates haematopoiesis and protects against atherosclerosis. Nature 566, 383–387 (2019).

43. Wang, Q. et al. mNeuCode Empowers Targeted Proteome Analysis of Arginine Dimethylation. Anal Chem 95, 3684–3693 (2023).

44. Xiao, H.B., Wang, Y.S., Luo, Z.F. & Lu, X.Y. SZSJ protects against insomnia by a decrease in ADMA level and an improvement in DDAH production in sleep-deprived rats. Life Sci 209, 97–102 (2018).

45. Fujiwara, T. et al. PLEKHM1/DEF8/RAB7 complex regulates lysosome positioning and bone homeostasis. JCI Insight 1, e86330 (2016).

46. Azeez, T.A. Osteoporosis and cardiovascular disease: a review. Mol Biol Rep 50, 1753–1763 (2023).

47. Zhang, S. et al. Felodipine blocks osteoclast differentiation and ameliorates estrogen-dependent bone loss in mice by modulating p38 signaling pathway. Exp Cell Res 387, 111800 (2020).

48. Kim, J.H., Moon, J.S., Yu, J. & Lee, S.K. Intracellular cytoplasm-specific delivery of SH3 and SH2 domains of SLAP inhibits TcR-mediated signaling. Biochem Biophys Res Commun 460, 603–8 (2015).

49. Nakajima, T. et al. HERG is protected from pharmacological block by alpha-1,2-glucosyltransferase function. J Biol Chem 282, 5506–13 (2007).

50. He, F. et al. The KCNH2 genetic polymorphism (1956, C>T) is a novel biomarker that is associated with CCB and alpha,beta-ADR blocker response in EH patients in China. PLoS One 8, e61317 (2013).

51. Ye, H. et al. Nebivolol induces distinct changes in profibrosis microRNA expression compared with atenolol, in salt-sensitive hypertensive rats. Hypertension 61, 1008–13 (2013).

52. Amado-Azevedo, J. et al. A CDC42-centered signaling unit is a dominant positive regulator of endothelial integrity. Sci Rep 7, 10132 (2017).

53. Wang, C., Zhang, Y., Wu, Y. & Xing, D. Developments of CRBN-based PROTACs as potential therapeutic agents. Eur J Med Chem 225, 113749 (2021).

54. Roman, V., Walstra, I., Luiten, P.G. & Meerlo, P. Too little sleep gradually desensitizes the serotonin 1A receptor system. Sleep 28, 1505–10 (2005).

55. Watts, S.W., Morrison, S.F., Davis, R.P. & Barman, S.M. Serotonin and blood pressure regulation. Pharmacol Rev 64, 359–88 (2012).

56. Vila-Pueyo, M. Targeted 5-HT(1F) Therapies for Migraine. Neurotherapeutics 15, 291–303 (2018).

57. Abrahamowicz, A.A., Ebinger, J., Whelton, S.P., Commodore-Mensah, Y. & Yang, E. Racial and Ethnic Disparities in Hypertension: Barriers and Opportunities to Improve Blood Pressure Control. Curr Cardiol Rep 25, 17–27 (2023).

58. Johnson, D.A., Jackson, C.L., Williams, N.J. & Alcantara, C. Are sleep patterns influenced by race/ethnicity - a marker of relative advantage or disadvantage? Evidence to date. Nat Sci Sleep 11, 79–95 (2019).

59. Irwin, M.R. Why sleep is important for health: a psychoneuroimmunology perspective. Annu Rev Psychol 66, 143–72 (2015).

60. Buysse, D.J. Sleep health: can we define it? Does it matter? Sleep 37, 9–17 (2014).

61. Dong, Q. et al. Familial natural short sleep mutations reduce Alzheimer pathology in mice. iScience 25, 103964 (2022).

62. Westerman, K.E. et al. GEM: scalable and flexible gene-environment interaction analysis in millions of samples. Bioinformatics 37, 3514–3520 (2021).

63. Winkler, T.W. et al. Quality control and conduct of genome-wide association meta-analyses. Nat Protoc 9, 1192–212 (2014).

64. Willer, C.J., Li, Y. & Abecasis, G.R. METAL: fast and efficient meta-analysis of genomewide association scans. Bioinformatics 26, 2190–1 (2010).

65. Manning, A.K. et al. Meta-analysis of gene-environment interaction: joint estimation of SNP and SNP x environment regression coefficients. Genet Epidemiol 35, 11–8 (2011).

66. Winkler, T.W. et al. EasyStrata: evaluation and visualization of stratified genome-wide association meta-analysis data. Bioinformatics 31, 259–61 (2015).

67. Gauderman, W.J. et al. Update on the State of the Science for Analytical Methods for Gene-Environment Interactions. Am J Epidemiol 186, 762–770 (2017).

