## Supplementary Note for "A Large-Scale Genome-Wide Study of Gene-Sleep Duration Interactions for Blood Pressure in 811,405 Individuals from Diverse Populations"

**Supplementary Notes**

**A. Study Descriptions**

**Age Gene/Environment Susceptibility Reykjavik Study (AGES):** The AGES Reykjavik study originally comprised a random sample of 30,795 men and women born in 1907-1935 and living in Reykjavik in 1967. A total of 19,381 people attended, resulting in a 71% recruitment rate. The study sample was divided into six groups by birth year and birth date within month. One group was designated for longitudinal follow up and was examined in all stages; another was designated as a control group and was not included in examinations until 1991. Other groups were invited to participate in specific stages of the study. Between 2002 and 2006, the AGES Reykjavik study re-examined 5,764 survivors of the original cohort who had participated before in the Reykjavik Study.

**ALSPAC:** Pregnant women resident in Avon, UK with expected dates of delivery 1st April 1991 to 31st December 1992 were invited to take part in the study (PMID 22507742, 22507743). The initial number of pregnancies enrolled is 14,541 (for these at least one questionnaire has been returned or a ‘Children in Focus’ clinic data had been attended by 19/07/99). Of these initial pregnancies, there was a total of 14,676 foetuses, resulting in 14,062 live births and 13,988 children who were alive at 1 year of age. Questionnaires were sent at regular intervals, and biological samples were taken from mothers and children including blood samples, from which DNA was extracted. Study data were collected and managed using REDCap electronic data capture tools hosted at the University of Bristol (PMID 18929686). REDCap (Research Electronic Data Capture) is a secure, web-based software platform designed to support data capture for research studies. The study website contains details of all the data that is available through a fully searchable data dictionary and variable search tool (<http://www.bristol.ac.uk/alspac/researchers/our-data>). Ethical approval for the study was obtained from the ALSPAC Ethics and Law Committee and the Local Research Ethics Committees. Consent for biological samples has been collected in accordance with the Human Tissue Act (2004). Informed consent for the use of data collected via questionnaires and clinics was obtained from participants following the recommendation of the ALSPAC Ethics and Law Committee at the time. Sleep duration was self-reported between July 2002 and May 2004 by ALSPAC mothers and during 25-year follow-up (November 2017 ~ July 2018) by ALSPAC children. Blood pressure traits were measured in the first focus on mothers clinic (December 2008 ~ July 2011) for ALSPAC mothers and in the focus clinic during follow-up (June 2015 ~ October 2017) for ALSPAC children when they were around 24-year-old. For eligible participants from the same family (i.e. ALSPAC mother-child pairs), we randomly selected one of them to be included in the analyses. Thus, our study relied on 3793 genetically unrelated participants of European descent.

**Atherosclerosis Risk in Communities Study (ARIC):** The ARIC study is a population-based prospective cohort study of cardiovascular disease sponsored by the National Heart, Lung, and Blood Institute (NHLBI). ARIC included 15,792 individuals, predominantly European American and African American, aged 45-64 years at baseline (1987-89), chosen by probability sampling from four US communities. Cohort members completed additional triennial follow-up examinations, a fifth exam in 2011-2013, a sixth exam in 2016-2017, and a seventh exam in 2018-2019, an eighth exam in 2020, and a ninth exam in 2022. The ARIC study has been described in detail previously (PMC8667593; Wright JD, Folsom AR, Coresh J, et al. The ARIC (Atherosclerosis Risk In Communities) Study: JACC Focus Seminar 3/8. J Am Coll Cardiol. 2021 Jun 15;77(23):2939-2959).

**Bogalusa Heart Study (BHS):** The BHS study is population-based panel study to investigate the early natural history of cardiovascular disease and risk factors from childhood to adulthood among a biracial sample (65% white and 35% African American) of residents from Bogalusa, Louisiana. The study was established in 1973 by Dr. Gerald Berenson. To date, 9 surveys were conducted in children and adolescents aged 4 to 17 years, and 11 surveys were conducted among adults aged 18 to 51 years who were examined previously as children. At each survey, a standard questionnaire was used by trained research staffs to collect participants’ information on family status, levels of education, income, medical history and health behaviors. Clinical measures were collected following stringent protocols. The current study included 630 participants who were born between 1959 and 1979, examined at least once in childhood, and surveyed during the follow-up visit between 2013 and 2016. Genome-wide genotypes were assayed using the Illumina Human610 BeadChip. The genotype data were further imputed to the TOPMed reference panel using the TOPMed Imputation Server following a stringent genotype imputation protocol developed by the University of Michigan.

**The Coronary Artery Risk Development in Young Adults (CARDIA):** CARDIA is a prospective multicenter study of the development and determinants of clinical and subclinical cardiovascular disease and their risk factors. It began in 1985-1986 with a group of 5,115 black and white men and women aged 18-30 years, recruited from four US centers. The recruitment was done from the total community in Birmingham, AL, from selected census tracts in Chicago, IL and Minneapolis, MN; and from the Kaiser Permanente health plan membership in Oakland, CA. The participants were selected so that there would be approximately the same number of people in subgroups of race, gender, education (high school or less and more than high school) and age (18-24 and 25-30). These same participants were asked to participate in follow-up examinations during 1987-1988 (Year 2), 1990-1991 (Year 5), 1992-1993 (Year 7), 1995-1996 (Year 10), 2000-2001 (Year 15), 2005-2006 (Year 20), 2010-2011 (Year 25), 2015-2016 (Year 30), and 2020-2022 (Year 35).

Data collection protocols have been described previously (PMID: 3204420). Weight, height, systolic blood pressure and diastolic blood pressure were measured using standard protocols with participants wearing light clothing and no shoes. Morning venous blood samples were obtained after an overnight fast of at least 8 hours. Total cholesterol and triglycerides were measured enzymatically, HDL-C was determined by precipitation with dextran sulfate magnesium chloride, and LDL cholesterol was calculated using the Friedewald equation. Fasting plasma glucose and insulin were measured with the hexokinase–ultraviolet and radioimmunoassay methods, respectively (Linco Research, St Charles, MO, USA).

Written informed consent was obtained from participants at each examination and all study protocols were approved by the institutional review boards of the participating institutions.

**Cameron County Hispanic Cohort (CCHC):** The CCHC (Cameron County Hispanic Cohort) is an ongoing, longitudinal study that was initiated in 2004 to investigate the burden of metabolic-related conditions and disparities in a Mexican American community. The ongoing study has recruited over 5000 participants through community-based research assistants who through a randomization process contact families living at the US-Mexico border in Cameron County, TX to voluntarily enroll in the program. They are then invited to visit the Clinical Research Unit located in facilities provided by Valley Baptist Medical Center, Brownsville, TX. Comprehensive clinical examinations and collection of specimens have been conducted and archived for further analysis. A subset of 4076 individuals have been genotyped using the Illumina MEGA array and the results have been imputed to TOPMed phase 8 reference data.

**The Cleveland Family Study (CFS):** is the largest family-based study of sleep apnea worldwide, consisting of 2,284 individuals (46% African American) from 361 families studied on up to 4 occasions over a period of 16 years. The study was begun in 1990 with the initial aims of quantifying the familial aggregation of sleep apnea. NIH renewals provided expansion of the original cohort (including increased minority recruitment) and longitudinal follow-up, with the last exam occurring in February 2006. 632 African Americans were genotyped on the Affymetrix array 6.0 platform through the CARe Consortium with suitable genotying quality control. A further 122 African-Americans had genotyping based on the Illumina OmniExpress + Exome platform. Genomes were imputed by TOPMed imputation server separately.

**Cardiovascular Health Study (CHS):** CHS is a population-based cohort study of risk factors for cardiovascular disease in adults 65 years of age or older conducted across four field centers (1). The original predominantly European ancestry cohort of 5,201 persons was recruited in 1989-1990 from random samples of the Medicare eligibility lists and an additional predominately African-American cohort of 687 persons was enrolled in 1992-93 for a total sample of 5,888. Blood samples were drawn from all participants at their baseline examination and DNA was subsequently extracted from available samples. European ancestry participants were excluded from the GWAS study sample due to prevalent coronary heart disease, congestive heart failure, peripheral vascular disease, valvular heart disease, stroke, or transient ischemic attack at baseline. After QC, genotyping was successful for 3271 European ancestry and 823 African-American participants. Data for this analysis were from the study visit in 1996-1997. CHS was approved by institutional review committees at each site and individuals in the present analysis gave informed consent including consent to use of genetic information for the study of cardiovascular disease.

1. Fried LP, Borhani NO, Enright P, Furberg CD, Gardin JM, Kronmal RA, et al. The Cardiovascular Health Study: design and rationale. Ann Epidemiol 1991; 1:263-76.

**CoLaus:** The CoLaus study is a population-based cohort of 6733 participants from Lausanne (Switzerland) aged 35-75 at recruitment (2003-2006). This study collects detailed and standardized phenotypes to investigate biological, genetic and environmental determinants of cardiovascular disease and their association with mental disorders.

Initiated in 2003, the main aims of the CoLaus study were:

- To better understand the epidemiology of cardiovascular risk factors and diseases in the Swiss population.
- To discover new genetic determinants of these conditions.
- To determine the nature of the associations between cardiovascular disease and mental disorders

Since then, three follow-up surveys have been completed (2009-2012, 2014-2016, 2018-2020) and a forth one is ongoing since 2022. A detailed description of the participants’ selection, inclusion criteria and methods used can be found in the following manuscript:

Firmann M, Mayor V, Marques Vidal P, Bochud M, Pécoud A, Hayoz D, Paccaud F, Preisig M, Song KS, Yuan X, et al. The CoLaus study: a population-based study to investigate the genetic determinants of cardiovascular risk factors and metabolic syndrome. BMC Cardiovascular Disorders. 2008; 8:6. PMID: 18366642; PMCID: [*PMC2311269*](https://www.ncbi.nlm.nih.gov/pmc/articles/PMC2311269/); DOI: 10.1186/1471-2261-8-6.

**DIAbetes COhoRtE (DIACORE):** is a German prospective cohort study of 3,000 adult European diabetes mellitus type 2 outpatients, with a primary focus on renal events. Four follow-up examinations were conducted within 10 years after baseline inclusion. At each visit including, a core phenotyping protocol was performed that included a standardized interview, a physical examination, the determination of standard laboratory parameters from serum, urine and whole blood in a central laboratory and biobanking of biomaterials. The design of the DIACAORE study (Dorhofer L, Lammert A, Krane V, et al. Study design of DIACORE (DIAbetes COhoRtE)—a cohort study of patients with diabetes mellitus type 2. BMC Med Genet. 2013;14:25) and exemplary results (Rheinberger M, Jung B, Segiet T, et al. Poor risk factor control in outpatients with diabetes mellitus type 2 in Germany: The DIAbetes COhoRtE (DIACORE) study. PLoS ONE 2019;14(3): e0213157) were published previously.

**DR’s EXTRA (Dose-Responses to Exercise Training):** The Dose-Responses to Exercise Training (DR’s EXTRA) study is a 4-year RCT on the health effects of regular physical exercise and a healthy diet in a population-based random sample of Finnish men and women aged 55-74 years living in the city of Kuopio in 2002. The 3000 men and women who were invited to participate in the study were obtained from the national population registry. Altogether 2062 men and women expressed their willingness to participate in the study, and 1479 of them attended the baseline measurements in 2005 - 2006. The prespecified exclusion criteria were medical or other conditions that prohibit engagement in exercise intervention or the assessments, as judged by a physician. After these exclusions, 1410 individuals aged 57-78 years at baseline in 2005-2006 were randomized into the resistance exercise, aerobic exercise, diet, combined resistance exercise and diet, combined aerobic exercise and diet, or control group.

**European Prospective Investigation into Cancer and Nutrition (EPIC)-Norfolk:** The EPIC-Norfolk study (DOI 10.22025/2019.10.105.00004) is a prospective population-based cohort study which recruited 25,639 men and women aged 40-79 years at baseline between 1993 and 1997 from 35 participating general practices in Norfolk, UK [PMID:10466767]. Individuals attended for a baseline health check including the provision of blood samples for concurrent and future analysis. Further health check visits have been conducted since the baseline visit. Participants have contributed information about their diet, lifestyle and health through questionnaires and health checks over two decades. Sleep information was collected at the second heath check that was conducted in 1998-2000 (ages 42-82 years) with 15,786 participants. DNA has been extracted from all EPIC participants and stored blood has been analysed for an extensive range of classical and novel biomarkers. Sample quality control was performed including gender check, relatedness check, and ancestry check. The Norwich Local Research Ethics Committee granted ethical approval for the study. All participants gave written informed consent.

**EstBB:** The population-based Estonian Biobank (EstBB) contains data for 213,000 inhabitants of Estonia (~20% of the adult population). For the participants, a comprehensive questionnaire (providing objective information, and information on, e.g., physical activity and diet) is filled out and DNA, plasma, and white blood cell samples are stored. The database is updated by regular linkage to the national health databases. The EstBB project is being conducted according to the Estonian Human Genes Research Act, and all participants have signed a broad informed consent form (PMID: 24518929).

**The Fenland Study:** The Fenland study (DOI 10.22025/2017.10.101.00001) is a population-based cohort study that uses objective measures of disease exposure to investigate the influence of diet, lifestyle and genetic factors on the development of diabetes and obesity. The first phase of the Fenland Study was conducted between 2005 and 2015, and the volunteers are recruited from general practice lists in and around Cambridgeshire (Cambridge, Ely, and Wisbech) in the United Kingdom from birth cohorts from 1950–1975. Participants who attended the first phase of the study were invited to phase 2 of the study between 2014 and 2020, and the sleep data is available at this phase.

**Framingham Heart Study (FHS):** FHS began in 1948 with the recruitment of an original cohort of 5,209 men and women (mean age 44 years; 55 percent women). In 1971 a second generation of study participants was enrolled; this cohort (mean age 37 years; 52% women) consisted of 5,124 children and spouses of children of the original cohort. A third generation cohort of 4,095 children of offspring cohort participants (mean age 40 years; 53 percent women) was enrolled in 2002-2005 and are seen every 4 to 8 years. Details of study designs for the three cohorts are summarized elsewhere. At each clinic visit, a medical history was obtained with a focus on cardiovascular content, and participants underwent a physical examination including measurement of height and weight from which BMI was calculated.

**Genetic and phenotypic determinants of blood pressure and other cardiovascular risk factors (GAPP):** GAPP is a population-based prospective cohort study involving a representative sample of initially healthy adults aged 25-41 years at baseline and residing in the Principality of Liechtenstein. Exclusion criteria were the presence of cardiovascular disease, diabetes, obstructive sleep apnea and a body mass index >35kg/m2. A standardized 12-lead ECG was obtained in all participants. Baseline characteristics were obtained in all participants using standard methodology. The main goal of GAPP is to assess the mechanisms for the development of cardiovascular risk factors over time.

**Genetic Epidemiology Network of Salt-Sensitivity (GenSalt)**: The GenSalt study is a unique NHLBI-sponsored family feeding-study designed to examine the interaction between genes and dietary sodium intake on BP. A detailed description of the GenSalt study design and participants has been reported previously^1^. Briefly, 3,142 participants from 633 Han families from rural, north China were ascertained through a proband with untreated pre-hypertension or stage-1 hypertension identified from a population-based BP screening. Individuals who had stage 2 hypertension, secondary hypertension, and a history of clinical cardiovascular disease or diabetes or were pregnant, heavy alcohol drinkers, or currently on a low-sodium diet or BP lowering medication were excluded from the study, with a total of 1,906 GenSalt probands and their siblings, spouses, and offspring eligible for the 7-day low sodium and 7-day high sodium dietary interventions. At baseline, a standard questionnaire was administered by a trained staff member to collect information on family pedigree, demographic characteristics, personal and family medical history, and lifestyle risk factors. Body weight and height were measured twice in light indoor clothing without shoes. Body mass index was calculated as weight in kilograms per height in square meters. BP was measured three times at the same time each morning during the three-day baseline examination by trained and certified. The mean of the 9 BP measures was used in subsequent analyses. Blood specimens were collected by venipuncture to measure lipids, creatinine, and other laboratory values. Among the 1,906 intervention participants, 1,881 underwent genome-wide genotyping and whole genome sequencing.

1. The GenSalt Collaborative Research Group. GenSalt: rationale, design, methods and baseline characteristics of study participants. *J Hyperten.*2007;21:639-646.
2. Perloff D, Grim C, Flack J, Frohglich ED, Hill M, McDonald M, Morgenstern BZ. Human blood pressure determination by sphygmomanometer. *Circulation.* 1993;88:2460–2470.

**Healthy Aging in Neighborhoods of Diversity across the Life Span (HANDLS):** HANDLS is a community-based, longitudinal epidemiologic study examining the influences of race and socioeconomic status (SES) on the development of age-related health disparities among a sample of socioeconomically diverse African American and White residents of Baltimore, Maryland. This unique study entering its 20^th^ year has longitudinally assessed physical parameters and also evaluate genetic, biologic, demographic, and psychosocial, parameters of African American and White participants in higher and lower SES to understand the driving factors behind persistent Black-White health disparities in overall longevity, age-associated disease, and cognitive decline. The study recruited 3,720 participants from Baltimore, MD with a mean age of 47.7 years, 2,200 African Americans and 1,520 whites, with 41% reporting household incomes below the 125% poverty delimiter. Genotyping was done on a subset of self-reporting African American participants by the Laboratory of Neurogenetics, National Institute on Aging, National Institutes of Health (NIH). A larger genotyping effort included a small subset of self-reporting European ancestry samples. This research was supported by the Intramural Research Program of the NIH, NIA Project number AG000513.

**Hispanic Community Health Study/Study of Latinos (HCHS/SOL):** The HCHS/SOL is a multicenter prospective cohort of 16,000 Hispanic/Latino adults designed to investigate the role of acculturation in disease, and to identify other traits that impact Hispanic/Latino health. HCHS/SOL is the most diverse and comprehensive study of Hispanic/Latino health, with participants of Cuban, Puerto Rican, Dominican, Mexican or Central/South American origin. Participants were recruited through four sites affiliated with San Diego State University, Northwestern University in Chicago, Albert Einstein College of Medicine in Bronx, New York, and the University of Miami, using a census block and household sampling design. Study participants who were self-identified Hispanic/Latino and aged 18-74 years underwent extensive psycho-social, clinical assessments, and biospecimen collection during the baseline visit (2008-2011). A re-examination of the HCHS/SOL cohort was conducted during 2015-2017, and visit 3, which began in 2020, is ongoing. Annual telephone follow-up interviews have been conducted since study inception to determine health outcomes of interest. (dbGaP study accession number: phs000555).

**HCS (Hunter Community Study):** The HCS is a community-based longitudinal investigation that was commenced in Australia in 2004-2005. The study aims to investigate retired and near-retired persons by sampling older Australians aged 55–85, randomly selected from electoral rolls in a regional area on the heavily populated east coast (New South Wales). There were 3253 participants who completed at least some baseline measures. Follow-up was obtained through health record linkage up to 2017, representing over 10 years of health outcomes and hospitalizations.

**Insulin Resistance Atherosclerosis Study Family Study (IRASFS):** The IRASFS was a family study designed to examine the genetic and epidemiologic basis of glucose homeostasis traits and abdominal adiposity. Briefly, self-reported Mexican American pedigrees were recruited in San Antonio, TX and San Luis Valley, CO. Probands with large families were recruited from the initial non-family-based IRAS, which was modestly enriched for impaired glucose tolerance and T2D. Inclusion of IRASFS data is limited to 1040 normoglycemic individuals in 88 pedigrees with genotype data from the Illumina OmniExpress and Omni 1S arrays and imputation to the 1000 Genome Integrated Reference Panel (phase I).

**JHS (Jackson Heart Study):** The JHS is a longitudinal, community-based observational cohort study of 5,306 adults investigating the role of environmental and genetic factors in the development of cardiovascular disease in African Americans. Between 2000 and 2004, participants were recruited from a tri-county area (Hinds, Madison, and Rankin Counties) that encompasses Jackson, MS. Details of the design and recruitment for the Jackson Heart Study cohort has been previously published [1-3]. Briefly, approximately 30% of participants were former members of the Atherosclerosis Risk in Communities (ARIC) study. The remainder were recruited by either 1) random selection from the Accudata list, 2) commercial listing, 3) a constrained volunteer sample, in which recruitment was distributed among defined demographic cells in proportions designed to mirror those in the overall population, or through the Jackson Heart Study Family Study.

1. Wyatt SB, Diekelmann N, Henderson F, Andrew ME, Billingsley G, Felder SH, et al. A community-driven model of research participation: the Jackson Heart Study Participant Recruitment and Retention Study. *Ethn Dis*. 2003;13(4):438-55. PubMed PMID: 14632263.

2. Taylor HA, Jr., Wilson JG, Jones DW, Sarpong DF, Srinivasan A, Garrison RJ, et al. Toward resolution of cardiovascular health disparities in African Americans: design and methods of the Jackson Heart Study. *Ethn Dis*. 2005;15(4 Suppl 6):S6-4-17. PubMed PMID: 16320381.

3. Fuqua SR, Wyatt SB, Andrew ME, Sarpong DF, Henderson FR, Cunningham MF, et al. Recruiting African-American research participation in the Jackson Heart Study: methods, response rates, and sample description. *Ethn Dis*. 2005;15(4 Suppl 6):S6-18-29. PubMed PMID: 16317982.

**Korean Genome and Epidemiology Study (KOGES):** KoGES is a large-scale prospective cohort study with a comprehensive range of phenotypic measures and biological samples collected on approximately 210,000 individuals. KoGES’s long-term objective is to develop comprehensive and applicable healthcare guidelines for common complex diseases in Koreans, reduce the burden of chronic diseases and improve the quality of life. KoGES includes population-based cohorts, the community-based Ansan and Ansung study, the urban community-based health examinee study, and the rural community-based cardiovascular disease association study. The cohorts consist of community-dwellers and participants recruited from the national health examinee registry, men and women, aged ≥ 40 years at baseline. A total of 72,000 samples were genotyped with KoreanChip, a customized array optimized for the Korean population, and imputed using IMPUTE4 with 1000 Genomes Project Phase 3 data and the Korean reference genome as a reference panel. Measures of the baseline recruitment, and only genotyped samples that met the following exclusion criteria were used in the analyses: low call rate (<97%), excessive heterozygosity, excessive singletons, gender discrepancy, and cryptic first-degree relatives. SNPs with low HWE p value (<10^−6^) or low call rate (<95%) were excluded. Variants with imputation quality score (IQS) < 0.8 and MAF <1% were excluded after imputation.

1. Kim, Yeonjung, Bok-Ghee Han, and KoGES Group. "Cohort profile: the Korean genome and epidemiology study (KoGES) consortium." International journal of epidemiology 46.2 (2017): e20-e20.
2. Moon, Sanghoon, et al. "The Korea Biobank Array: design and identification of coding variants associated with blood biochemical traits." Scientific reports 9.1 (2019): 1-11.
3. Nam, Kisung, Jangho Kim, and Seunggeun Lee. "Genome-wide study on 72,298 individuals in Korean biobank data for 76 traits." Cell Genomics 2.10 (2022): 100189.

**Lothian Birth Cohort 1936 (LBC1936):** LBC1936 consists of 1091 relatively healthy individuals, most of whom took part in the Scottish Mental Survey of 1947 at the age of ~11 years old. They were recruited to a study to determine influences on cognitive ageing at age ~70 years, when almost all lived independently in the Lothian region of Scotland. For this project, data was drawn from LBC1936 Wave 3 (when sleep data was obtained).They have taken part in six waves of testing in later life (at mean ages 70, 73, 76, 79, 82 and 85 years). At each wave they underwent a series of cognitive and physical tests.^1,2^

1. Deary IJ, Gow AJ, Pattie A, Starr JM. Cohort profile: the Lothian Birth Cohorts of 1921 and 1936. Int J Epidemiol 2012;41:1576-1584.

2. Taylor AM, Pattie A, Deary IJ. Cohort Profile Update: The Lothian Birth Cohorts of 1921 and 1936. Int J Epidemiol 2018;47:1042-1042r

**Lifelines**: (https://lifelines.nl/) Lifelines is a multi-disciplinary prospective population-based cohort study using a unique three-generation design to examine the health and health-related behaviors of 165,000 persons living in the North East region of The Netherlands. It employs a broad range of investigative procedures in assessing the biomedical, socio-demographic, behavioral, physical and psychological factors which contribute to the health and disease of the general population, with a special focus on multimorbidity. In addition, the Lifelines project comprises a number of cross-sectional sub-studies, which investigate specific age-related conditions. These include investigations into metabolic and hormonal diseases, including obesity, cardiovascular and renal diseases, pulmonary diseases and allergy, cognitive function and depression, and musculoskeletal conditions. All survey participants are between 18 and 90 years old at the time of enrollment. Recruitment has been going on since the end of 2006, and over 130,000 participants had been included by April 2013. At the baseline examination, the participants in the study were asked to fill in a questionnaire (on paper or online) before the first visit. During the first and second visit, the first or second part of the questionnaire, respectively, are checked for completeness, a number of investigations are conducted, and blood and urine samples are taken. Lifelines is a facility that is open for all researchers. Information on application and data access procedure is summarized on www.lifelines.nl. (Scholtens S, Smidt N, Swertz MA, Bakker SJ, Dotinga A, Vonk JM, et al. Cohort Profile: LifeLines, a three-generation cohort study and biobank. Int J Epidemiol. 2014 Dec 14.)

Lifelines was genotyped in 2 stages: the first stage consisted of ~15,000 participants and used the Illumina CytoSNP chip; the second stage included ~35,000 participants and used the Illumina GSA chip. Due to the differences in genotyping, these two groups are analysed separately. Samples that are present, or have close relatives in both groups, are excluded from the CytoSNP dataset prior to analysis.

**The Long Life Family Study (LLFS):** LLFS is a longitudinal, population-based multigenerational family cohort designed to study genetic, behavioral, and environmental factors in families exhibiting exceptional longevity. Families were sampled from four clinical centers: Boston University Medical Center in Boston, MA; Columbia College of Physicians and Surgeons in New York City, NY; the University of Pittsburgh in Pittsburgh, PA, USA; and the University of Southern Denmark, Denmark. The characteristics, recruitment, eligibility, and enrollment were previously described (PMID: 21258136, PMID: 34739053). The first clinical exam started in 2006 and recruited 4,953 individuals in 539 two-generational families that demonstrated clustering for exceptional survival in the upper generation. The second clinical exam (2014-2017) revisited 2,933 European descent individuals from 528 families. The third clinical exam (2021-) is recruiting the participants from second exam and a few new ones from the grandchild generation. The individuals were genotyped using ~2.3 million SNPs from the Illumina Omni chip, then imputed on Version R2 of the TOPMed reference panel using the Michigan Imputation Server (https://imputation.biodatacatalyst.nhlbi.nih.gov/#!), which used Eagle v2.4 for phasing and minimac4 v1.3.3 for imputation.

**Multi-Ethnic Study of Atherosclerosis (MESA):** The Multi-Ethnic Study of Atherosclerosis (MESA) is a study of the characteristics of subclinical cardiovascular disease and the risk factors that predict progression to clinically overt cardiovascular disease or progression of the subclinical disease. MESA consisted of a diverse, population-based sample of an initial 6,814 asymptomatic men and women aged 45-84. 38 percent of the recruited participants were white, 28 percent African American, 22 percent Hispanic, and 12 percent Asian, predominantly of Chinese descent. Participants were recruited from six field centers across the United States: Wake Forest University, Columbia University, Johns Hopkins University, University of Minnesota, Northwestern University and University of California - Los Angeles. Participants are being followed for identification and characterization of cardiovascular disease events, including acute myocardial infarction and other forms of coronary heart disease (CHD), stroke, and
congestive heart failure; for cardiovascular disease interventions; and for mortality. The first examination took place over two years, from July 2000 - July 2002. It was followed by five examination periods that were 17-20 months in length, including the recently completed Exam 6 (2016-2018). MESA Exam 7 will be completed by early 2024. Participants have been contacted every 9 to 12 months throughout the study to assess clinical morbidity and mortality. Informed consent was obtained for extensive data sharing (dbGaP) and genetic/omic studies, including candidate genes (NHLBI CARe), genome-wide scans (NHLBI SHARe), exome sequencing (NHBLI ESP) and, most recently, the NHLBI TOPMed program.

1. Bild DE, Bluemke DA, Burke GL, Detrano R, Diez Roux AV, Folsom AR, Greenland P, Jacob DR Jr, Kronmal R, Liu K, Nelson JC, O'Leary D, Saad MF, Shea S, Szklo M, Tracy RP. Multi-ethnic study of atherosclerosis: objectives and design. Am J Epidemiol. 2002 Nov 1;156(9):871-81. PubMed PMID: 12397006.

**Million Veteran Program (MVP):** The MVP is a mega-biobank that was launched in 2011 and supported entirely by the Veterans Health Administration Office of Research and Development in the United States. The MVP received ethical and study protocol approval from the VA Central Institutional Review Board (IRB) in accordance with the principles outlined in the Declaration of Helsinki. The specific design, initial demographics and quality-control procedures of the MVP have been detailed previously ([10.1016/j.jclinepi.2015.09.016](https://doi.org/10.1016/j.jclinepi.2015.09.016)).

**The Nagahama Study (NAGAHAMA):** Details on this cohort can be found in the profile paper for this study at https://link.springer.com/chapter/10.1007/978-981-16-5727-6_7. This is a large-scale genome cohort in Japan that commenced in 2007, as The Nagahama Prospective Genome Cohort for Comprehensive Human Bioscience. At the frequency of every 5 years, lifestyle, clinical, and environmental measurements were collected along with data on disease trajectories, and blood and urine samples.

**The Netherlands Epidemiology of Obesity study (NEO):** The NEO was designed for extensive phenotyping to investigate pathways that lead to obesity-related diseases. The NEO study is a population-based, prospective cohort study that includes 6,671 individuals aged 45–65 years, with an oversampling of individuals with overweight or obesity. At baseline, information on demography, lifestyle, and medical history have been collected by questionnaires. In addition, samples of 24-h urine, fasting and postprandial blood plasma and serum, and DNA were collected. Genotyping was performed using the Illumina HumanCoreExome chip, which was subsequently imputed to the 1000 genome reference panel. Participants underwent an extensive physical examination, including anthropometry, electrocardiography, spirometry, and measurement of the carotid artery intima-media thickness by ultrasonography. In random subsamples of participants, magnetic resonance imaging of abdominal fat, pulse wave velocity of the aorta, heart, and brain, magnetic resonance spectroscopy of the liver, indirect calorimetry, dual energy X-ray absorptiometry, or accelerometry measurements were performed. The collection of data started in September 2008 and completed at the end of September 2012. Participants are currently being followed for the incidence of obesity-related diseases and mortality.

### **Netherlands Study of Depression and Anxiety (NESDA):** The NESDA is an ongoing longitudinal cohort study to examine the prevalence, long-term course and consequences of depressive and anxiety disorders in the adult population. A detailed description of the study can be found elsewhere^1^. Briefly, a total of 2981 participants, consisting of a healthy control group, people with a history of depressive or anxiety disorder and people with current depressive and/or anxiety disorder, were recruited from community (19%), primary care (54%), and outpatient psychiatric clinics (27%), and included at the baseline assessment in 2004-2007. Inclusion criteria were a lifetime diagnosis of major depressive disorder or anxiety disorder, age 18-65 years, and self-reported western European ancestry. Excluded were those who were not fluent in Dutch, and those with a primary diagnosis of psychotic disorder, obsessive compulsive disorder, bipolar disorder, or severe substance use or dependence. Biological sample collection and biobanking procedures (i.e. blood sampling and DNA isolation) took place during the baseline visit and has been previously described in detail ^2^.

NESDA samples were genotyped using either the Perlegen-Affymetrix 5.0, or Affymetrix 6.0 genotyping chip and called with BirdSeed. SNPs were excluded if unmapped or mapped to multiple locations, call rate<95%, MAF<0.01, HWE p-value<10^-5^, unmapped to build 36, allele frequency difference with 1000Genomes reference >20%, or palindromic SNPs with allele frequency>35%. Samples were removed if call rate<90%, deviant heterozygosity, abs(PLINK F)>0.1, sex mismatch, unexpected relatedness, or non-Caucasian. Imputation was performed with Impute software using the 1000Genomes Phase 1 Integrated Release 3 ALL reference panel.

1. Penninx BW, Beekman AT, Smit JH, et al. The netherlands study of depression and anxiety (NESDA): Rationale, objectives and methods. *International journal of methods in psychiatric research*. 2008;17(3):121-140.

2. Boomsma DI, Willemsen G, Sullivan PF, et al. Genome-wide association of major depression: Description of samples for the GAIN major depressive disorder study: NTR and NESDA biobank projects. *European Journal of Human Genetics*. 2008;16(3):335.

**Study of Health in Pomerania (SHIP):** The Study of Health In Pomerania (SHIP) is a prospective longitudinal population-based cohort study in Mecklenburg-Western Pomerania assessing the prevalence and incidence of common diseases and their risk factors (PMID: 20167617 and 35348705). SHIP encompasses the two independent cohorts SHIP-START and SHIP-TREND. Participants aged 20 to 79 with German citizenship and principal residency in the study area were recruited from a random sample of residents living in the three local cities, 12 towns as well as 17 randomly selected smaller towns. Individuals were randomly selected stratified by age and sex in proportion to population size of the city, town or small towns, respectively. A total of 4,308 participants were recruited between 1997 and 2001 in the SHIP-START cohort. Between 2008 and 2012 a total of 4,420 participants were recruited in the SHIP-TREND cohort. Individuals were invited to the SHIP study center for a computer-assisted personal interviews and extensive physical examinations. The study protocol was approved by the medical ethics committee of the University of Greifswald. Oral and written informed consent was obtained from each of the study participants.

Genome-wide SNP-typing was performed using the Affymetrix Genome-Wide Human SNP Array 6.0 (SHIP-START samples), the Illumina Infinium HumanOmni2.5 BeadChip, or the Illumina Infinium Global Screening Array (SHIP-TREND samples). Array processing was carried out in accordance with the manufacturer’s standard recommendations. Genotypes were determined using the Birdseed2 clustering algorithm for SHIP-START, GenomeStudio Genotyping Module v1.0, and GenomeStudio 2.0 Genotyping Module (GenCall) for SHIP-TREND.

**Study of Women's Health Across the Nation (SWAN):** The **S**tudy of **W**omen’s Health **A**cross the **N**ation (SWAN) is a multi-site, multiracial/ethnic longitudinal study of women’s health designed to describe the biological, behavioral, and psychosocial characteristics that occur during midlife and the menopausal transition. In addition to characterizing reproductive aging, SWAN focuses on the impact of menopause on age-related chronic diseases, such as diabetes, cardiovascular disease, depression, bone loss and osteoporosis, as well as physical and cognitive functioning. The SWAN cohort was enrolled in 1996-97 and consists of 3302 community-based women from seven sites with data from five race/ethnic groups:  Black (n=935), Chinese (n=250), Hispanic (n=286), Japanese (n=281), and White (n=1550). To be eligible for enrollment women had to be aged 42 to 52 years old, have an intact uterus and at least one ovary, have had a menstrual period in the previous three months, and not be taking hormones. SWAN participants have completed 15 approximately annual follow-up visits since enrollment. During follow-up visits 5 and 6 (2001-2003), 1757 women from six sites were consented to provide genetic materials for whom extracted DNA was successfully obtained for 1536 women: Black (n=410), Chinese (n=151), Hispanic (n=0), Japanese (n=168), and White (n=807).  This analysis was done separately for Whites and Blacks.

Sowers MF, Crawford S, Sternfeld B, Morganstein DG, Gold EB, Greendale G, Evans D, Neer R, Matthews K, Sherman S, Lo A, Weiss G, Kelsey J. SWAN: A multi-center, multi-ethnic, community-based cohort study of women and the menopausal transition. In: Lobo RA, Kelsey JL, Marcus R, editors. Menopause: biology and pathobiology. San Diego, CA: Academic Press; 2000. p. 175-88

**TwinsUK:** The TwinsUK registry is a national register of adult twins recruited as volunteers without selecting for any particular disease or traits PMID: 31526404. It is among the most detailed omics and phenotypic BioResource worldwide, including over 14,000 twins comparable to the general population for lifestyle characteristics. Genome-wide genotypes were assayed using a combined Illumina Human310 + Illumina Human610 BeadChip. The genotype data were imputed to the TOPMed reference panel using the TOPMed Imputation Server following a stringent genotype imputation protocol developed by the University of Michigan. A subset of 2,827 of genotyped twins with overlapping sleep and BP data were included in the analysis.

All twins provided informed written consent and the study was approved by St Thomas’ Hospital Research Ethics Committee (REC Ref: EC04/015).

**UK Biobank (UKB):** UK Biobank (UKB, www.ukbiobank.ac.uk) is a large longitudinal biobank study in the United Kingdom which was established to improve understanding of the genetic and environmental causes of common diseases including cardiovascular diseases. In addition to self-reported disease outcomes and extensive health and life-style questionnaire data, UKB participants are being tracked through their NHS records and national registries (including cause of death and Hospital Episode Statistics). In 2017, UKB released the genotypes of 488,377 participants profiled with a custom SNP array. Genotyping QC was performed centrally by UKB, and genotypes imputed to Haplotype Reference Consortium (HRC) panel were released for 488,377 participants. Sample selection for the Gene-Lifestyle Interaction projects was based on available datasets for traits and lifestyle measures.

**Women’s Health Initiative (WHI):** is a long-term national health study that focuses on strategies for preventing common diseases such as heart disease, cancer and fracture in postmenopausal women. A total of 161,838 women aged 50–79 years old were recruited from
40 clinical centers in the US between 1993 and 1998(1, 2). WHI consists of an observational study, two clinical trials of postmenopausal hormone therapy (HT, estrogen alone or estrogen plus progestin), a calcium and vitamin D supplement trial, and a dietary modification trial. Study
recruitment and exclusion criteria have been described previously(1, 2). Recruitment was done through mass mailing to age-eligible women obtained from voter registration, driver’s license and Health Care Financing Administration or other insurance list, with emphasis on recruitment of minorities and older women. Exclusions included participation in other randomized trials, predicted survival < 3 years, alcoholism, drug dependency, mental illness and dementia. For
the CT, women were ineligible if they had a systolic BP > 200 mm Hg or diastolic BP > 105 mm Hg, a history of hypertriglyceridemia or breast cancer. Study protocols and consent forms were approved by the IRB at all participating institutions. Socio-demographic characteristics, lifestyle, medical history and self-reported medications were collected using standardized questionnaires at the screening visit.
Genome wide association study (GWAS) non-overlapping samples are composed of (a) a case-control study (WHI Genomics and Randomized Trials Network – GARNET, which included all coronary heart disease, stroke, venous thromboembolic events and selected diabetes cases that happened during the active intervention phase in the WHI HT clinical trials and aged matched controls), (b) women selected to be "representative" of the HT trial (mostly younger white HT subjects that were also enrolled in the WHI memory study - WHIMS) and (c) the WHI SNP Health Association Resource (WHI SHARe), a randomly selected sample of 8,515 African American and 3,642 Hispanic women from WHI. Genotyping was performed using
Affymetrix 6.0 (WHI-SHARe), HumanOmniExpressExome-8v1_B (WHIMS) and Illumina HumanOmni1-Quad v1-0 B (GARNET). Quality control of the GWAS data included variant and sample call rates >95%, and included identification of genetically related individuals. Principal components were computed using methods developed by Price et al(3). Imputation was performed using the TOPMed Imputation Server and freeze 8 reference multi-ethnic panel (build hg38). Due to some overlap of WHI participants with the TOPMed reference panel, we re-calculated the estimated imputation quality based on only the samples not on TOPMed reference panel, to account for the over-estimation of imputation quality given by the imputation software(4). Variants with an imputation quality (Rsq) <0.3 were filtering out. After QC and exclusions from analysis protocol, the number of women included in analysis is 4,423 whites for GARNET, 5,202 white for WHIMS, 7,919 for SHARe African American and 3,377 for SHARe Hispanics. Analyses were performed using LinGxEScanR software.

1.	Design of the Women's Health Initiative clinical trial and observational study. The Women's Health Initiative Study Group. Control Clin Trials. 1998;19(1):61-109. Epub 1998/03/11. doi: S0197245697000780 [pii]. PubMed PMID: 9492970.

2.	Anderson GL, Manson J, Wallace R, Lund B, Hall D, Davis S, Shumaker S, Wang CY, Stein E, Prentice RL. Implementation of the Women's Health Initiative study design. Ann Epidemiol. 2003;13(9 Suppl):S5-17. Epub 2003/10/25. PubMed PMID: 14575938.

3.	Price AL, Patterson NJ, Plenge RM, Weinblatt ME, Shadick NA, Reich D. Principal components analysis corrects for stratification in genome-wide association studies. Nat Genet. 2006;38(8):904-9. doi: 10.1038/ng1847. PubMed PMID: 16862161.

4.	Sun Q, Liu W, Rosen JD, Huang L, Pace RG, Dang H, Gallins PJ, Blue EE, Ling H, Corvol H, Strug LJ, Bamshad MJ, Gibson RL, Pugh EW, Blackman SM, Cutting GR, O'Neal WK, Zhou YH, Wright FA, Knowles MR, Wen J, Li Y, Cystic Fibrosis Genome P. Leveraging TOPMed imputation server and constructing a cohort-specific imputation reference panel to enhance genotype imputation among cystic fibrosis patients. HGG Adv. 2022;3(2):100090. doi: 10.1016/j.xhgg.2022.100090. PubMed PMID: 35128485; PMCID: PMC8804187.

**The Cardiovascular Risk in Young Finns Study (YFS):** The YFS is a population-based follow up-study started in 1980. The main aim of the YFS is to determine the contribution made by childhood lifestyle, biological and psychological measures to the risk of cardiovascular diseases in adulthood. In 1980, over 3,500 children and adolescents all around Finland participated in the baseline study. The follow-up studies have been conducted mainly with 3-year intervals. The latest 30-year follow-up study was conducted in 2010-11 (ages 33-49 years) with 2,063 participants. The study was approved by the local ethics committees (University Hospitals of Helsinki, Turku, Tampere, Kuopio and Oulu) and was conducted following the guidelines of the Declaration of Helsinki. All participants gave their written informed consent.

**B. Study Acknowledgments**

**Age Gene/Environment Susceptibility Reykjavik Study (AGES):** This study has been funded by NIH contract N01-AG012100, HSSN271201200022C, the NIA Intramural Research Program, an Intramural Research Program Award (ZIAEY000401) from the National Eye Institute, an award from the National Institute on Deafness and Other Communication Disorders (NIDCD) Division of Scientific Programs (IAA Y2-DC_1004-02), Hjartavernd (the Icelandic Heart Association), and the Althingi (the Icelandic Parliament). The study is approved by the Icelandic National Bioethics Committee, VSN: 00-063. The researchers are indebted to the participants for their willingness to participate in the study.

**ALSPAC:** The UK Medical Research Council and Wellcome (Grant ref: 217065/Z/19/Z) and the University of Bristol provide core support for ALSPAC. This publication is the work of the authors and Heming Wang will serve as guarantors for the contents of this paper. Genomewide genotyping data was generated by Sample Logistics and Genotyping Facilities at Wellcome Sanger Institute and LabCorp (Laboratory Corporation of America) using support from 23andMe. A comprehensive list of grants funding is available on the ALSPAC website (<http://www.bristol.ac.uk/alspac/external/documents/grant-acknowledgements.pdf>); This research was specifically funded by British Heart Foundation (SP/07/008/24066, CS/15/6/31468). We are extremely grateful to all the families who took part in this study, the midwives for their help in recruiting them, and the whole ALSPAC team, which includes interviewers, computer and laboratory technicians, clerical workers, research scientists, volunteers, managers, receptionists and nurses.

**Atherosclerosis Risk in Communities Study (ARIC):** The ARIC study has been funded in whole or in part with Federal funds from the National Heart, Lung, and Blood Institute, National Institutes of Health, Department of Health and Human Services, under Contract nos. (75N92022D00001, 75N92022D00002, 75N92022D00003, 75N92022D00004, 75N92022D00005). The authors thank the staff and participants of the ARIC study for their important contributions. Funding was also supported by R01HL087641 and R01HL086694; National Human Genome Research Institute contract U01HG004402; and National Institutes of Health contract HHSN268200625226C. Infrastructure was partly supported by Grant Number UL1RR025005, a component of the National Institutes of Health and NIH Roadmap for Medical Research.

**Bogalusa Heart Study (BHS):** The BHS has been supported by multiple grants from the National Institute of Health, including R01AG077000, RF1AG041200, R01AG062309, and R33AG057983 from the National Institute on Aging, and R21HL161718 from the National Heart, Lung, and Blood Institute. The BHS study is extremely grateful to the participants for their willingness to participate in the study.

**The Coronary Artery Risk Development in Young Adults (CARDIA):** The Coronary Artery Risk Development in Young Adults Study (CARDIA) is conducted and supported by the National Heart, Lung, and Blood Institute (NHLBI) in collaboration with the University of Alabama at Birmingham (HHSN268201800005I & HHSN268201800007I), Northwestern University (HHSN268201800003I), University of Minnesota (HHSN268201800006I), and Kaiser Foundation Research Institute (HHSN268201800004I). Genotyping was funded as part of the NHLBI Candidate-gene Association Resource (N01-HC-65226) and the NHGRI Gene Environment Association Studies (GENEVA) (U01-HG004729, U01-HG04424, and U01-HG004446).

**Cameron County Hispanic Cohort (CCHC):** The CCHC is funded by the National Institutes of Health (NIH) CTSA UL1 TR00371, R01 HL142302-05A1, R01 DK127084-01A1, R01AG078452-01A1, CPRIT RP230063. We are grateful to all participants and members of the study team without whom this work would not be possible.

**The Cleveland Family Study (CFS):** was supported by grants from the National Institutes of Health (HL46380, M01 RR00080-39, T32-HL07567, RO1-46380).

**Cardiovascular Health Study (CHS):** This CHS research was supported by NHLBI contracts HHSN268201200036C, HHSN268200800007C, HHSN268201800001C, N01HC55222, N01HC85079, N01HC85080, N01HC85081, N01HC85082, N01HC85083, N01HC85086, 75N92021D00006; and NHLBI grants U01HL080295, R01HL085251, R01HL087652, R01HL105756, R01HL103612, R01HL120393, and U01HL130114 with additional contribution from the National Institute of Neurological Disorders and Stroke (NINDS). Additional support was provided through R01AG023629 from the National Institute on Aging (NIA). A full list of principal CHS investigators and institutions can be found at CHS-NHLBI.org. The provision of genotyping data was supported in part by the National Center for Advancing Translational Sciences, CTSI grant UL1TR001881, and the National Institute of Diabetes and Digestive and Kidney Disease Diabetes Research Center (DRC) grant DK063491 to the Southern California Diabetes Endocrinology Research Center. The content is solely the responsibility of the authors and does not necessarily represent the official views of the National Institutes of Health.

**CoLaus:** The CoLaus study was supported by research grants from GlaxoSmithKline, the Faculty of Biology and Medicine of Lausanne (Switzerland), the Swiss National Science Foundation (grants 3200B0–105993, 3200B0-118308, 33CSCO-122661, 33CS30-139468, 33CS30-148401, 33CS30_177535 and 3247730_204523) and the Swiss Personalized Health Network (grant 2018DRI01).The authors would like to express their gratitude to the participants in the Lausanne CoLaus study.

**DIAbetes COhoRtE (DIACORE):** Cohort recruiting and management was funded by the KfH Stiftung Präventivmedizin e.V. (Carsten A. Böger). Genome-wide genotyping was funded the Else Kröner-Fresenius-Stiftung (2012_A147), the KfH Stiftung Präventivmedizin and the University Hospital Regensburg. The German Research Foundation supported this work (DFG BO 3815/4-1 to Iris M. Heid and Carsten A. Böger, and Project-ID 387509280 – SFB 1350 subproject C6 to Iris M. Heid). The computational work supervised by Iris M. Heid was funded by the German Research Foundation (Project-ID 387509280 – SFB 1350 Subproject C6, Project-ID 509149993 - TRR 374/1 Subproject C6, INF).

**Dose-Responses to Exercise Training (DR’s EXTRA):** The DR's EXTRA Study was supported by the Ministry of Education and Culture of Finland (722 and 627;2004-2011), Academy of Finland (102318; 104943;123885; 211119), Kuopio University Hospital, Finnish Diabetes Association, Finnish Foundations for Cardiovascular Research, Päivikki and Sakari Sohlberg Foundation, by European Commission FP6 Integrated Project (EXGENESIS); LSHM-CT-2004-005272, City of Kuopio and Social Insurance Institution of Finland (4/26/2010).

**European Prospective Investigation into Cancer and Nutrition (EPIC)-Norfolk:** The EPIC-Norfolk study (DOI 10.22025/2019.10.105.00004) has received funding from the Medical Research Council (MR/N003284/1 MC-UU_12015/1 and MC_UU_00006/1) and Cancer Research UK (C864/A14136). The genetics work in the EPIC-Norfolk study was funded by the Medical Research Council (MC_PC_13048). We are grateful to all the participants who have been part of the project and to the many members of the study teams at the University of Cambridge who have enabled this research.

**EstBB:** This work was supported by the Estonian Research Council grants PRG1911 and TK (TK214).

**The Fenland Study:** The Fenland Study (DOI 10.22025/2017.10.101.00001) is funded by the Medical Research Council (MC_UU_12015/1). We are grateful to all the volunteers and to the General Practitioners and practice staff for assistance with recruitment. We thank the Fenland Study Investigators, Fenland Study Co-ordination team and the Epidemiology Field, Data and Laboratory teams. We further acknowledge support for genomics from the Medical Research Council (MC_PC_13046).

**Framingham Heart Study (FHS):** This research was conducted in part using data and resources from the Framingham Heart Study of the National Heart Lung and Blood Institute of the National Institutes of Health and Boston University School of Medicine. The analyses reflect intellectual input and resource development from the Framingham Heart Study investigators participating in the SNP Health Association Resource (SHARe) project. This work was partially supported by the National Heart, Lung and Blood Institute's Framingham Heart Study (Contract Nos. N01-HC-25195 and HSN268201500001I) and its contract with Affymetrix, Inc for genotyping services (Contract No. N02-HL-6-4278). This research was partially supported by grant R01DK122503 R01DK122503from the National Institute of Diabetes and Digestive and Kidney Diseases (MPIs: Kari North, Anne Justice, and Ching-Ti Liu).

**Genetic and phenotypic determinants of blood pressure and other cardiovascular risk factors (GAPP):** The GAPP study was supported by the Liechtenstein Government, the Swiss Heart Foundation, the Swiss Society of Hypertension, the University of Basel, the University Hospital Basel, the Hanela Foundation, the Mach-Gaensslen Foundation, Schiller AG, and Novartis. We thank the GAPP staff and all GAPP study participants for their important contributions.

**Genetic Epidemiology Network of Salt-Sensitivity (GenSalt)**: This work was supported by a cooperative agreement project grant (U01HL072507, R01HL087263, and R01HL090682) from the National Heart, Lung and Blood Institute, National Institutes of Health, Bethesda, MD

**Healthy Aging in Neighborhoods of Diversity across the Life Span (HANDLS):** The Healthy Aging in Neighborhoods of Diversity across the Life Span (HANDLS) study was supported by the Intramural Research Program of the NIH, National Institute on Aging and the National Center on Minority Health and Health Disparities (project # Z01-AG000513 and human subjects protocol number 09-AG-N248). Data analyses for the HANDLS study utilized the high-performance computational resources of the Biowulf Linux cluster at the National Institutes of Health, Bethesda, MD. (http://biowulf.nih.gov; http://hpc.nih.gov)).

**The Hispanic Community Health Study/Study of Latinos (HCHS/SOL):** The HCHS/SOL is a collaborative study supported by contracts from the National Heart, Lung, and Blood Institute (NHLBI) to the University of North Carolina (HHSN268201300001I / N01-HC-65233), University of Miami (HHSN268201300004I / N01-HC-65234), Albert Einstein College of Medicine (HHSN268201300002I / N01-HC-65235), University of Illinois at Chicago (HHSN268201300003I / N01- HC-65236 Northwestern Univ), and San Diego State University (HHSN268201300005I / N01-HC-65237). The following Institutes/Centers/Offices have contributed to the HCHS/SOL through a transfer of funds to the NHLBI: National Institute on Minority Health and Health Disparities, National Institute on Deafness and Other Communication Disorders, National Institute of Dental and Craniofacial Research, National Institute of Diabetes and Digestive and Kidney Diseases, National Institute of Neurological Disorders and Stroke, NIH Institution-Office of Dietary Supplements. The Genetic Analysis Center at the University of Washington was supported by NHLBI and NIDCR contracts (HHSN268201300005C AM03 and MOD03).

**Hunter Community Study (HCS):** The authors would like to thank the men and women participating in the HCS as well as all the staff, investigators and collaborators who have supported or been involved in the project to date. The authors would also like to thank the Hunter Medical Research Institute who provided media support during the initial recruitment of participants; and Dr Anne Crotty, Prof. Rodney Scott and Associate Prof. Levi who provided financial support towards freezing costs for the long-term storage of participant blood samples.

**Insulin Resistance Atherosclerosis Study Family Study (IRASFS):** The IRASFS was supported by the National Heart Lung and Blood Institute (HL060944, HL061019, and HL060919). Genotyping and analysis for this study was supported by the GUARDIAN Consortium with grant support from the National Institute of Diabetes, Digestive, and Kidney Diseases (NIDDK; DK085175 and DK118062) and in part by UL1TR000124 (CTSI) and DK063491 (DRC). The authors thank study investigators, staff, and participants for their valuable contributions.

**Jackson Heart Study (JHS):** The Jackson Heart Study (JHS) is supported and conducted in collaboration with Jackson State University (HHSN268201800013I), Tougaloo College (HHSN268201800014I), the Mississippi State Department of Health (HHSN268201800015I) and the University of Mississippi Medical Center (HHSN268201800010I, HHSN268201800011I and HHSN268201800012I) contracts from the National Heart, Lung, and Blood Institute (NHLBI) and the National Institute on Minority Health and Health Disparities (NIMHD). The authors also wish to thank the staffs and participants of the JHS.

JHS disclaimer- The views expressed in this manuscript are those of the authors and do not necessarily represent the views of the National Heart, Lung, and Blood Institute; the National Institutes of Health; or the U.S. Department of Health and Human Services.

**Korean Genome and Epidemiology Study (KOGES):** Data were collected and supported by the National Research Institute of Health, Centers for Disease Control and Prevention, and Ministry for Health and Welfare, Republic of Korea. The authors thank the staff and participants of the KoGES study for their important contributions. Additional support was provided through grants the Brain Pool Plus (BP+, Brain Pool+) Program (2020H1D3A2A03100666) and Sejong Science Fellowship (2022R1C1C2006474) through the National Research Foundation of Korea (NRF) funded by the Ministry of Science and ICT.

**Lothian Birth Cohort 1936 (LBC1936):** The authors thank all LBC1936 study participants and research team members who have contributed, and continue to contribute, to ongoing studies. LBC1936 is supported by the Biotechnology and Biological Sciences Research Council (BBSRC), and the Economic and Social Research Council [BB/W008793/1] (which supports SEH), Age UK (Disconnected Mind project), and the University of Edinburgh. SRC is supported by a Sir Henry Dale Fellowship jointly funded by the Wellcome Trust and the Royal Society (221890/Z/20/Z). Genotyping was funded by the BBSRC (BB/F019394/1).

**Lifelines Cohort Study:**

Raul Aguirre-Gamboa (1), Patrick Deelen (1), Lude Franke (1), Jan A Kuivenhoven (2), Esteban A Lopera Maya (1), Ilja M Nolte (3), Serena Sanna (1), Harold Snieder (3), Morris A Swertz (1), Peter M. Visscher (3,4), Judith M Vonk (3), Cisca Wijmenga (1), Naomi Wray (4)

(1) *Department of Genetics, University of Groningen, University Medical Center Groningen, The Netherlands*
(2) *Department of Pediatrics, University of Groningen, University Medical Center Groningen, The Netherlands*
(3) *Department of Epidemiology, University of Groningen, University Medical Center Groningen, The Netherlands*
(4) *Institute for Molecular Bioscience, The University of Queensland, Brisbane, Queensland, Australia.*

The Lifelines Biobank initiative has been made possible by funding from the Dutch Ministry of Health, Welfare and Sport, the Dutch Ministry of Economic Affairs, the University Medical Center Groningen (UMCG the Netherlands), University of Groningen and the Northern Provinces of the Netherlands. The generation and management of GWAS genotype data for the Lifelines Cohort Study is supported by the UMCG Genetics Lifelines Initiative (UGLI). UGLI is partly supported by a Spinoza Grant from NWO, awarded to Cisca Wijmenga.

The authors wish to acknowledge the services of the Lifelines Cohort Study, the contributing research centers delivering data to Lifelines, and all the study participants.

**The Long Life Family Study (LLFS):** This work was supported by the National Institute on Aging (U01AG023746, U01AG023712, U01AG023749, U01AG023755, U01AG023744, and U19AG063893).

**Multi-Ethnic Study of Atherosclerosis (MESA):** MESA and the MESA SHARe projects are conducted and supported by the National Heart, Lung, and Blood Institute (NHLBI) in collaboration with MESA investigators. Support for MESA is provided by contracts 75N92020D00001, HHSN268201500003I, N01-HC-95159, 75N92020D00005, N01-HC-95160, 75N92020D00002, N01-HC-95161, 75N92020D00003, N01-HC-95162, 75N92020D00006, N01-HC-95163, 75N92020D00004, N01-HC-95164, 75N92020D00007, N01-HC-95165, N01-HC-95166, N01-HC-95167, N01-HC-95168, N01-HC-95169, UL1-TR-000040, UL1-TR-001079, and UL1-TR-001420, UL1TR001881, DK063491, R01HL105756 and R01HL15699. Funding for SHARe genotyping was provided by NHLBI Contract N02-HL-64278. Genotyping was performed at Affymetrix (Santa Clara, California, USA) and the Broad Institute of Harvard and MIT (Boston, Massachusetts, USA) using the Affymetrix Genome-Wide Human SNP Array 6.0. The authors thank the other investigators, the staff, and the participants of the MESA study for their valuable contributions. A full list of participating MESA investigators and institutes can be found at [http://www.mesa-nhlbi.org](https://nam02.safelinks.protection.outlook.com/?url=http%3A%2F%2Fwww.mesa-nhlbi.org%2F&data=04%7C01%7Cwpost%40jhmi.edu%7Ce250d1c265a847ec090f08d9fbc439a1%7C9fa4f438b1e6473b803f86f8aedf0dec%7C0%7C0%7C637817642593854966%7CUnknown%7CTWFpbGZsb3d8eyJWIjoiMC4wLjAwMDAiLCJQIjoiV2luMzIiLCJBTiI6Ik1haWwiLCJXVCI6Mn0%3D%7C3000&sdata=tVqOVyqCZnioV87T5M39EUUQjtfxVyF4i%2FhQtZTLbwY%3D&reserved=0).

**Million Veteran Program (MVP):** **VA Million Veteran Program: Core Acknowledgement for Publications**

**MVP Program Office**

- Sumitra Muralidhar, Ph.D., Program Director

US Department of Veterans Affairs, 810 Vermont Avenue NW, Washington, DC 20420

- Jennifer Moser, Ph.D., Associate Director, Scientific Programs

US Department of Veterans Affairs, 810 Vermont Avenue NW, Washington, DC 20420

- Jennifer E. Deen, B.S., Associate Director, Cohort & Public Relations

US Department of Veterans Affairs, 810 Vermont Avenue NW, Washington, DC 20420

**MVP Executive Committee**

- Co-Chair: Philip S. Tsao, Ph.D.

VA Palo Alto Health Care System, 3801 Miranda Avenue, Palo Alto, CA 94304

- Co-Chair: Sumitra Muralidhar, Ph.D.

US Department of Veterans Affairs, 810 Vermont Avenue NW, Washington, DC 20420

- J. Michael Gaziano, M.D., M.P.H.

VA Boston Healthcare System, 150 S. Huntington Avenue, Boston, MA 02130

- Elizabeth Hauser, Ph.D.

Durham VA Medical Center, 508 Fulton Street, Durham, NC 27705

- Amy Kilbourne, Ph.D., M.P.H.

VA HSR&D, 2215 Fuller Road, Ann Arbor, MI 48105

- Michael Matheny, M.D., M.S., M.P.H.

VA Tennessee Valley Healthcare System, 1310 24^th^ Ave. South, Nashville, TN 37212

- Dave Oslin, M.D.

Philadelphia VA Medical Center, 3900 Woodland Avenue, Philadelphia, PA 19104

**MVP Co-Principal Investigators**

- J. Michael Gaziano, M.D., M.P.H.

VA Boston Healthcare System, 150 S. Huntington Avenue, Boston, MA 02130

- Philip S. Tsao, Ph.D.

VA Palo Alto Health Care System, 3801 Miranda Avenue, Palo Alto, CA 94304

**MVP Core Operations**

- Lori Churby, B.S., Director, MVP Regulatory Affairs

VA Palo Alto Health Care System, 3801 Miranda Avenue, Palo Alto, CA 94304

- Stacey B. Whitbourne, Ph.D., Director, MVP Cohort Management

VA Boston Healthcare System, 150 S. Huntington Avenue, Boston, MA 02130

- Jessica V. Brewer, M.P.H., Director, MVP Recruitment & Enrollment

VA Boston Healthcare System, 150 S. Huntington Avenue, Boston, MA 02130

- Shahpoor (Alex) Shayan, M.S., Director, MVP Recruitment and Enrollment Informatics

VA Boston Healthcare System, 150 S. Huntington Avenue, Boston, MA 02130

- Luis E. Selva, Ph.D., Executive Director, MVP Biorepositories

VA Boston Healthcare System, 150 S. Huntington Avenue, Boston, MA 02130

- Saiju Pyarajan Ph.D., Director, Data and Computational Sciences

VA Boston Healthcare System, 150 S. Huntington Avenue, Boston, MA 02130

- Kelly Cho, M.P.H, Ph.D., Director, MVP Phenomics Data Core

VA Boston Healthcare System, 150 S. Huntington Avenue, Boston, MA 02130

- Scott L. DuVall, Ph.D., Director, VA Informatics and Computing Infrastructure (VINCI)

VA Salt Lake City Health Care System, 500 Foothill Drive, Salt Lake City, UT 84148

- Mary T. Brophy M.D., M.P.H., Director, VA Central Biorepository

VA Boston Healthcare System, 150 S. Huntington Avenue, Boston, MA 02130

- MVP Coordinating Centers
  - MVP Coordinating Center, Boston - J. Michael Gaziano, M.D., M.P.H.

VA Boston Healthcare System, 150 S. Huntington Avenue, Boston, MA 02130

- MVP Coordinating Center, Palo Alto – Philip S. Tsao, Ph.D.

VA Palo Alto Health Care System, 3801 Miranda Avenue, Palo Alto, CA 94304

- MVP Information Center, Canandaigua – Brady Stephens, M.S.

Canandaigua VA Medical Center, 400 Fort Hill Avenue, Canandaigua, NY 14424

- Cooperative Studies Program Clinical Research Pharmacy Coordinating Center, Albuquerque – Todd Connor, Pharm.D.; Dean P. Argyres, B.S., M.S.

New Mexico VA Health Care System, 1501 San Pedro Drive SE, Albuquerque, NM 87108

**MVP Publications and Presentations Committee**

- Co-Chair: Themistocles L. Assimes, M.D., Ph. D

VA Palo Alto Health Care System, 3801 Miranda Avenue, Palo Alto, CA 94304

- Co-Chair: Adriana Hung, M.D.; M.P.H

VA Tennessee Valley Healthcare System, 1310 24^th^ Ave. South, Nashville, TN 37212

- Co-Chair: Henry Kranzler, M.D.

Philadelphia VA Medical Center, 3900 Woodland Avenue, Philadelphia, PA 19104

**MVP Local Site Investigators**

- Samuel Aguayo, M.D., Phoenix VA Health Care System

650 E. Indian School Road, Phoenix, AZ 85012

- Sunil Ahuja, M.D., South Texas Veterans Health Care System

7400 Merton Minter Boulevard, San Antonio, TX 78229

- Kathrina Alexander, M.D., Veterans Health Care System of the Ozarks

1100 North College Avenue, Fayetteville, AR 72703

- Xiao M. Androulakis, M.D., Columbia VA Health Care System

6439 Garners Ferry Road, Columbia, SC 29209

- Prakash Balasubramanian, M.D., William S. Middleton Memorial Veterans Hospital

2500 Overlook Terrace, Madison, WI 53705

- Zuhair Ballas, M.D., Iowa City VA Health Care System

601 Highway 6 West, Iowa City, IA 52246-2208

- Elizabeth S. Bast, M.D., M.P.H., Miami VA Health Care System

1201 NW 16th Street, 11 GRC, Miami FL 33125

- Jean Beckham, Ph.D., Durham VA Medical Center

508 Fulton Street, Durham, NC 27705

- Sujata Bhushan, M.D., VA North Texas Health Care System

4500 S. Lancaster Road, Dallas, TX 75216

- Edward Boyko, M.D., VA Puget Sound Health Care System

1660 S. Columbian Way, Seattle, WA 98108-1597

- David Cohen, M.D., Portland VA Medical Center

3710 SW U.S. Veterans Hospital Road, Portland, OR 97239

- Louis Dellitalia, M.D., Birmingham VA Medical Center

700 S. 19th Street, Birmingham AL 35233

- Gerald Wayne Dryden, Jr., M.D., Ph.D., Louisville VA Medical Center

800 Zorn Avenue, Louisville, KY 40206

- L. Christine Faulk, M.D., Robert J. Dole VA Medical Center

5500 East Kellogg Drive, Wichita, KS 67218-1607

- Joseph Fayad, M.D., VA Southern Nevada Healthcare System

6900 North Pecos Road, North Las Vegas, NV 89086

- Daryl Fujii, Ph.D., VA Pacific Islands Health Care System

459 Patterson Rd, Honolulu, HI 96819

- Saib Gappy, M.D., John D. Dingell VA Medical Center

4646 John R Street, Detroit, MI 48201

- Frank Gesek, Ph.D., White River Junction VA Medical Center

163 Veterans Drive, White River Junction, VT 05009

- Jennifer Greco, M.D., Sioux Falls VA Health Care System

2501 W 22nd Street, Sioux Falls, SD 57105

- Michael Godschalk, M.D., Richmond VA Medical Center

1201 Broad Rock Blvd., Richmond, VA 23249

- Todd W. Gress, M.D., Ph.D., Hershel “Woody” Williams VA Medical Center

1540 Spring Valley Drive, Huntington, WV 25704

- Samir Gupta, M.D., M.S.C.S., VA San Diego Healthcare System

3350 La Jolla Village Drive, San Diego, CA 92161

- Salvador Gutierrez, M.D., Edward Hines, Jr. VA Medical Center

5000 South 5th Avenue, Hines, IL 60141

- John Harley, M.D., Ph.D., Cincinnati VA Medical Center

3200 Vine Street, Cincinnati, OH 45220

- Mark Hamner, M.D., Ralph H. Johnson VA Medical Center

109 Bee Street, Mental Health Research, Charleston, SC 29401

- Daniel J. Hogan, M.D., Bay Pines VA Healthcare System

10,000 Bay Pines Blvd Bay Pines, FL 33744

- Adriana Hung, M.D., M.P.H., VA Tennessee Valley Healthcare System

1310 24th Avenue, South Nashville, TN 37212

- Robin Hurley, M.D., W.G. (Bill) Hefner VA Medical Center

1601 Brenner Ave, Salisbury, NC 28144

- Pran Iruvanti, D.O., Ph.D., Hampton VA Medical Center

100 Emancipation Drive, Hampton, VA 23667

- Frank Jacono, M.D., VA Northeast Ohio Healthcare System

10701 East Boulevard, Cleveland, OH 44106

- Darshana Jhala, M.D., Philadelphia VA Medical Center

3900 Woodland Avenue, Philadelphia, PA 19104

- Scott Kinlay, M.B.B.S., Ph.D., VA Boston Healthcare System

150 S. Huntington Avenue, Boston, MA 02130

- Michael Landry, Ph.D., Southeast Louisiana Veterans Health Care System

2400 Canal Street, New Orleans, LA 70119

- Peter Liang, M.D., M.P.H., VA New York Harbor Healthcare System

423 East 23rd Street, New York, NY 10010

- Suthat Liangpunsakul, M.D., M.P.H., Richard Roudebush VA Medical Center

1481 West 10th Street, Indianapolis, IN 46202

- Jack Lichy, M.D., Ph.D., Washington DC VA Medical Center

50 Irving St, Washington, D. C. 20422

- Tze Shien Lo, M.D., Fargo VA Health Care System

2101 N. Elm, Fargo, ND 58102

- C. Scott Mahan, M.D., Charles George VA Medical Center

1100 Tunnel Road, Asheville, NC 28805

- Ronnie Marrache, M.D., VA Maine Healthcare System Center, Augusta, ME 04330
- Stephen Mastorides, M.D., James A. Haley Veterans’ Hospital

13000 Bruce B. Downs Blvd, Tampa, FL 33612

- Kristin Mattocks, Ph.D., M.P.H., Central Western Massachusetts Healthcare System

421 North Main Street, Leeds, MA 01053

- Paul Meyer, M.D., Ph.D., Southern Arizona VA Health Care System

3601 S 6th Avenue, Tucson, AZ 85723

- Jonathan Moorman, M.D., Ph.D., James H. Quillen VA Medical Center

Corner of Lamont & Veterans Way, Mountain Home, TN 37684

- Providencia Morales, R.N., Northern Arizona VA Health Care System

500 Highway 89 North, Prescott, AZ 86313

- Timothy Morgan, M.D., VA Long Beach Healthcare System

5901 East 7th Street Long Beach, CA 90822

- Maureen Murdoch, M.D., M.P.H., Minneapolis VA Health Care System

One Veterans Drive, Minneapolis, MN 55417

- Eknath Naik, M.D., Ph.D., West Palm Beach VA Medical Center,

7305 North Military Trail, West Palm Beach, FL 33410-6400

- James Norton, Ph.D., VA Health Care Upstate New York

113 Holland Avenue, Albany, NY 12208

- Olaoluwa Okusaga, M.D., Michael E. DeBakey VA Medical Center

2002 Holcombe Blvd, Houston, TX 77030

- Michael K. Ong, M.D., VA Greater Los Angeles Health Care System

11301 Wilshire Blvd, Los Angeles, CA 90073

- Kris Ann Oursler, M.D., Salem VA Medical Center

1970 Roanoke Blvd, Salem, VA 24153

- Ismene Petrakis, M.D., VA Connecticut Healthcare System

950 Campbell Avenue, West Haven, CT 06516

- Samuel Poon, M.D., Manchester VA Medical Center

718 Smyth Road, Manchester, NH 03104

- Emily Potter, Pharm.D., VA Eastern Kansas Health Care System

4101 S 4th Street Trafficway, Leavenworth, KS 66048

- Michael Rauchman, M.D., St. Louis VA Health Care System

915 North Grand Blvd, St. Louis, MO 63106

- Amneet S. Rai, Pharm.D., VA Sierra Nevada Health Care System

975 Kirman Avenue, Reno, NV 89502

- Richard Servatius, Ph.D., Syracuse VA Medical Center

800 Irving Avenue, Syracuse, NY 13210

- Satish Sharma, M.D., Providence VA Medical Center

830 Chalkstone Avenue, Providence, RI 02908

- River Smith, Ph.D., Eastern Oklahoma VA Health Care System

1011 Honor Heights Drive, Muskogee, OK 74401

- Peruvemba Sriram, M.D., N. FL/S. GA Veterans Health System

1601 SW Archer Road, Gainesville, FL 32608

- Patrick Strollo, Jr., M.D., VA Pittsburgh Health Care System

University Drive, Pittsburgh, PA 15240

- Neeraj Tandon, M.D., Overton Brooks VA Medical Center

510 East Stoner Ave, Shreveport, LA 71101

- Philip Tsao, Ph.D., VA Palo Alto Health Care System

3801 Miranda Avenue, Palo Alto, CA 94304-1290

- Gerardo Villareal, M.D., New Mexico VA Health Care System

1501 San Pedro Drive, S.E. Albuquerque, NM 87108

- Jessica Walsh, M.D., VA Salt Lake City Health Care System

500 Foothill Drive, Salt Lake City, UT 84148

- John Wells, Ph.D., Edith Nourse Rogers Memorial Veterans Hospital

200 Springs Road, Bedford, MA 01730

- Jeffrey Whittle, M.D., M.P.H., Clement J. Zablocki VA Medical Center

5000 West National Avenue, Milwaukee, WI 53295

- Mary Whooley, M.D., San Francisco VA Health Care System

4150 Clement Street, San Francisco, CA 94121

- Peter Wilson, M.D., Atlanta VA Medical Center

1670 Clairmont Road, Decatur, GA 30033

- Junzhe Xu, M.D., VA Western New York Healthcare System

3495 Bailey Avenue, Buffalo, NY 14215-1199

- Shing Shing Yeh, Ph.D., M.D., Northport VA Medical Center

79 Middleville Road, Northport, NY 11768

- Andrew W. Yen, M.D., VA Northern California Health Care System

10535 Hospital Way, Mather, CA 95655

**The Netherlands Epidemiology of Obesity study (NEO):** The authors of the NEO study thank all individuals who participated in the Netherlands Epidemiology in Obesity study, all participating general practitioners for inviting eligible participants and all research nurses for collection of the data. We thank the NEO study group, Petra Noordijk, Pat van Beelen and Ingeborg de Jonge for the coordination, lab and data management of the NEO study. The genotyping in the NEO study was supported by the Centre National de Génotypage (Paris, France), headed by Jean-Francois Deleuze. The NEO study is supported by the participating Departments, the Division and the Board of Directors of the Leiden University Medical Center, and by the Leiden University, Research Profile Area Vascular and Regenerative Medicine.

**Netherlands Study of Depression and Anxiety (NESDA):** The infrastructure for the NESDA study (www.nesda.nl) is funded through the Geestkracht program of the Netherlands Organisation for Health Research and Development (ZonMw, grant number 10-000-1002) and financial contributions by participating universities and mental health care organizations (VU University Medical Center, GGZ inGeest, Leiden University Medical Center, Leiden University, GGZ Rivierduinen, University Medical Center Groningen, University of Groningen, Lentis, GGZ Friesland, GGZ Drenthe, Rob Giel Onderzoekscentrum).

**Study of Health in Pomerania (SHIP):** SHIP is part of the Community Medicine Research net of the University of Greifswald, Germany, which is funded by the Federal Ministry of Education and Research (grants no. 01ZZ9603, 01ZZ0103, and 01ZZ0403), the Ministry of Cultural Affairs as well as the Social Ministry of the Federal State of Mecklenburg-West Pomerania, and the network ‘Greifswald Approach to Individualized Medicine (GANI_MED)’ funded by the Federal Ministry of Education and Research (grant 03IS2061A). Genome-wide data were supported by the Federal Ministry of Education and Research (grant no. 03ZIK012) and a joint grant from Siemens Healthcare, Erlangen, Germany and the Federal State of Mecklenburg- West Pomerania.

**Study of Women's Health Across the Nation (SWAN):** Study of Women’s Health Across the Nation (SWAN): The Study of Women’s Health Across the Nation (SWAN) has grant support from the National Institutes of Health (NIH), DHHS, through the National Institute on Aging (NIA), the National Institute of Nursing Research (NINR) and the NIH Office of Research on Women’s Health (ORWH) (Grants U01NR004061; U01AG012505, U01AG012535, U01AG012531, U01AG012539, U01AG012546, U01AG012553, U01AG012554, U01AG012495 and U19AG063720 and The SWAN Repository (U01AG017719)). The

content of this article is solely the responsibility of the authors and does not necessarily represent

the official views of the NIA, NINR, ORWH or the NIH.

Clinical Centers: University of Michigan, Ann Arbor – Carrie Karvonen-Gutierrez PI, 2021-present, Siobán Harlow, PI 2011 – 2021, MaryFran Sowers, PI 1994-2011; Massachusetts General Hospital, Boston, MA – Massachusetts General Hospital, Boston, MA – Sherri‐Ann Burnett‐Bowie, PI 2020 – Present; Joel Finkelstein, PI 1999 – 2020; Robert Neer, PI 1994 –1999; Rush University, Rush University Medical Center, Chicago, IL – Imke Janssen, PI 2020 –Present; Howard Kravitz, PI 2009 – 2020; Lynda Powell, PI 1994 – 2009; University of California, Davis/Kaiser – Elaine Waetjen and Monique Hedderson, PIs 2020 – Present; Ellen Gold, PI 1994 - 2020; University of California, Los Angeles – Arun Karlamangla, PI 2020 – Present; Gail Greendale, PI 1994 - 2020; Albert Einstein College of Medicine, Bronx, NY – Carol Derby, PI 2011 – present, Rachel Wildman, PI 2010 – 2011; Nanette Santoro, PI 2004 – 2010; University of Medicine and Dentistry – New Jersey Medical School, Newark – Gerson Weiss, PI 1994 – 2004; and the University of Pittsburgh, Pittsburgh, PA – Rebecca Thurston, PI 2020 – Present; Karen Matthews, PI 1994 - 2020.

NIH Program Office: National Institute on Aging, Bethesda, MD – Rosaly Correa-de-Araujo 2020 - present; Chhanda Dutta 2016- present; Winifred Rossi 2012–2016; Sherry Sherman 1994 – 2012; Marcia Ory 1994 – 2001; National Institute of Nursing Research, Bethesda, MD – Program Officers. Central Laboratory: University of Michigan, Ann Arbor – Daniel McConnell (Central Ligand Assay Satellite Services). NIA Biorepository - Rosaly Correa-de-Araujo 2019 - Present; SWAN Repository: University of Michigan, Ann Arbor – Siobán Harlow 2013- present; Dan McConnell 2011 - 2013; MaryFran Sowers 2000 – 2011. Coordinating Center: University of Pittsburgh, Pittsburgh, PA – Maria Mori Brooks, PI 2012 - present; Kim Sutton-Tyrrell, PI 2001 – 2012; New England Research Institutes, Watertown, MA - Sonja McKinlay, PI 1995 – 2001. Steering Committee:	Susan Johnson, Current Chair; Chris Gallagher, Former Chair. We thank the study staff at each site and all the women who participated in SWAN.

**TwinsUK:** The Department of Twin Research receives support from grants from the Wellcome Trust (212904/Z/18/Z) and the Medical Research Council (MRC)/British Heart Foundation (BHF) Ancestry and Biological Informative Markers for Stratification of Hypertension (AIM-HY; MR/M016560/1), European Union, Chronic Disease Research Foundation (CDRF), Zoe Global Ltd., the NIHR Clinical Research Facility and Biomedical Research Centre (based at Guy’s and St Thomas’ NHS Foundation Trust in partnership with King’s College London). C.M. is funded by the Chronic Disease Research Foundation; M.M. is funded by the National Institute for Health Research (NIHR)-funded BioResource, Clinical Research Facility and Biomedical Research Centre based at Guy’s and St Thomas’ NHS Foundation Trust in partnership with King’s College London.

**UK Biobank (UKB):** This research has been conducted using the UK Biobank Resource under Application Number 8343. This research used data assets made available by National Safe Haven as part of the Data and Connectivity National Core Study, led by Health Data Research UK in partnership with the Office for National Statistics and funded by UK Research and Innovation (grant ref MC_PC_20029). Copyright © (2022), NHS Digital.  Re-used with the permission of the NHS Digital [and/or UK Biobank].  All rights reserved.

**Women’s Health Initiative (WHI):** The WHI program is funded by the National Heart, Lung, and Blood Institute, National Institutes of Health, U.S. Department of Health and Human Services through contracts HHSN268201600018C, HHSN268201600001C, HHSN268201600002C,
HHSN268201600003C, and HHSN268201600004C. The authors thank the WHI investigators
and staff for their dedication, and the study participants for making the program possible. A full
listing of WHI investigators can be found at: https://www-whi-org.s3.us-west-
2.amazonaws.com/wp-content/uploads/WHI-Investigator-Long-List.pdf. Additional support to NF was provided by NIH DK117445 and MD012765.

**The Cardiovascular Risk in Young Finns Study (YFS):** The Young Finns Study has been financially supported by the Academy of Finland: grants 286284, 134309 (Eye), 126925, 121584, 124282, 129378 (Salve), 117787 (Gendi), and 41071 (Skidi); the Social Insurance Institution of Finland; Kuopio, Tampere and Turku University Hospital Medical Funds (grant X51001); Juho Vainio Foundation; Paavo Nurmi Foundation; Finnish Foundation for Cardiovascular Research ; Finnish Cultural Foundation; Tampere Tuberculosis Foundation; Emil Aaltonen Foundation; Yrjö Jahnsson Foundation; Signe and Ane Gyllenberg Foundation; and Diabetes Research Foundation of Finnish Diabetes Association. The expert technical assistance in the statistical analyses by Leo-Pekka Lyytikäinen and Irina Lisinen is gratefully acknowledged.
