## Supplementary Figures for "A Large-Scale Genome-Wide Study of Gene-Sleep Duration Interactions for Blood Pressure in 811,405 Individuals from Diverse Populations"

**Supplementary Fig S1.** **Quantile-Quantile (QQ) Plots**

**Note:** Both lifestyle exposures (E) Long Total Sleep Time (LTST) and Short Total Sleep Time (STST) are depicted. All 3 Blood Pressure (BP) outcome traits are plotted – Systolic (SBP), Diastolic (DBP), and Pulse Pressure (PP). All population-group specific results - African (AFR), East Asian (EAS), European (EUR), Hispanic/Latino (HIS), and South Asian (SAS) - are denoted along with cross population meta-analysis (CPMA). The top row of plots in each figure A-F depicts 1 degree of freedom Interaction Test results, while the bottom row depicts 2 degree of freedom Joint Test results.

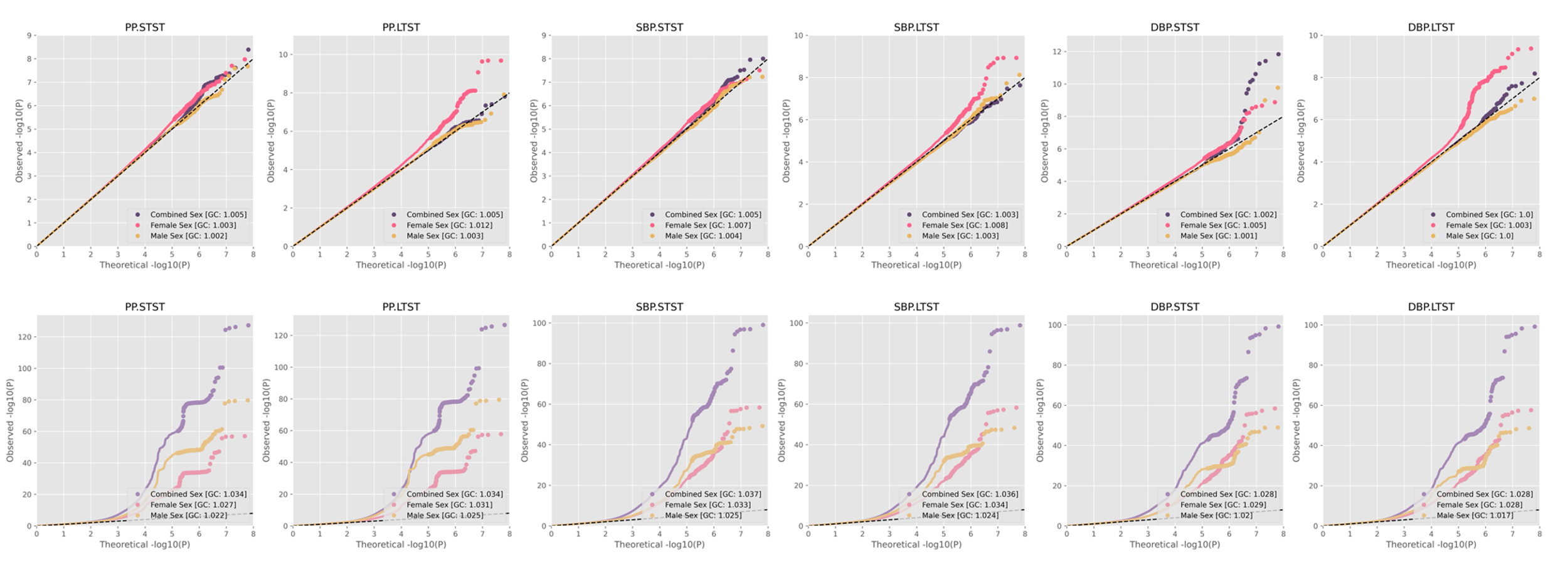
A. Cross Population Meta-Analysis

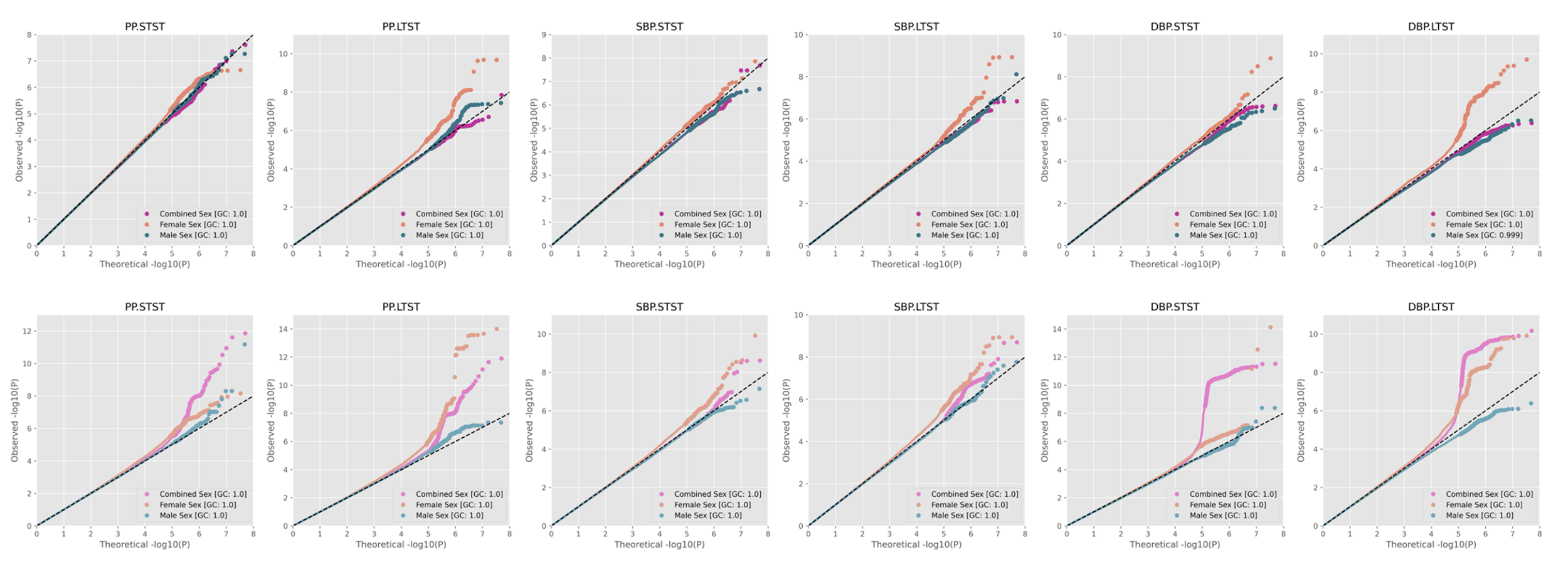
B. AFR-Specific Meta-Analysis

C. EAS-Specific Meta-Analysis

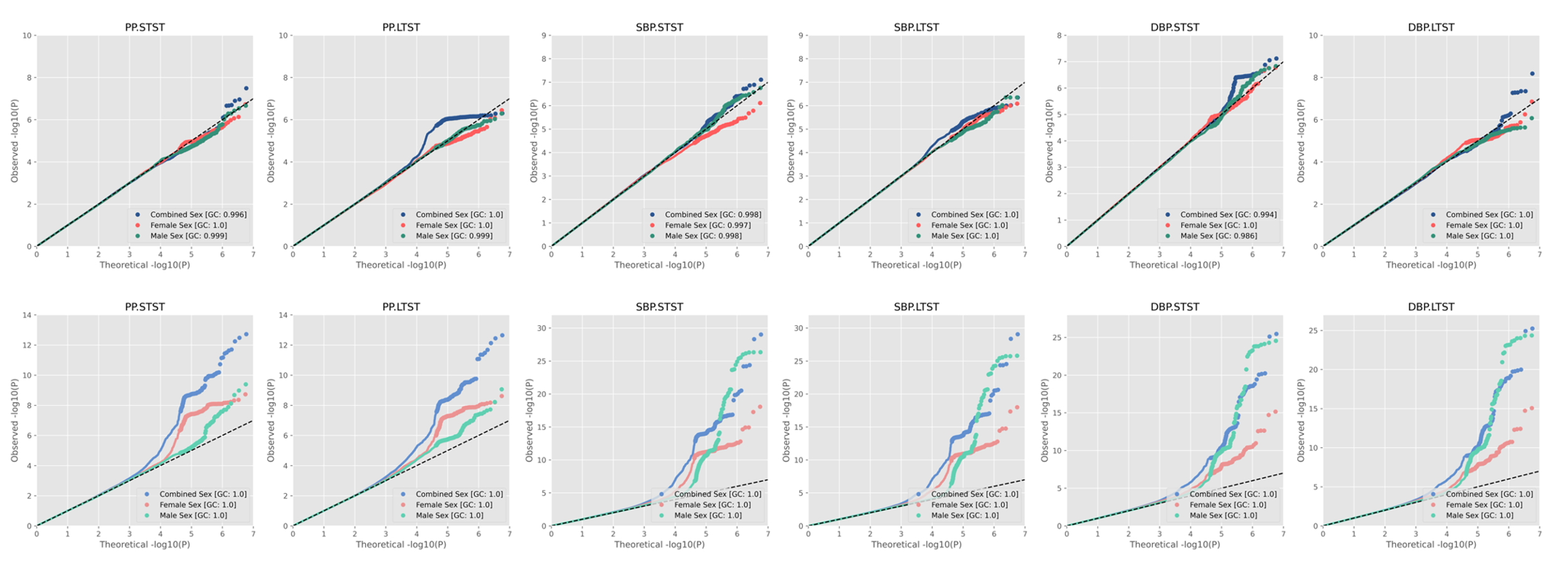

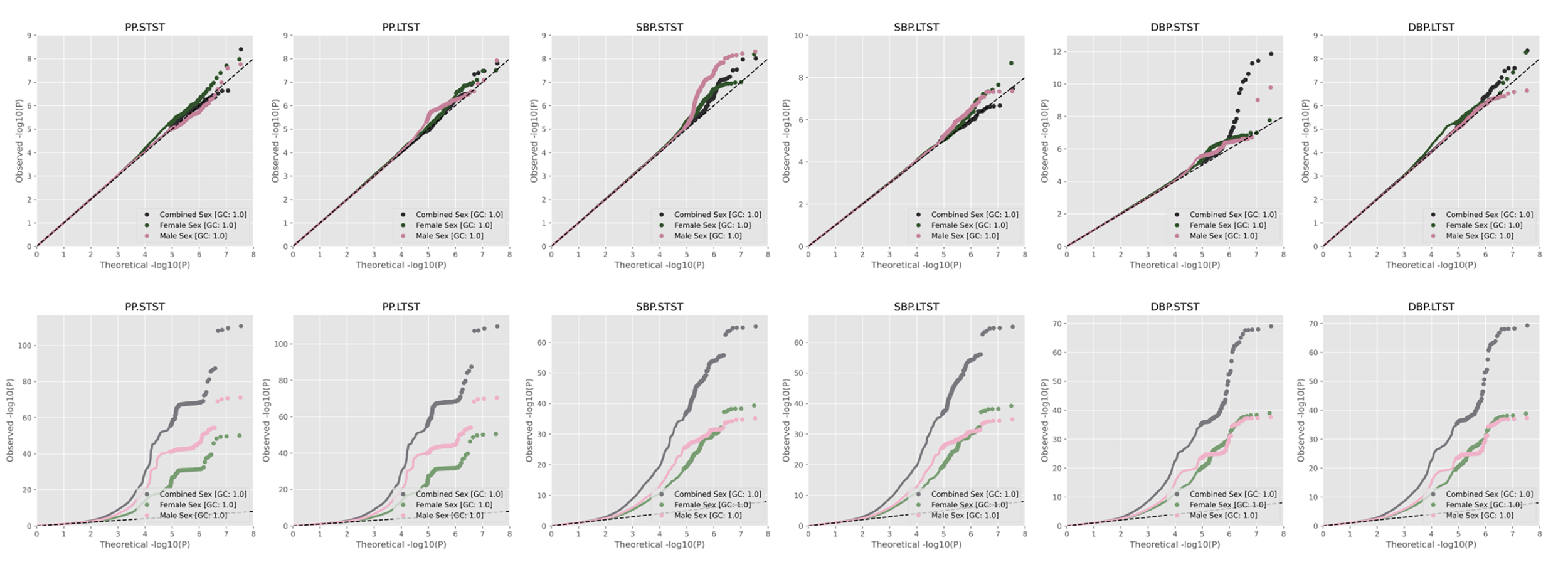
D. EUR-Specific Meta-Analysis

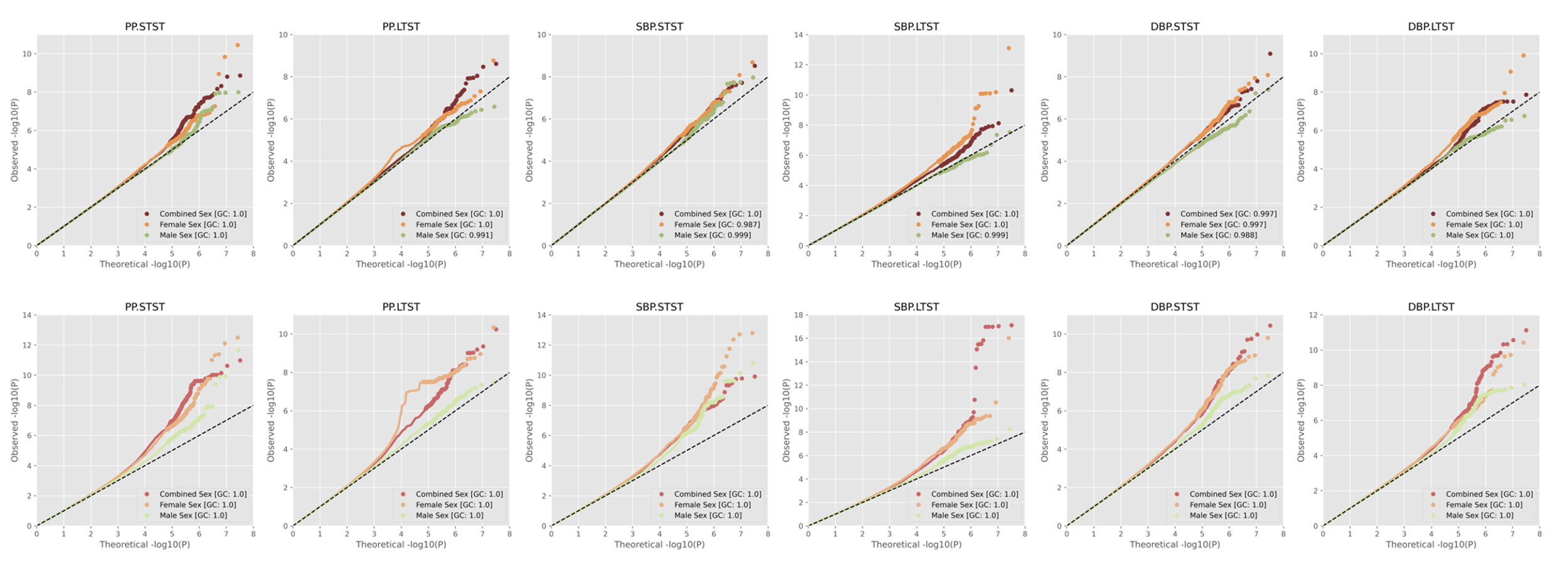
E. HIS-Specific Meta-Analysis

F. SAS-Specific Meta-Analysis

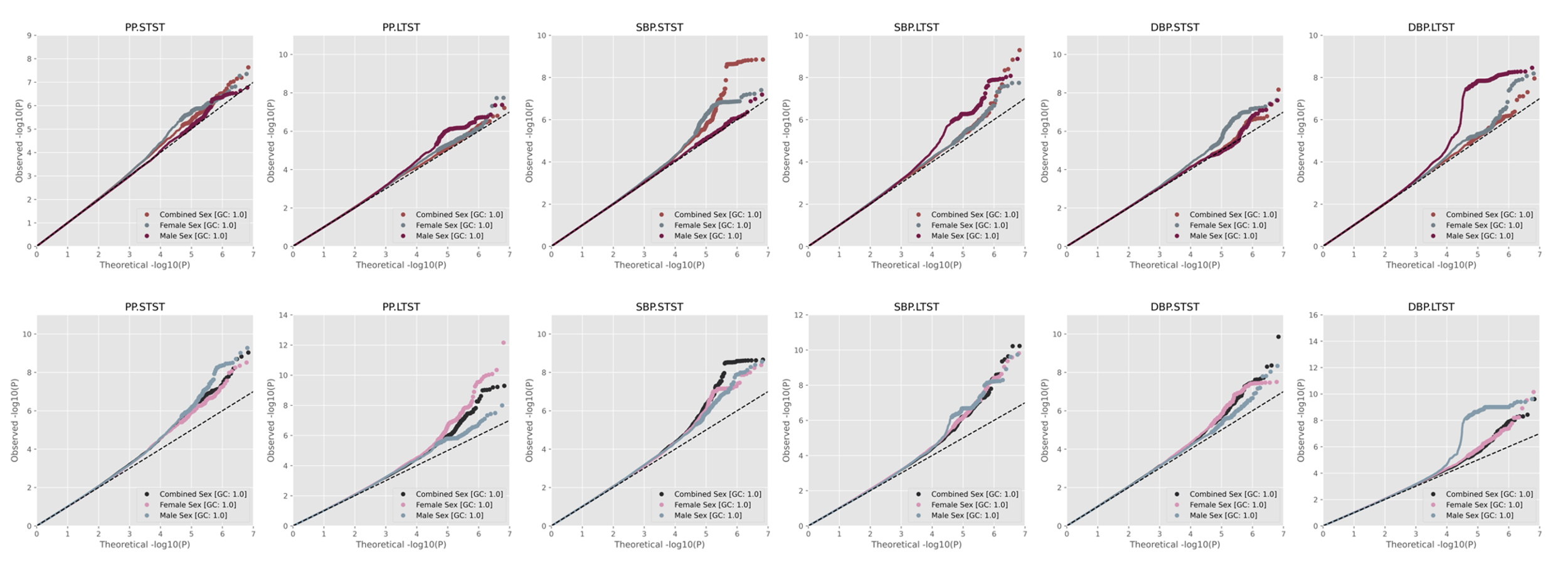

**Supplementary Figure S2. Miami Plots**

**Note:** Both lifestyle exposures (E) Long Total Sleep Time (LTST) and Short Total Sleep Time (STST) are depicted for each blood pressure trait specified below as the outcome variable. All population-group specific results are denoted - African (AFR), East Asian (EAS), European (EUR), Hispanic/Latino (HIS), and South Asian (SAS), along with cross population meta-analysis (CPMA). The top plot in each Miami plot depicts the STST interaction p-values with the bottom plot in each Miami Plot depicting the LTST interaction p-values. The left plot of each figure shows 1 degree of freedom GxE interaction effect p-values, with on the right, 2 degree of freedom joint effect p-values. For joint P-values, y-axis has a breakpoint at –log10(P)=20 to enable view, signified by the light grey dashed line. For all plots, genome-wide significance y=-log10(5e-09) is plotted as a dashed line in dark grey. Red loci signify top loci prioritized - those identified to be driven by the interaction effect, with orange loci signifying those supported by, but not driven by the interaction effect.

Regarding SNPs that passed the genome-wide threshold but did not end up being top prioritized loci, our Methods section details prioritization criteria and quality control criteria for independent top loci marked for prioritization. Briefly, all novel top loci required a SNP-specific sample size of N>20000 with respective thresholding criteria. For novel interaction loci prioritized by the 1df interaction test this was: p<5e-09, FDR<0.05. For novel interaction loci prioritized by the 2df joint test to be driven by the interaction effect this was: non-overlap with past GWAS BP loci (+/- 100 Mb distance), significant joint effect (p<5e-09, FDR<0.05), stronger interaction effect signal relative to the main genetic effect (p_M1_GxE_ < p_M1_G_), and nonsignificant marginal effect (p_M2_G_>5e-09, FDR>0.05). For secondary loci noted by the 2df joint test to be supported but not driven by the interaction effect this was: non-overlap with past GWAS BP loci (+/- 100 Mb distance), significant joint effect (p<5e-09, FDR<0.05), weaker interaction effect signal relative to the main genetic effect (p_M1_GxE_ > p_M1_G_), and nonsignificant marginal effect (p_M2_G_>5e-09, FDR>0.05). Variants were filtered to independent genomic loci (across 500 kb regions) using LD reference panels, with missing variants not found in panels to be reported if the variant was found to be the top most significant in a respective 500kb region in combined sex analyses.

**A. Miami Plots for Pulse Pressure in Combined Sex**

1. Cross Population Meta-Analysis

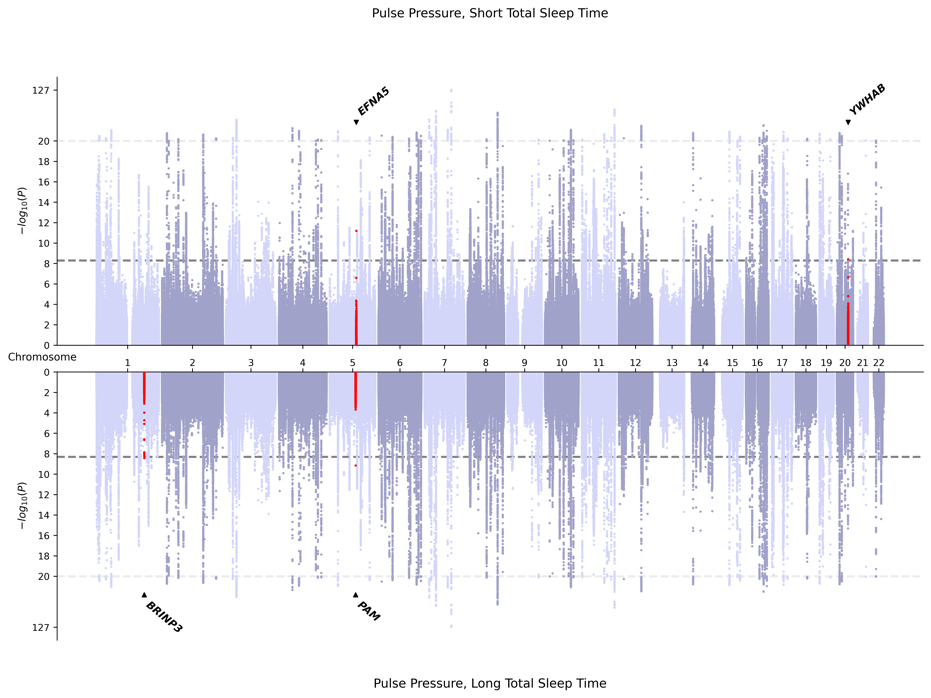

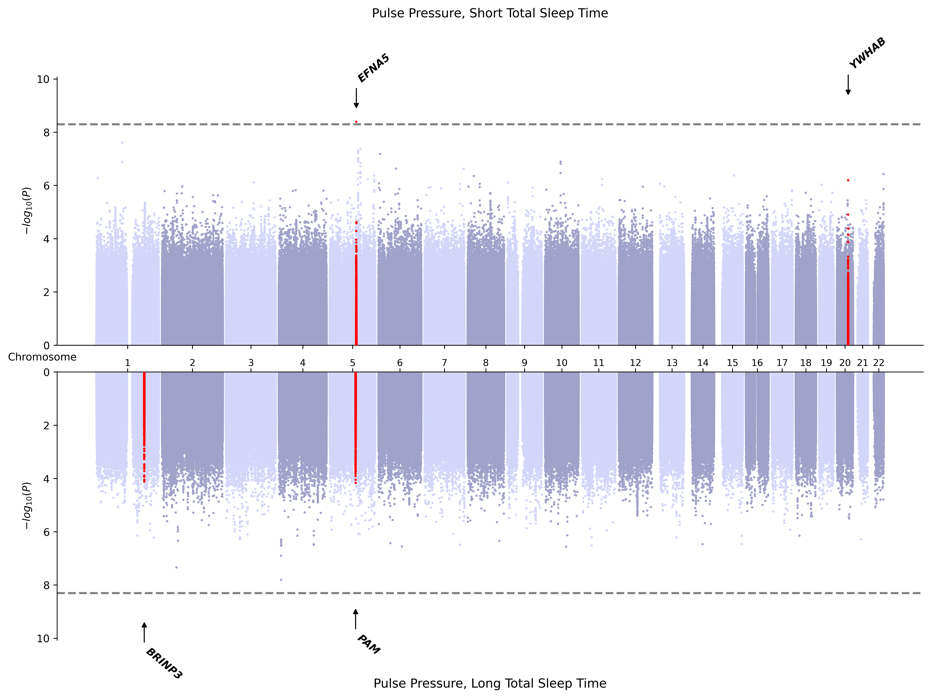

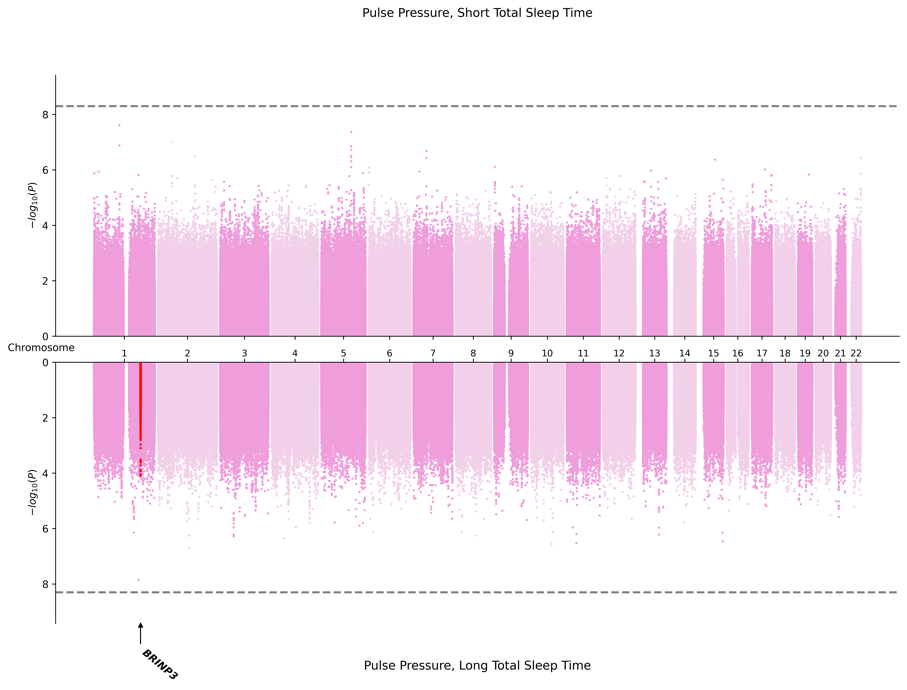
2. AFR-Specific Meta-Analysis

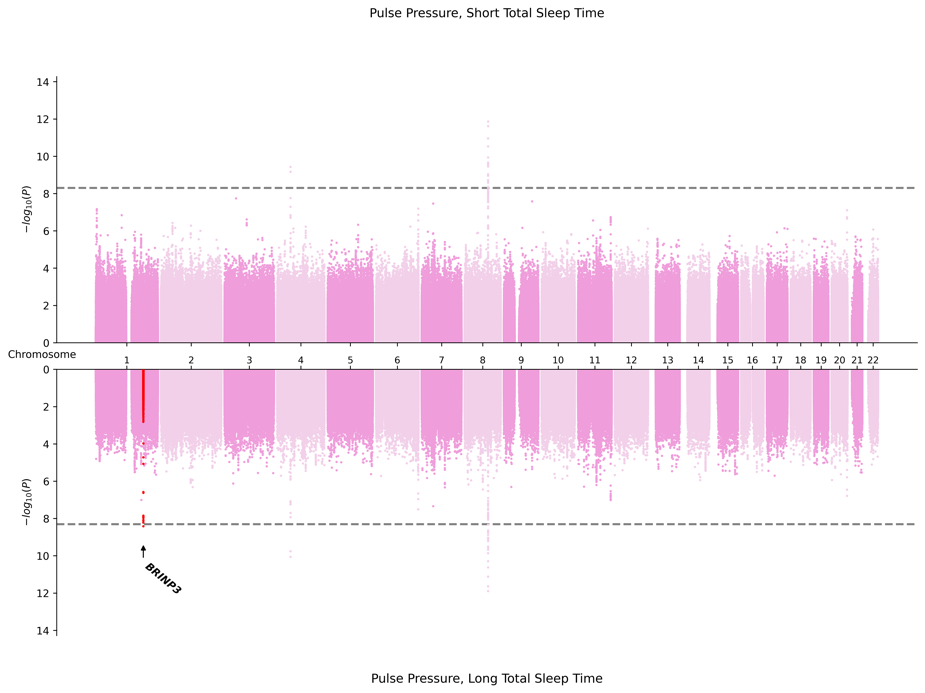

3. EAS-Specific Meta-Analysis

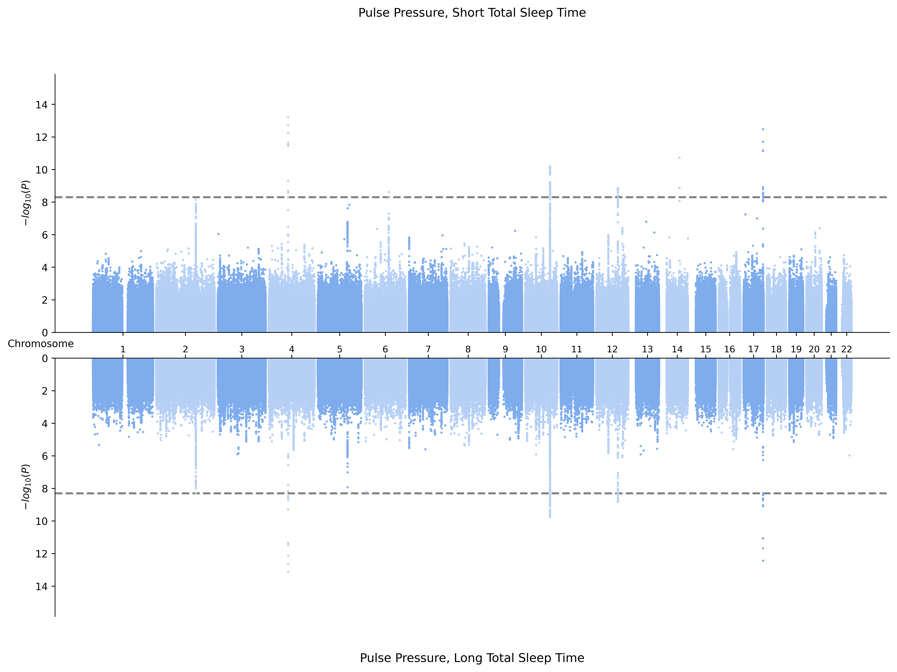

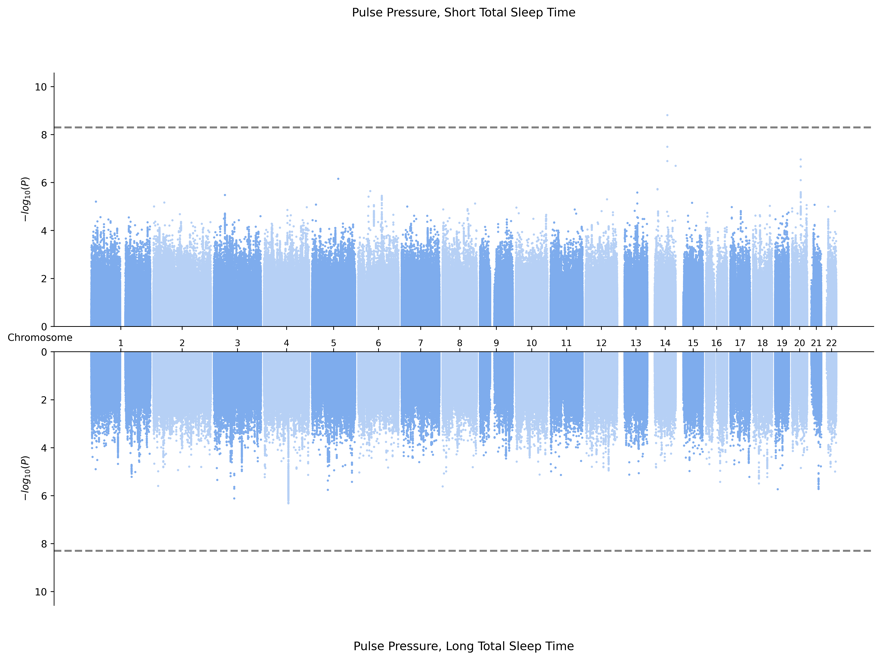

4. EUR-Specific Meta-Analysis
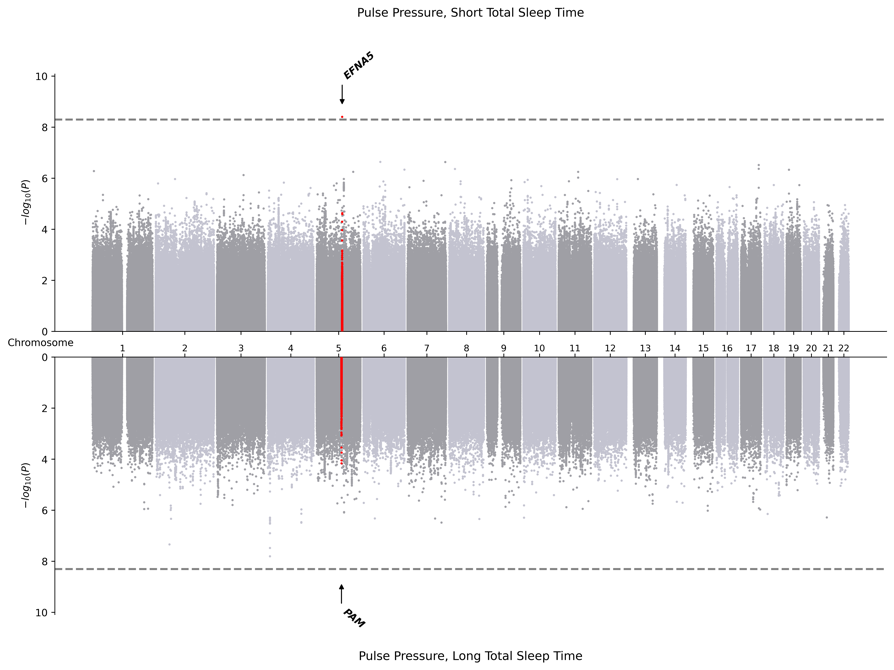

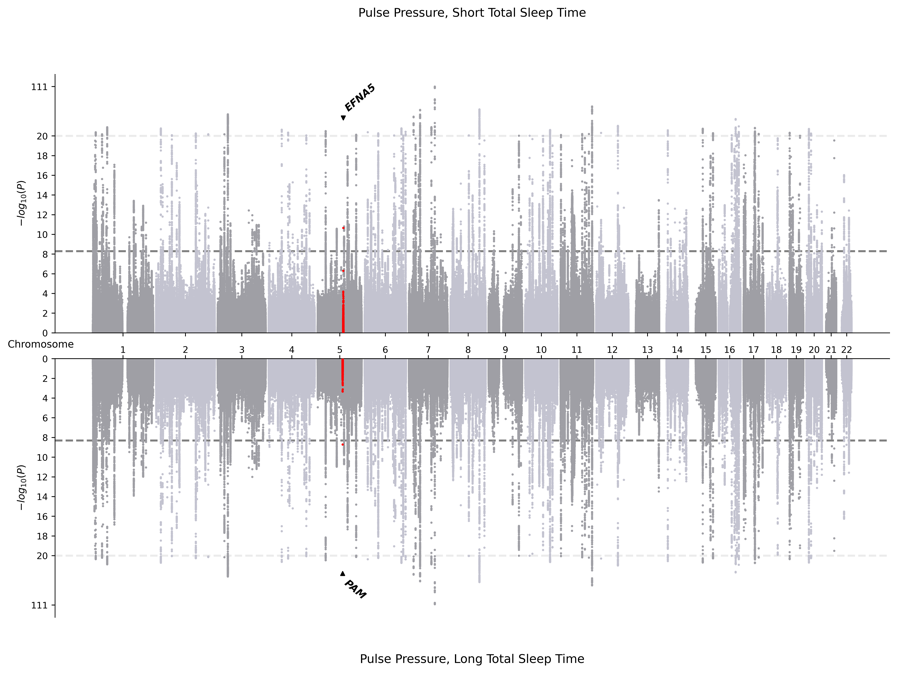

5. HIS-Specific Meta-Analysis
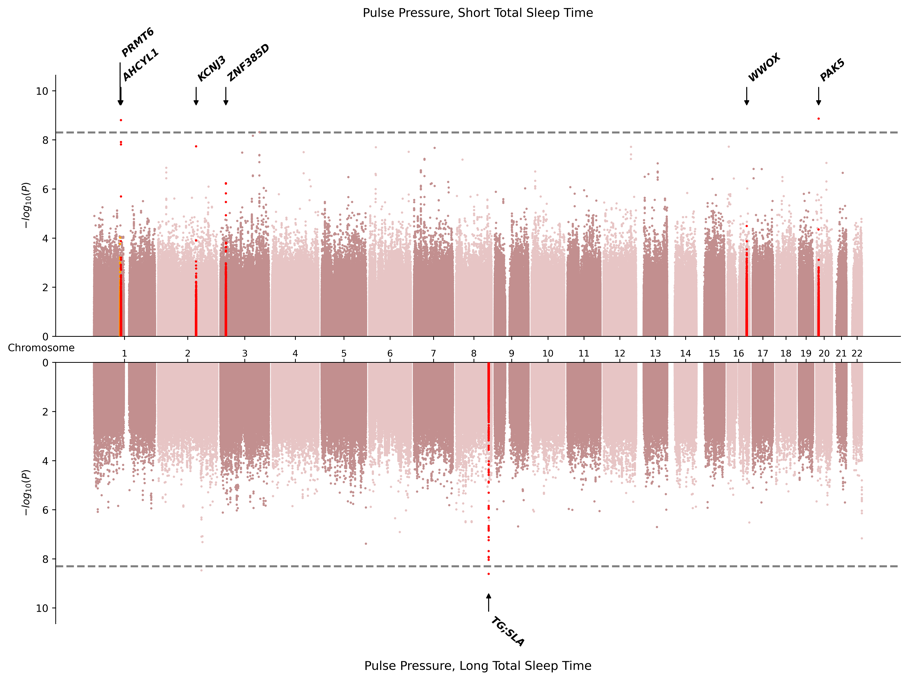

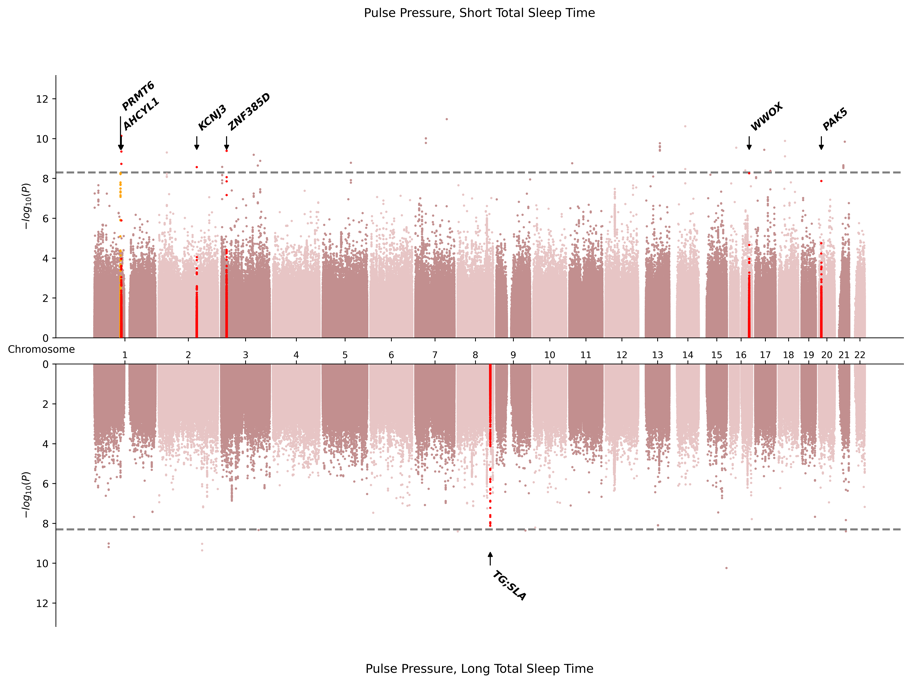

6. SAS-Specific Meta-Analysis

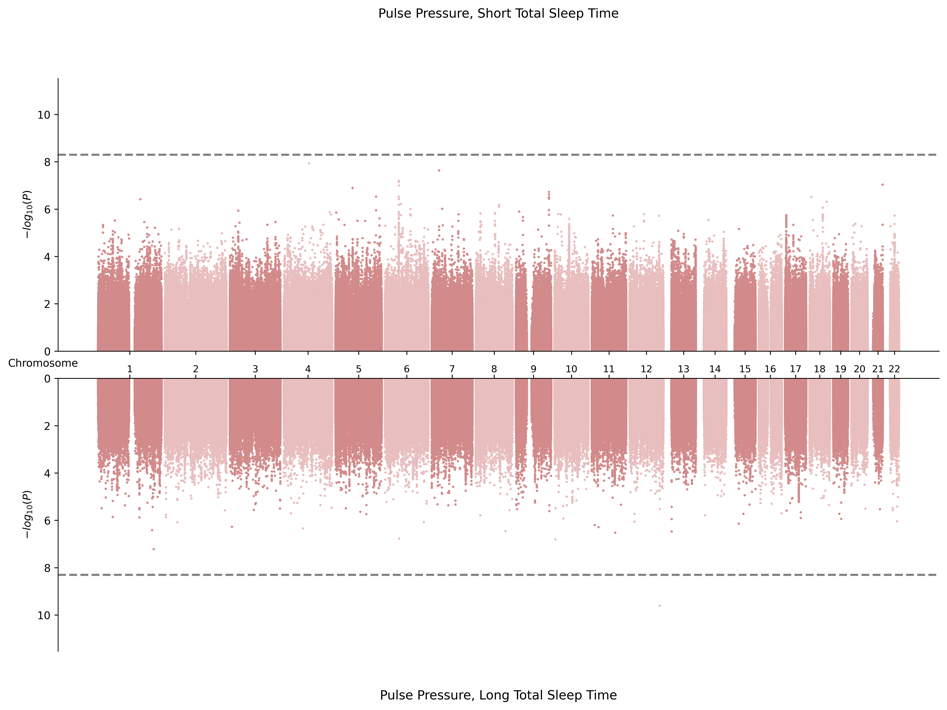

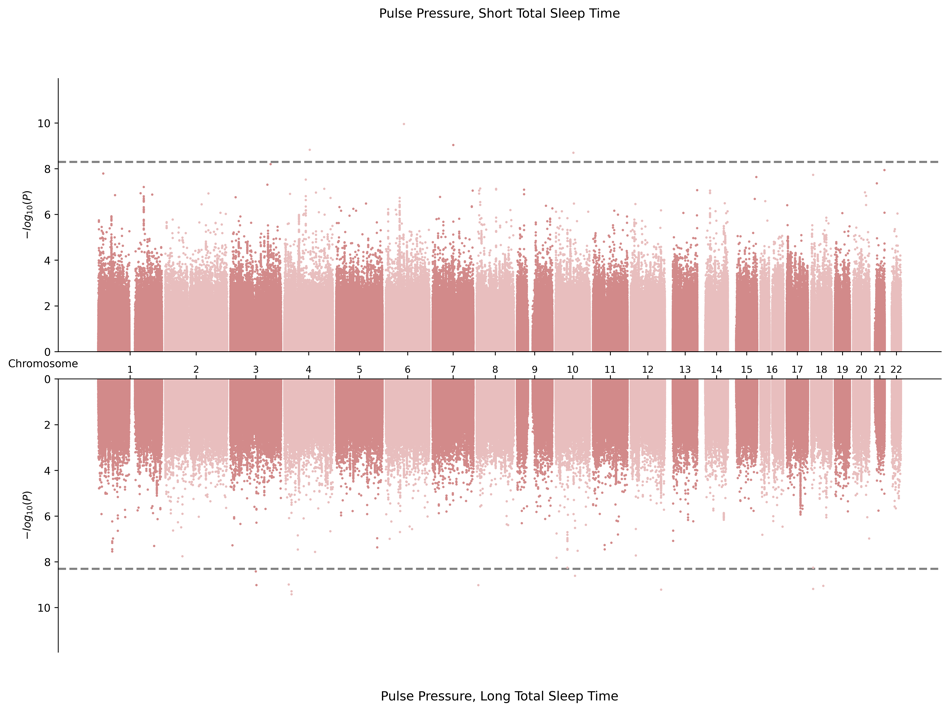

**B.** **Miami Plots for Systolic Blood Pressure in Combined Sex**

1. Cross Population Meta-Analysis

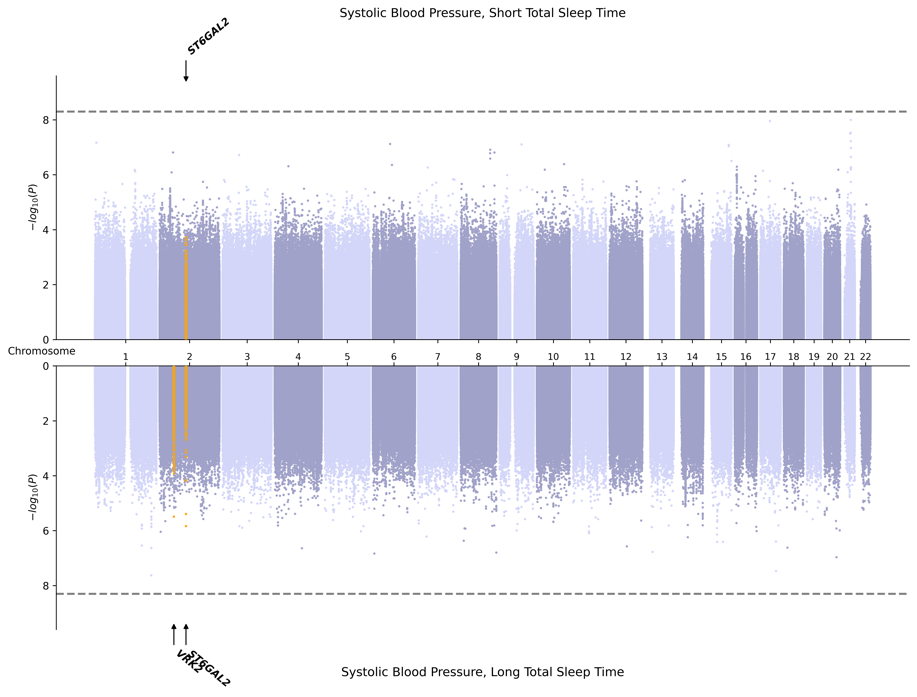

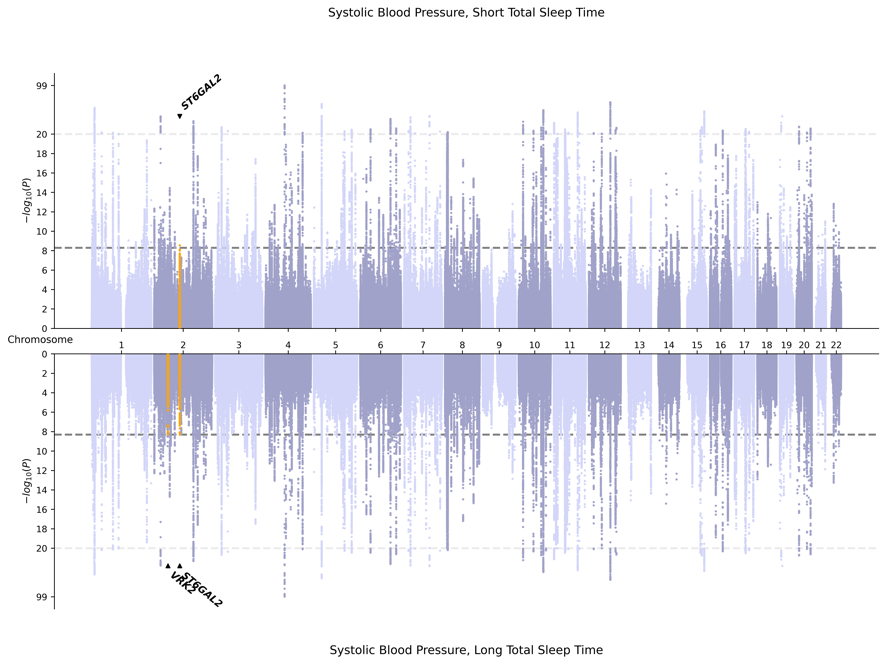

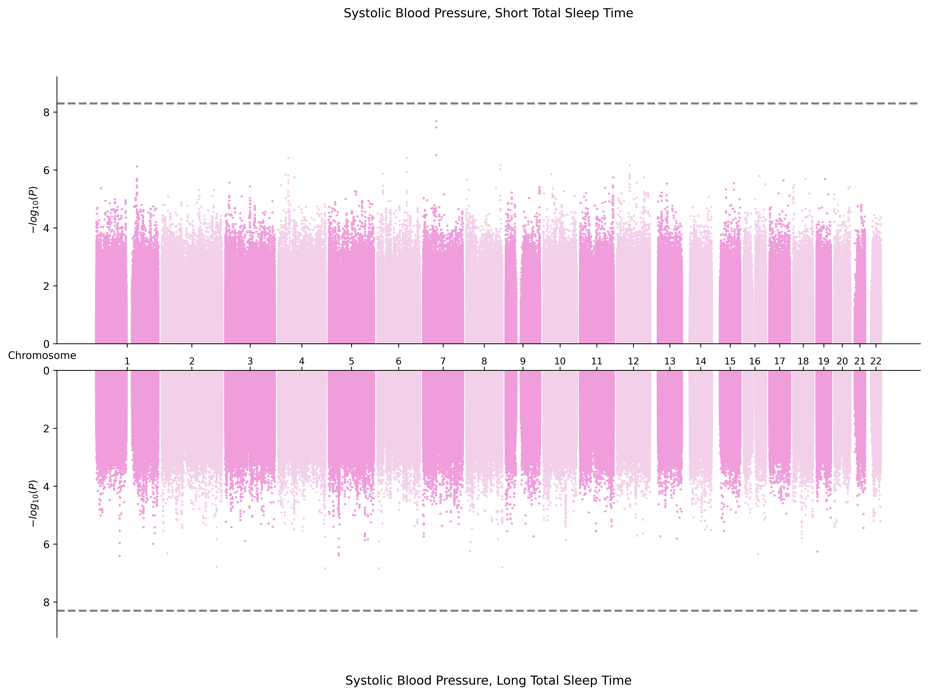

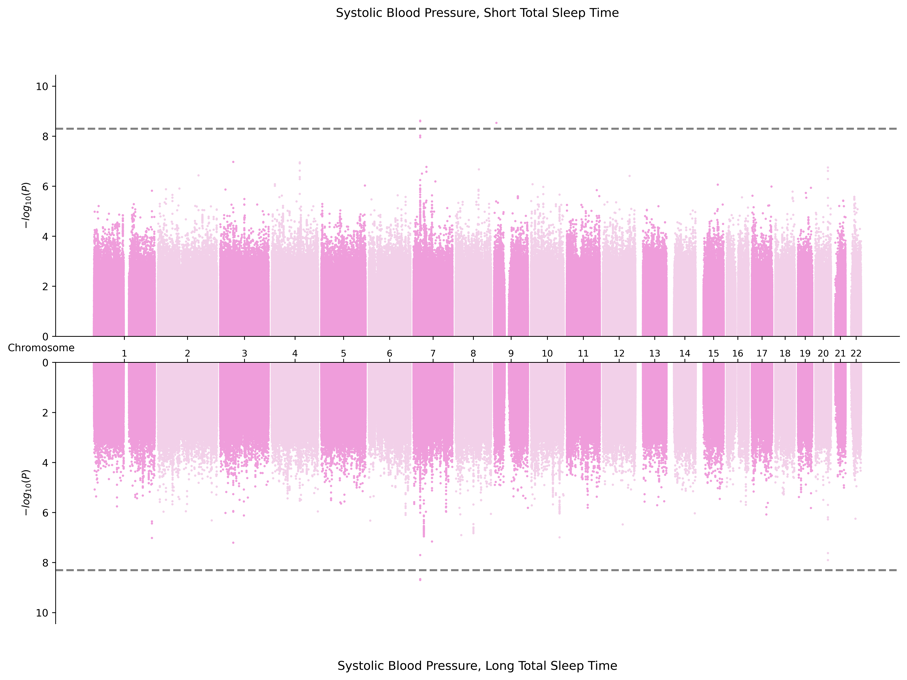
2. AFR-Specific Meta-Analysis

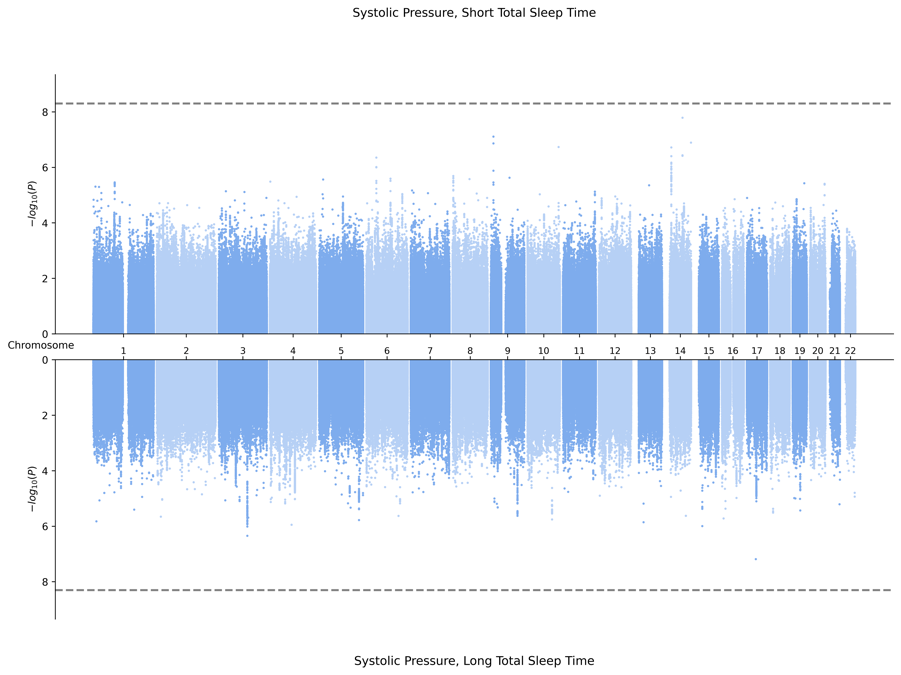

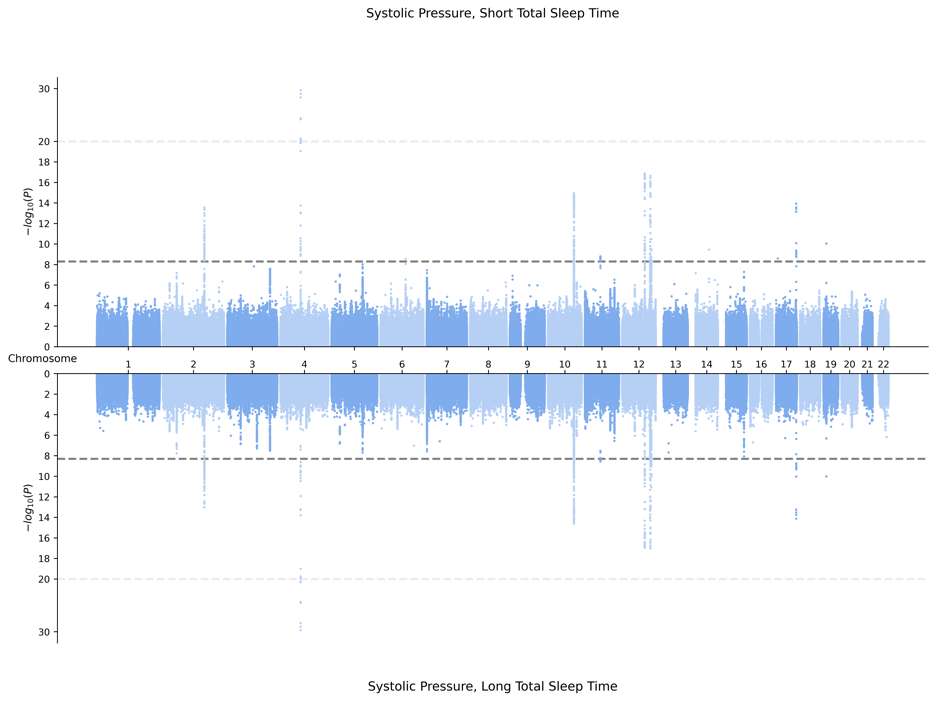
3. EAS-Specific Meta-Analysis

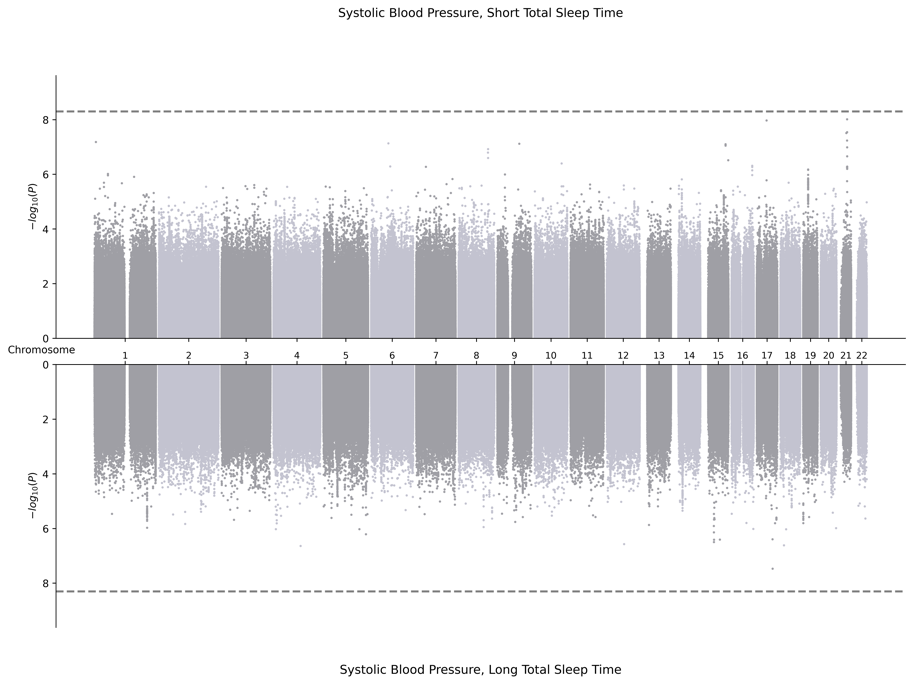
4. EUR-Specific Meta-Analysis

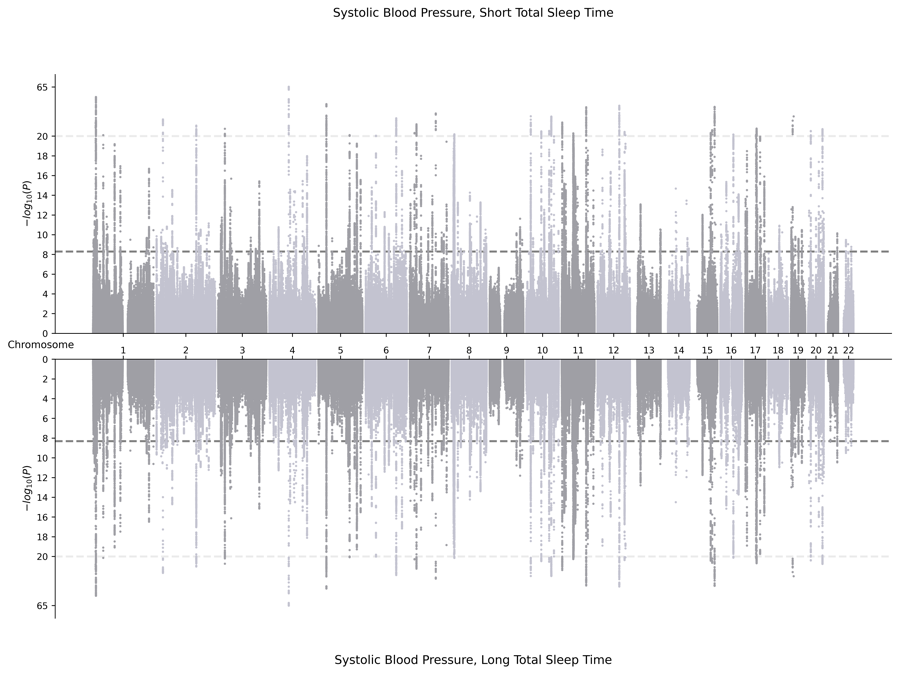

5. HIS-Specific Meta-Analysis

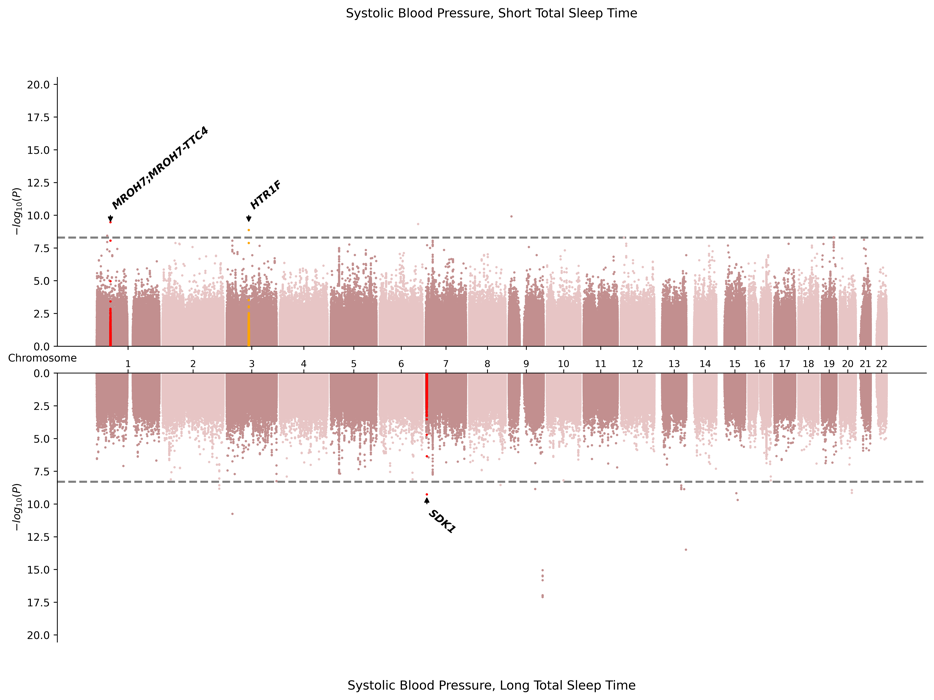

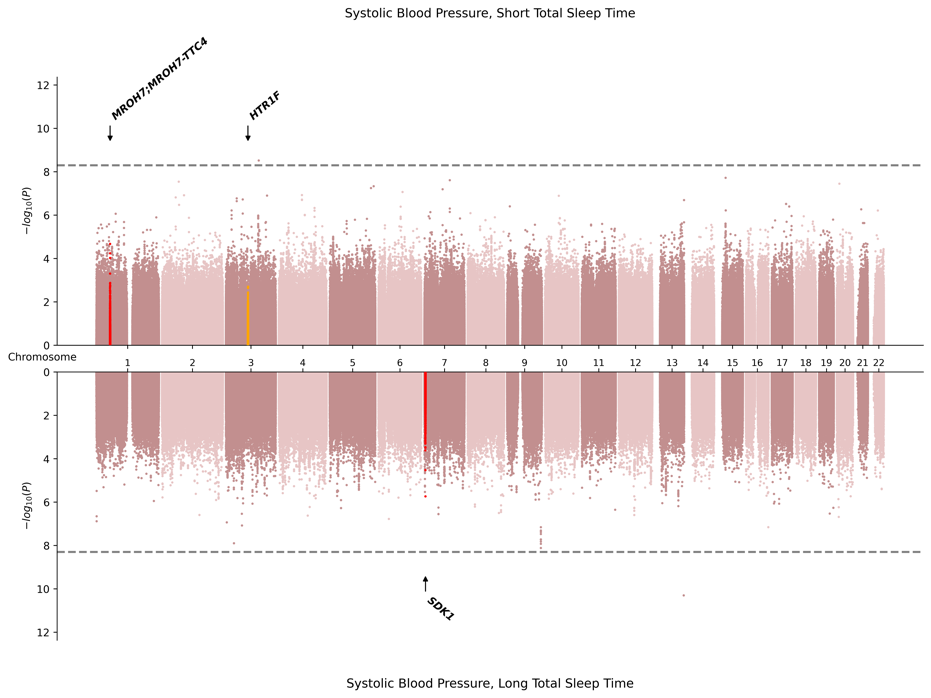

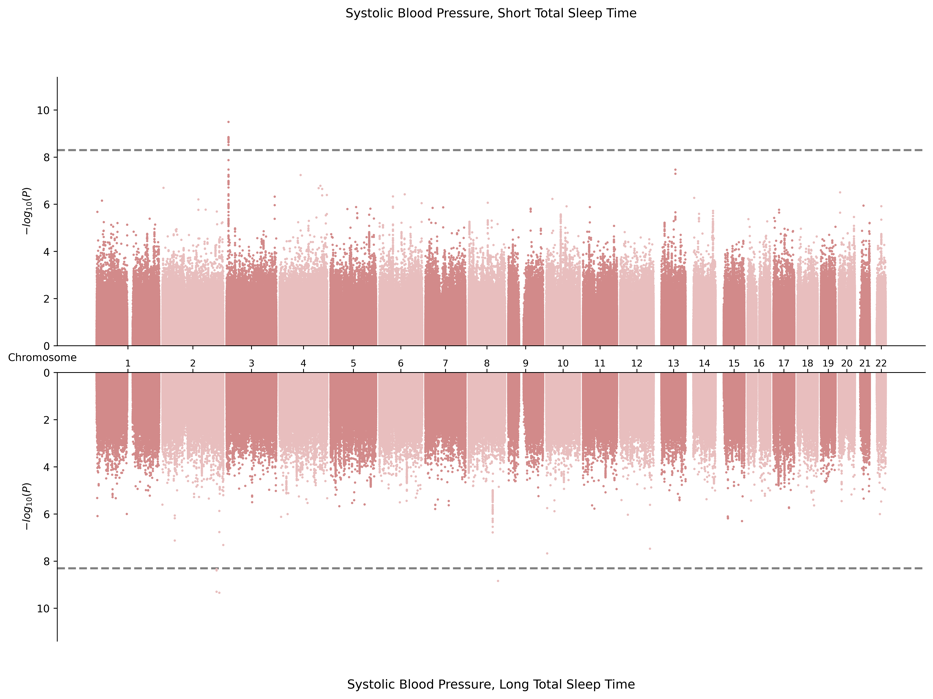
6. SAS-Specific Meta-Analysis

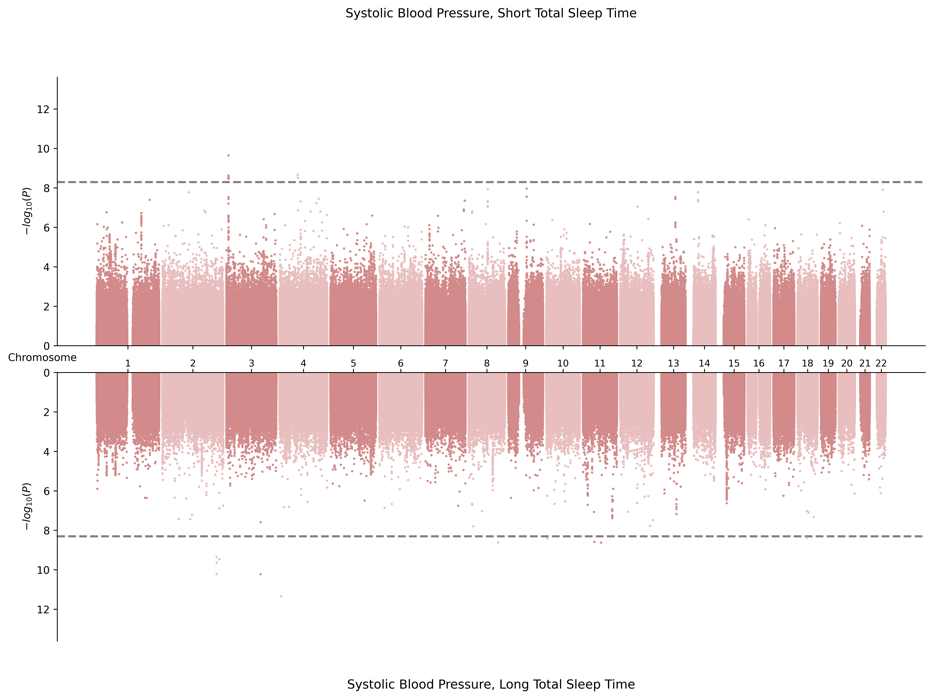

**C.** **Miami Plots for Diastolic Pressure in Combined Sex**

1. Cross Population Meta-Analysis

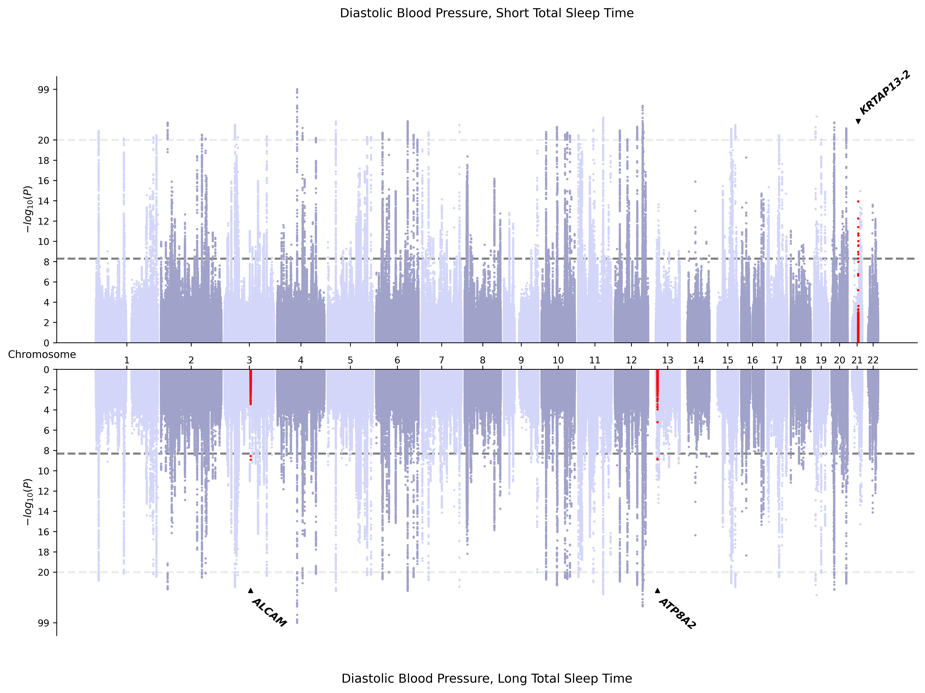

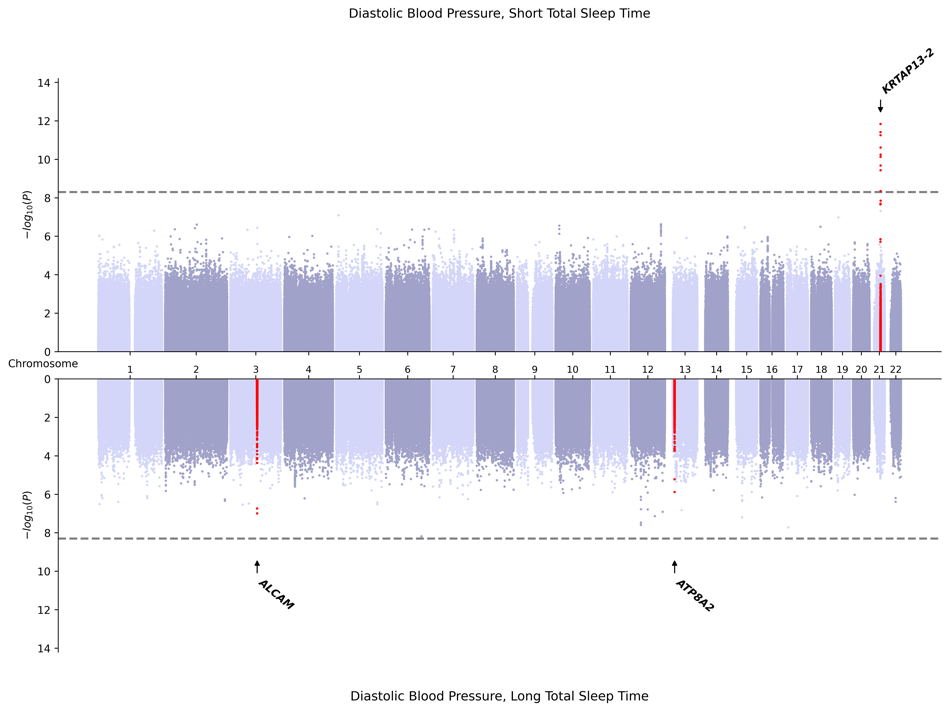

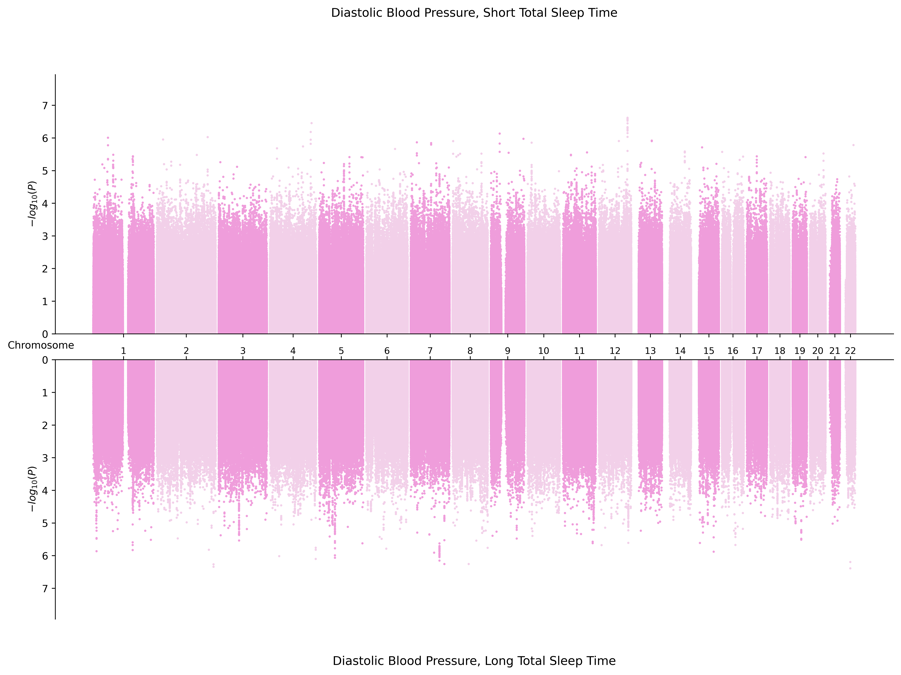
2. AFR-Specific Meta-Analysis

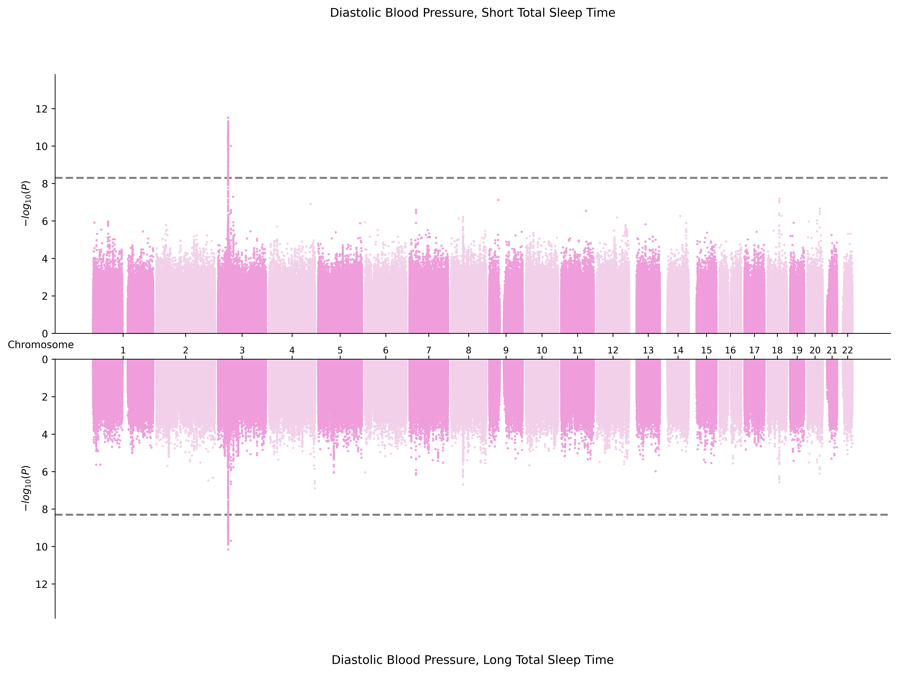

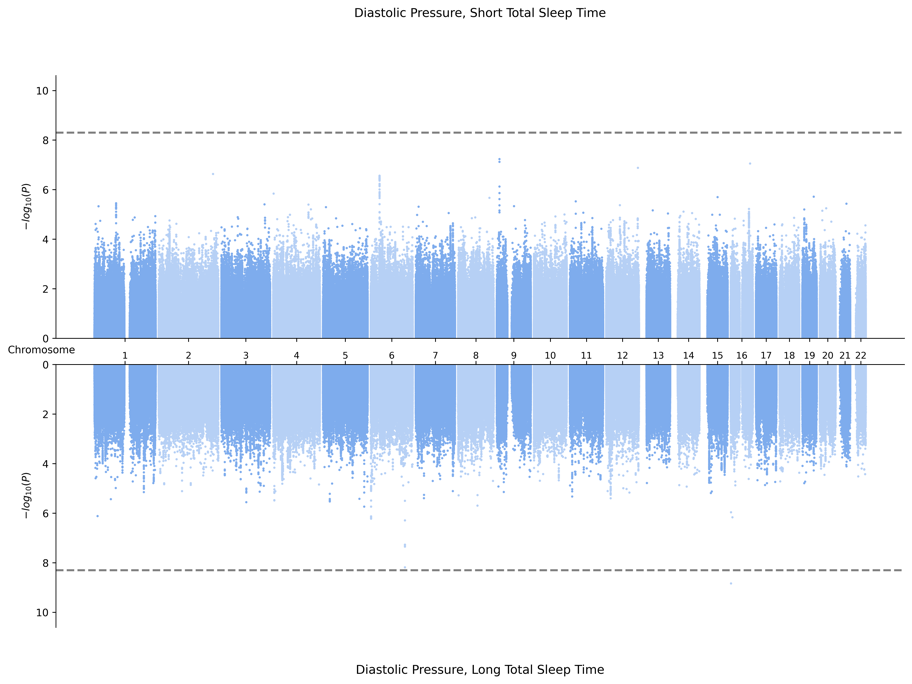
3. EAS-Specific Meta-Analysis

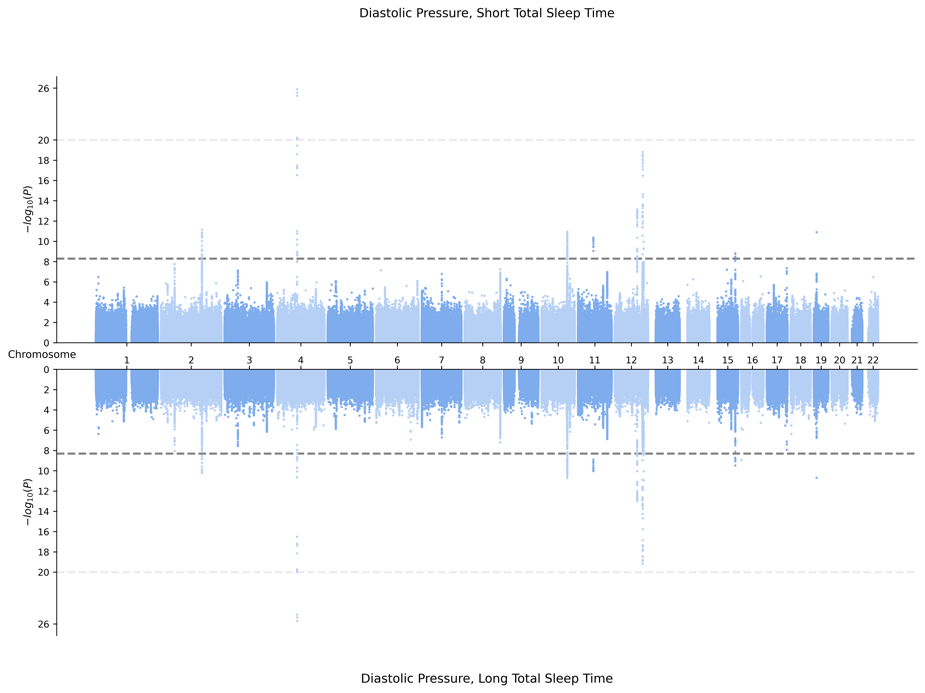

4. EUR-Specific Meta-Analysis

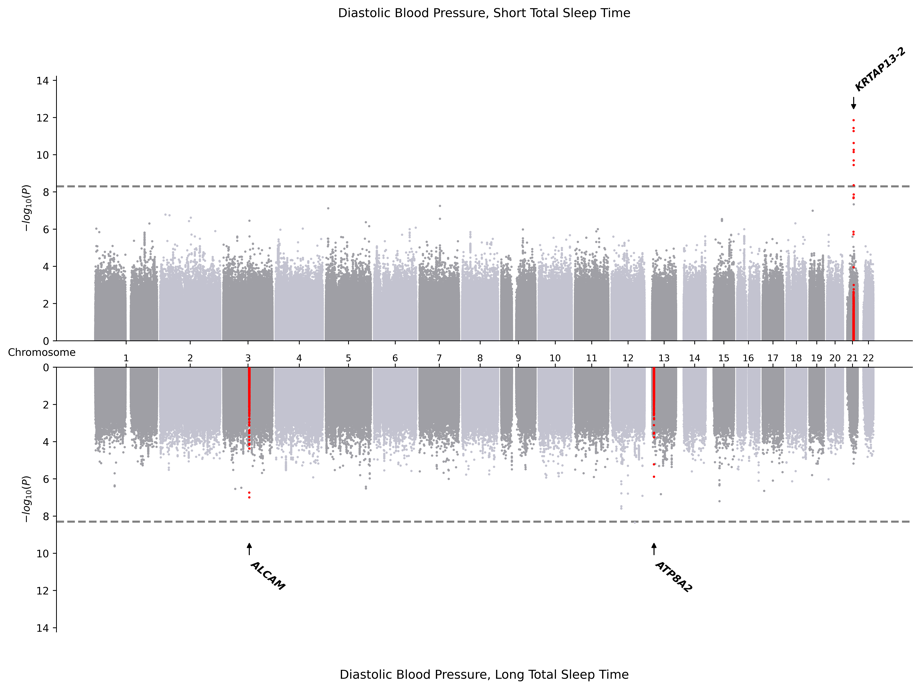

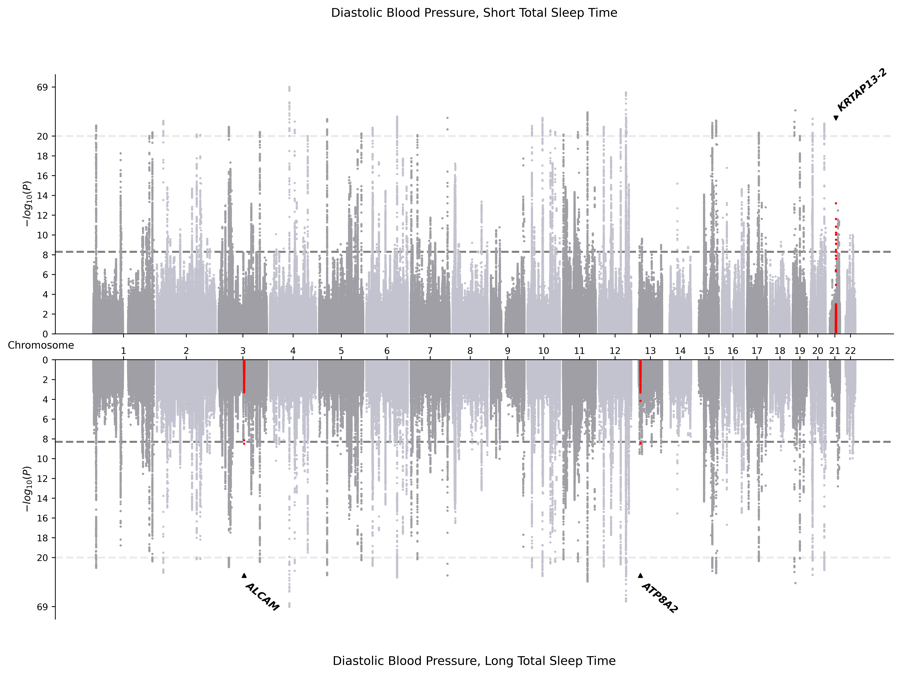

5. HIS-Specific Meta-Analysis

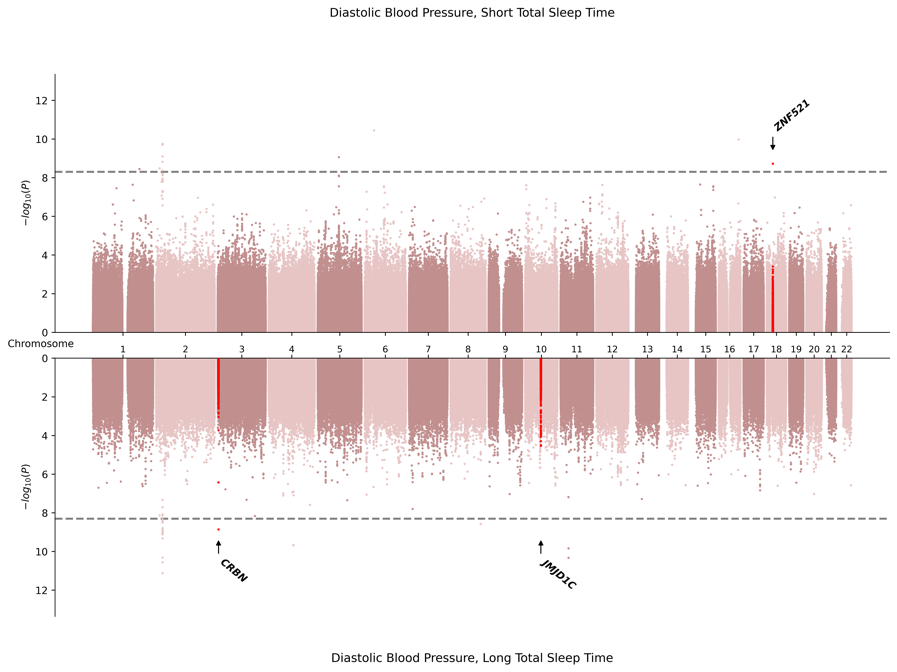

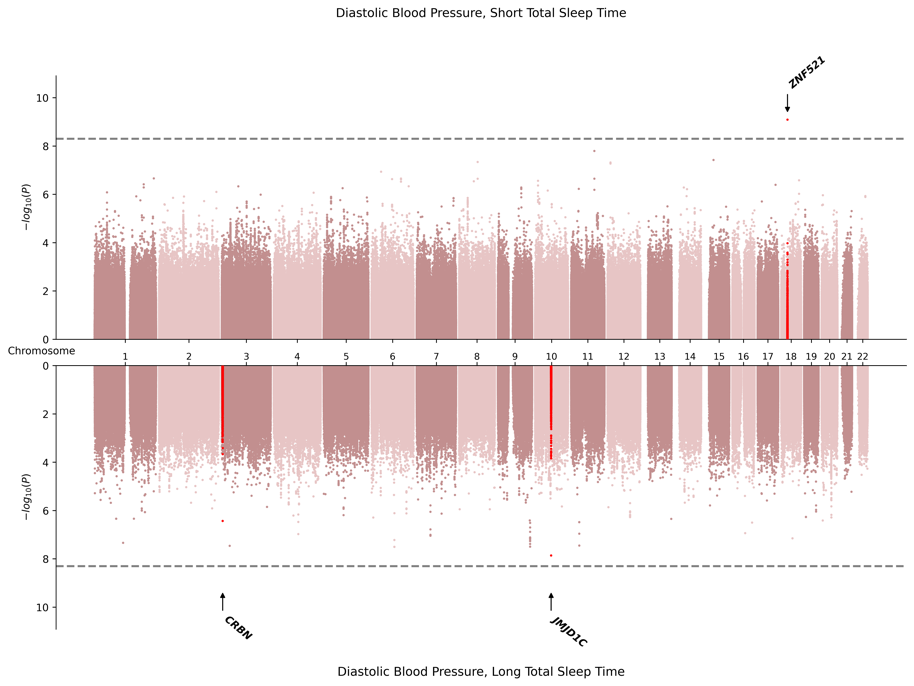

6. SAS-Specific Meta-Analysis

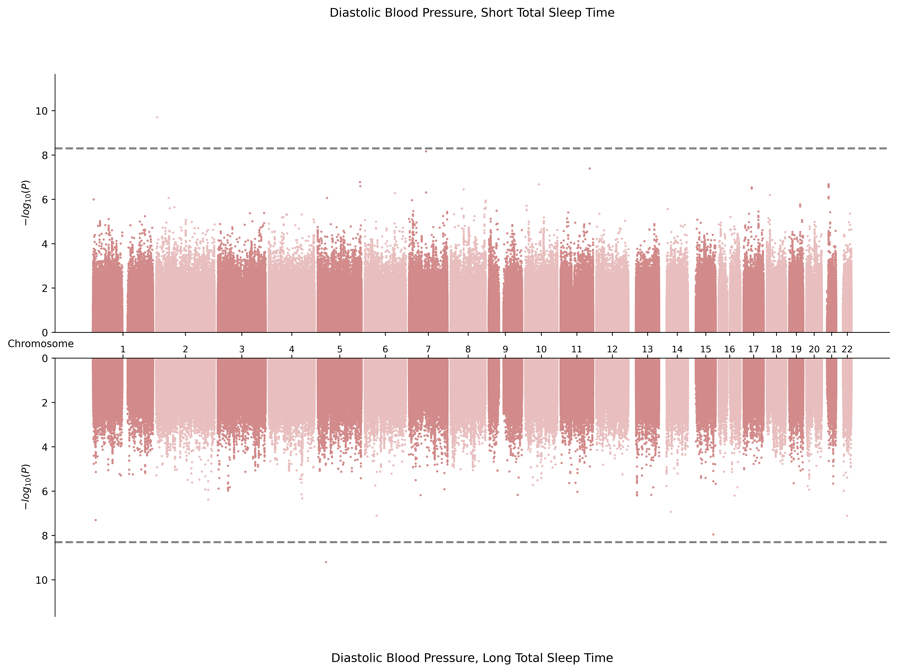

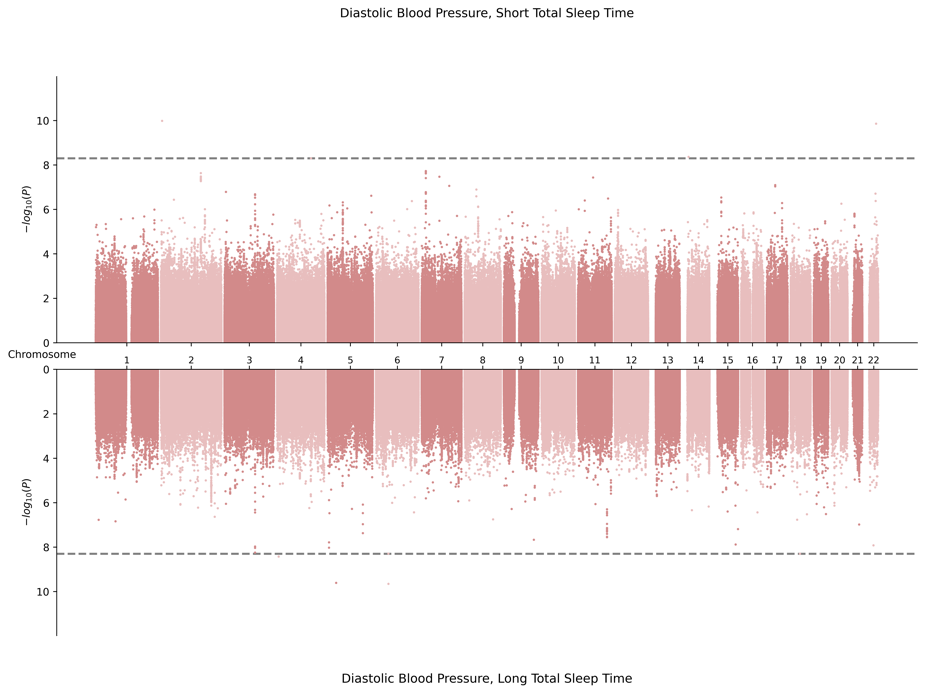

**D.** **Miami Plots for Pulse Pressure in Female Sex**

1. Cross Population-Meta Analysis

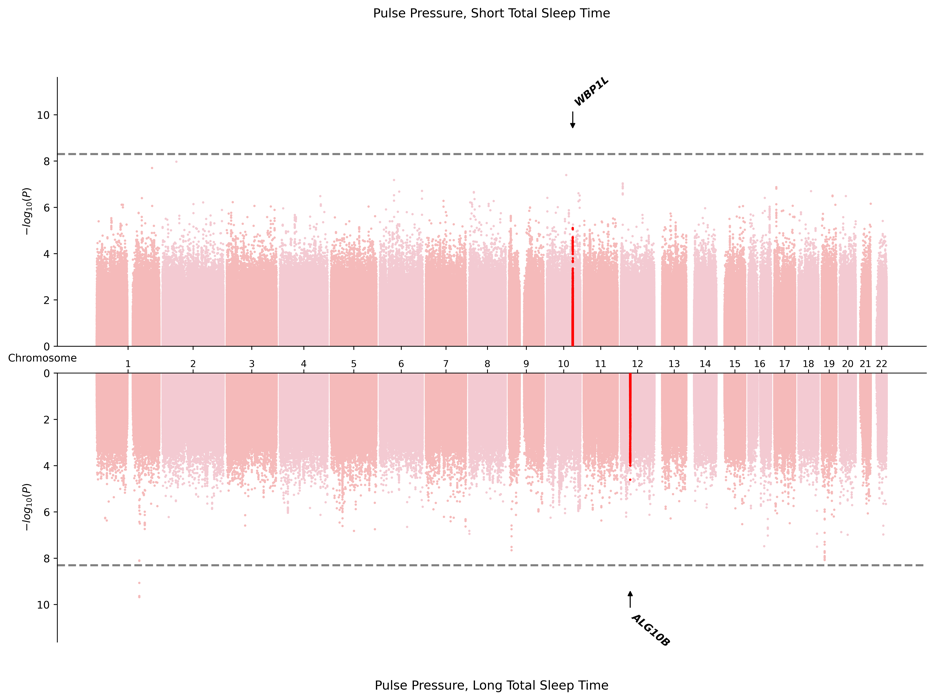

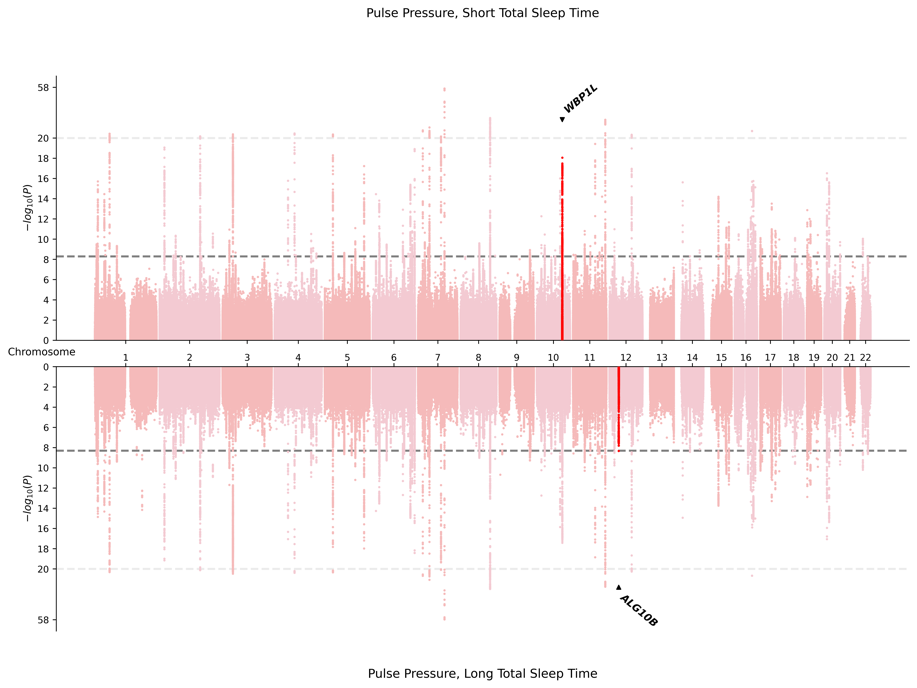

2. AFR-Specific Meta-Analysis

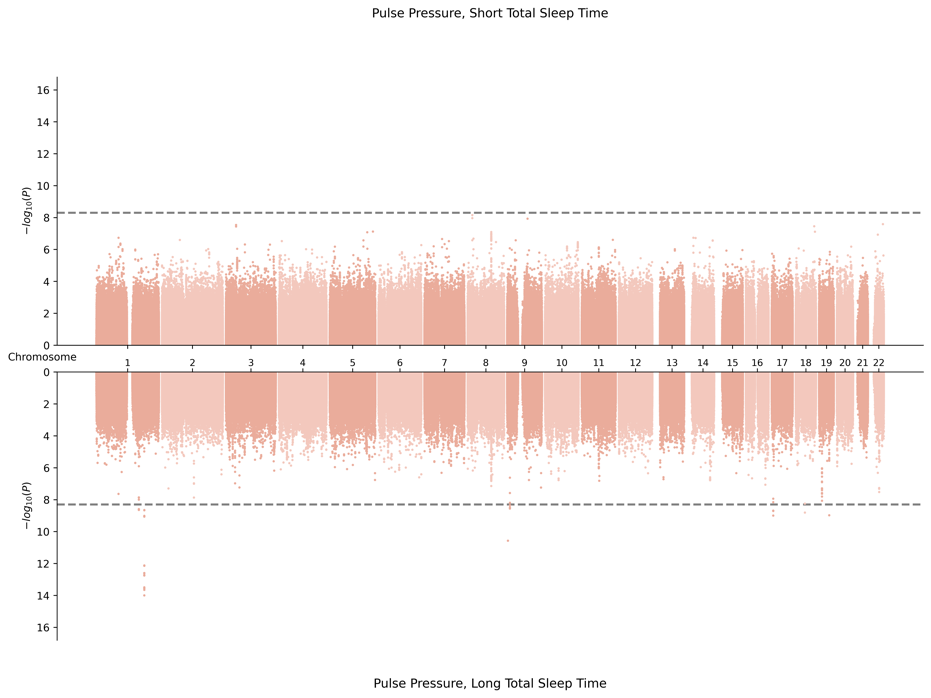

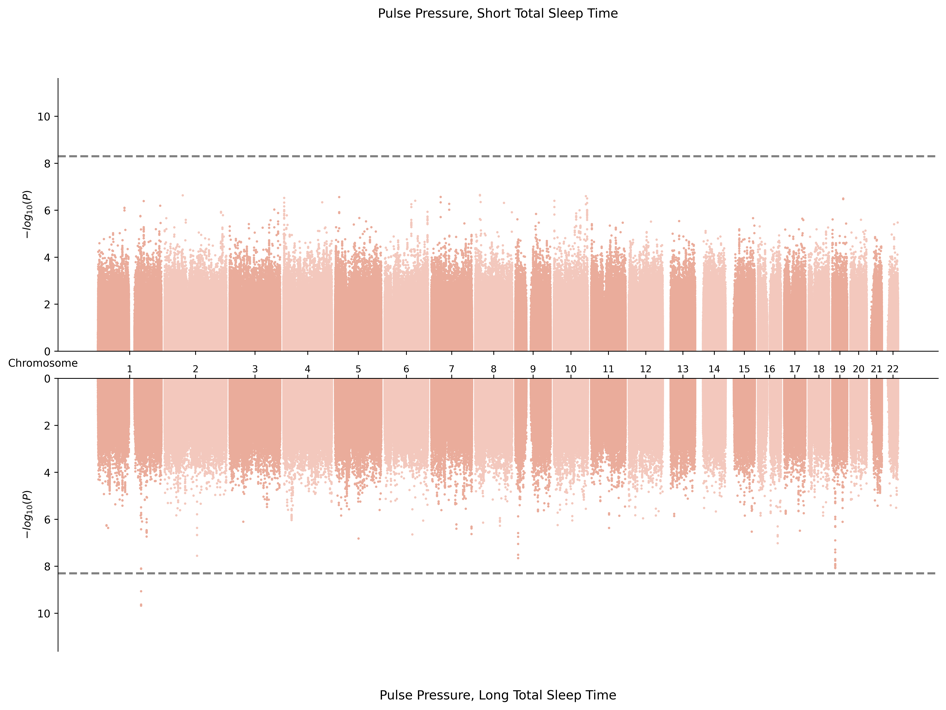

3. EAS-Specific Meta-Analysis

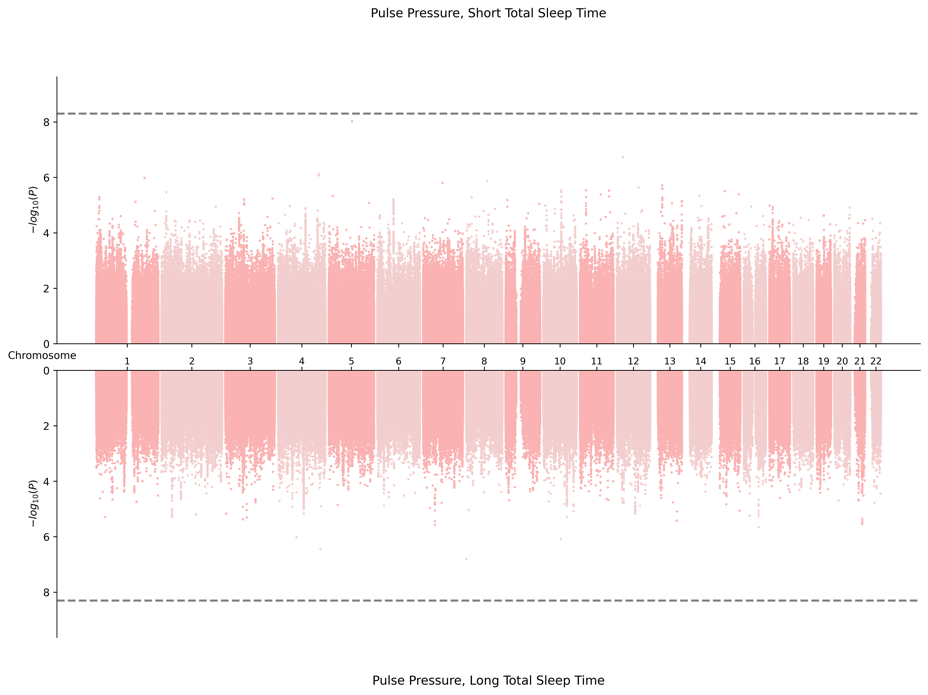

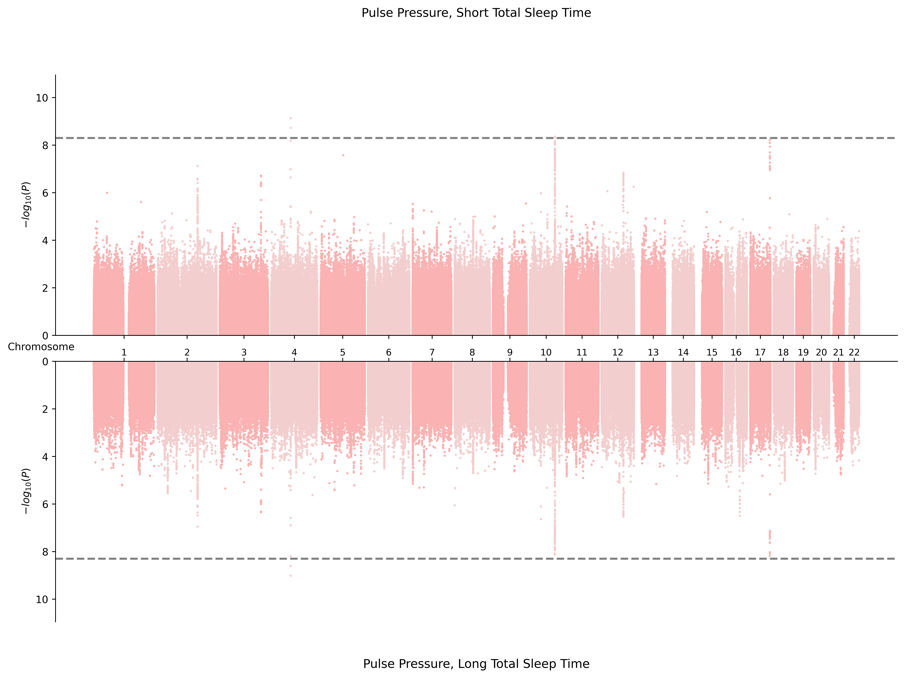

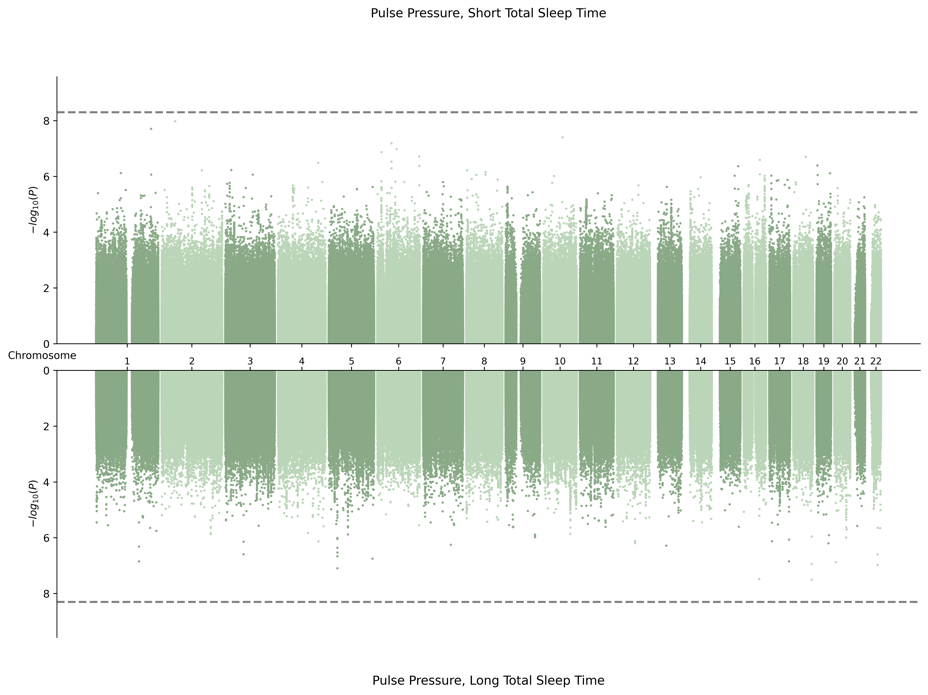
4. EUR-Specific Meta-Analysis

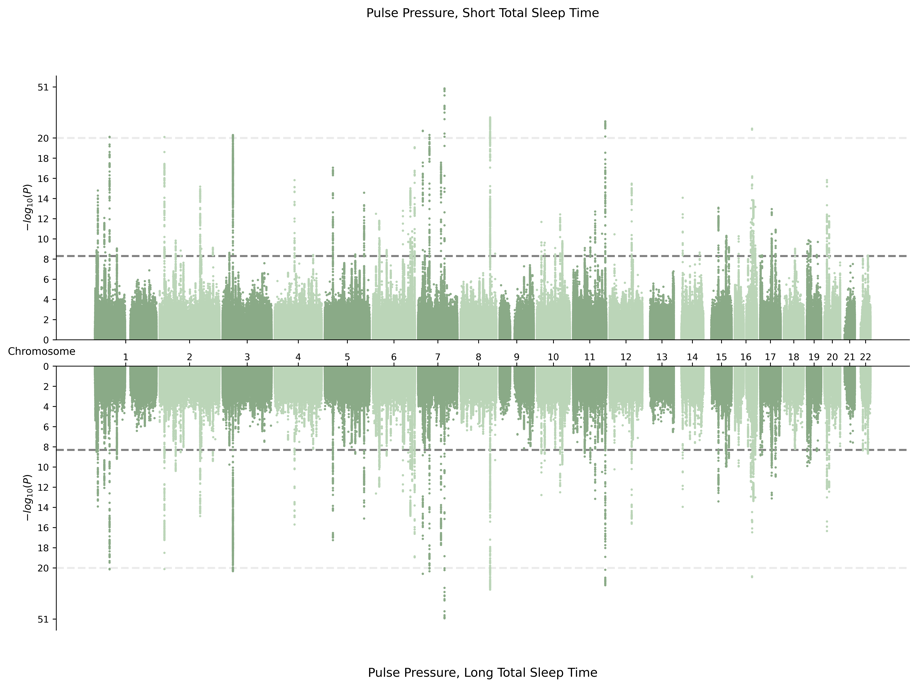

5. HIS-Specific Meta-Analysis

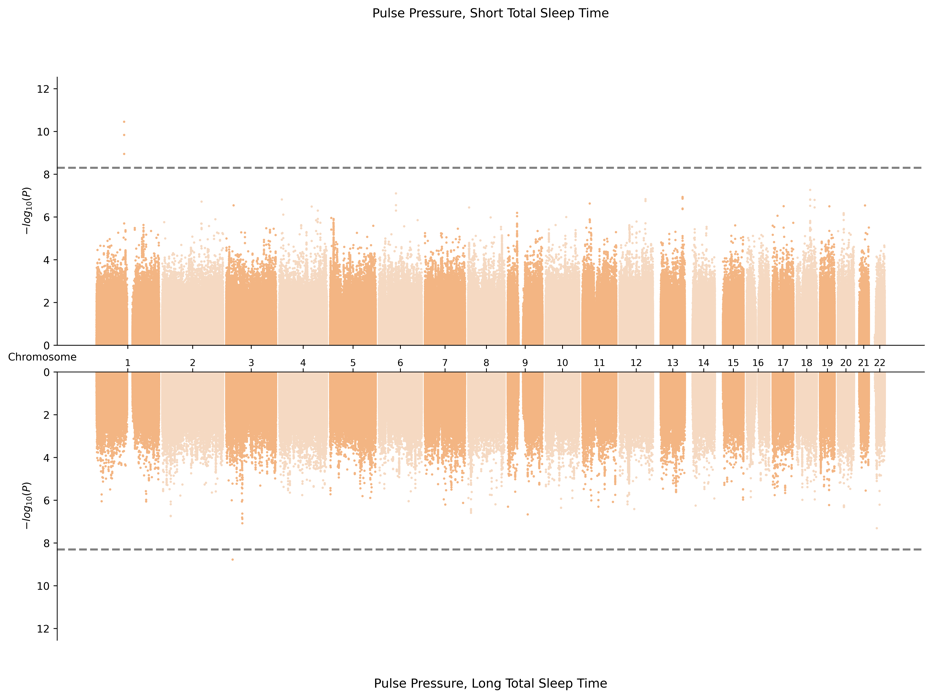

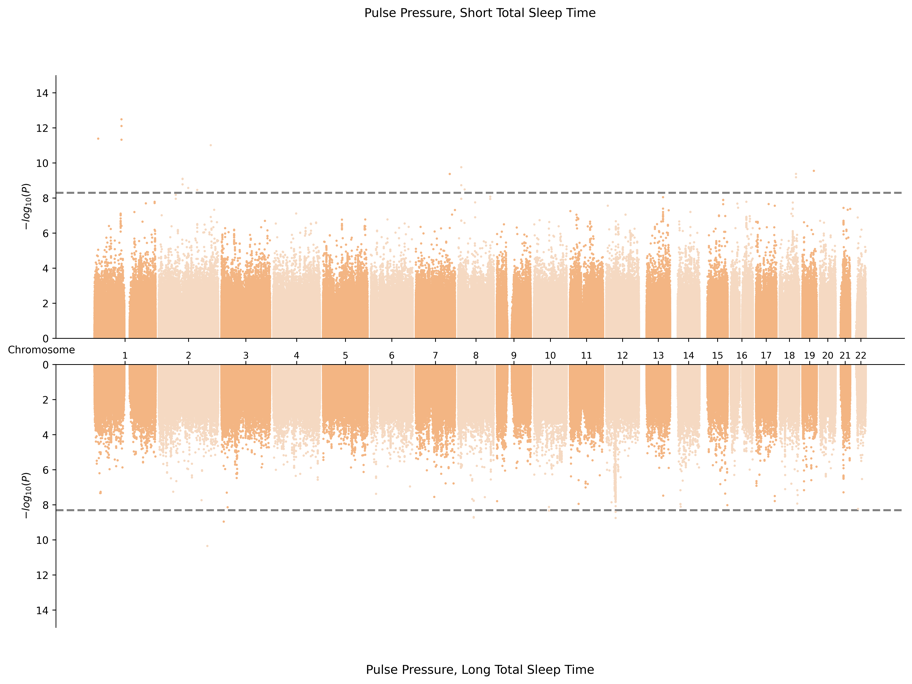

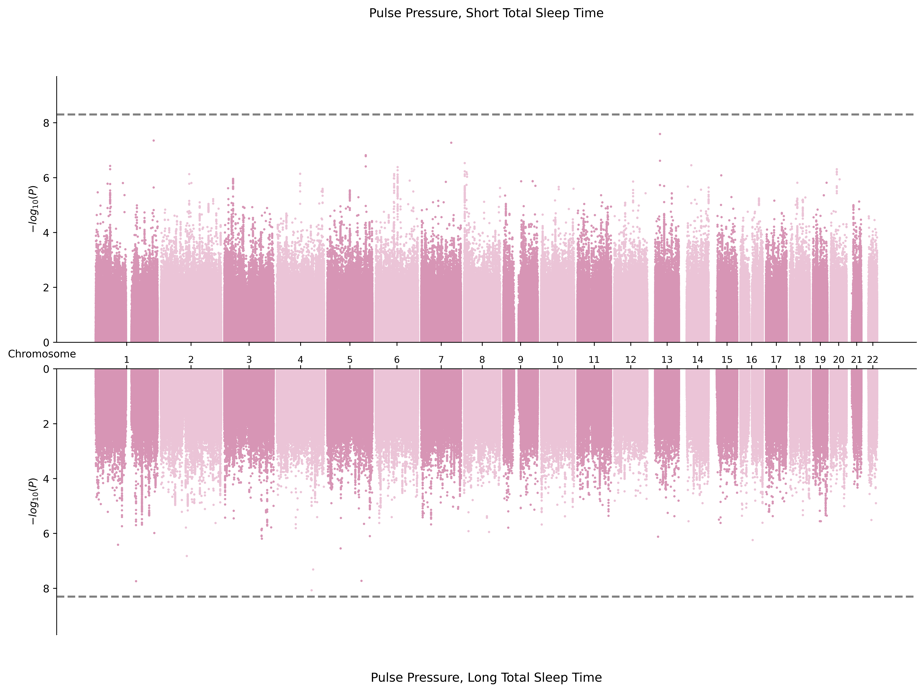
6. SAS-Specific Meta-Analysis

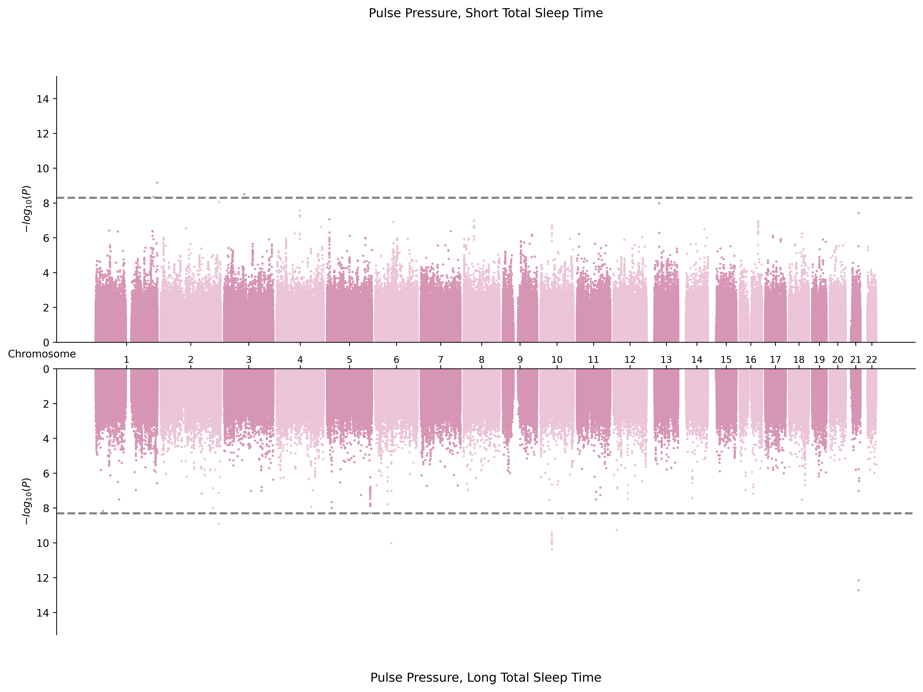

**E.** **Miami Plots for Systolic Blood Pressure in Female Sex**

1. Cross Population Meta-Analysis

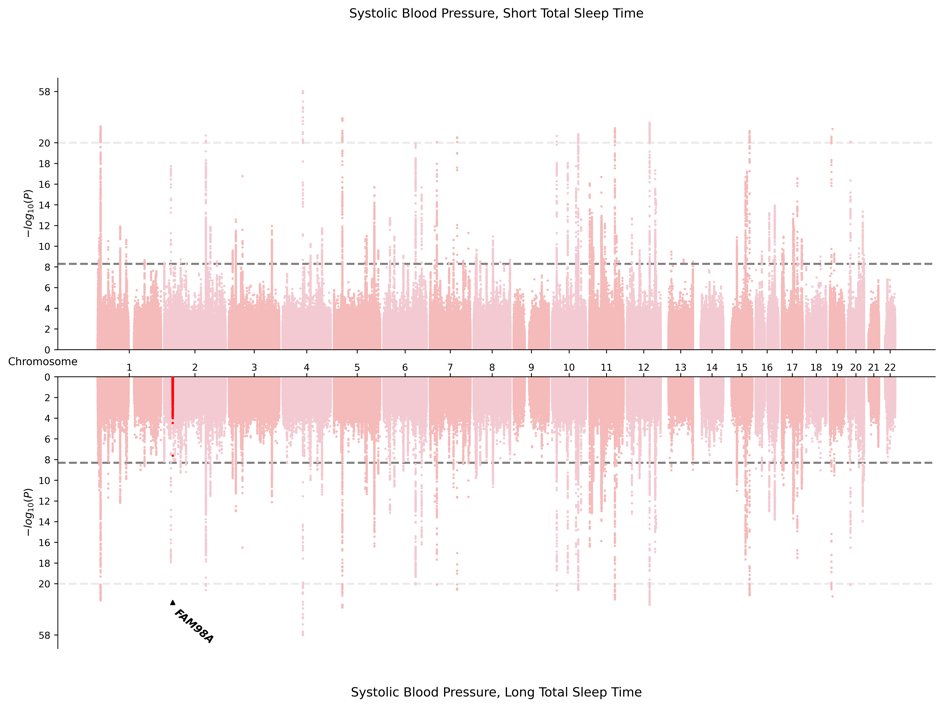

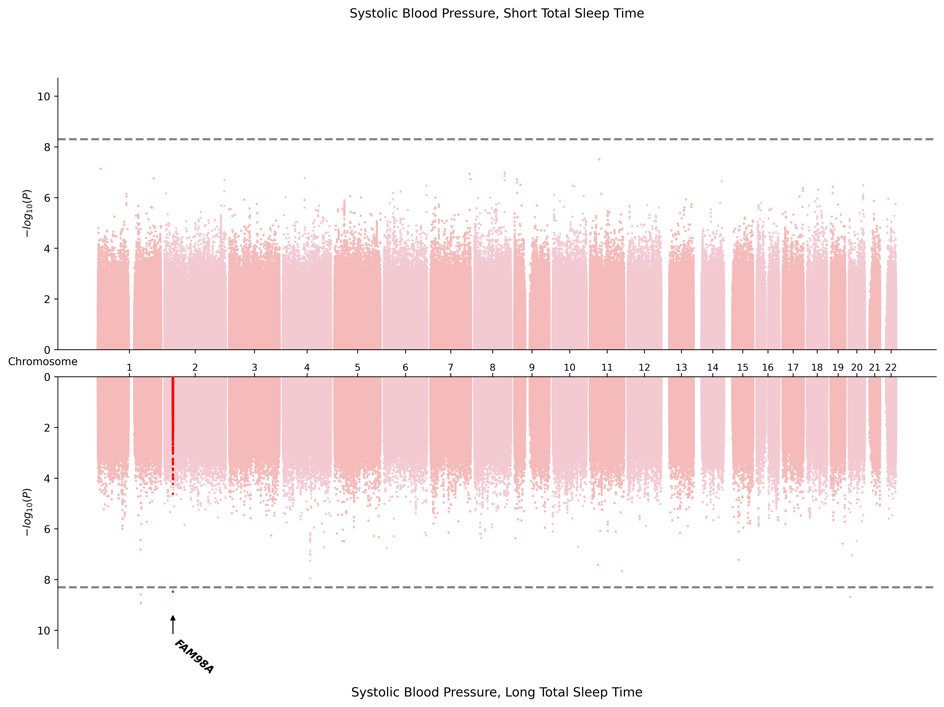

2. AFR-Specific Meta-Analysis

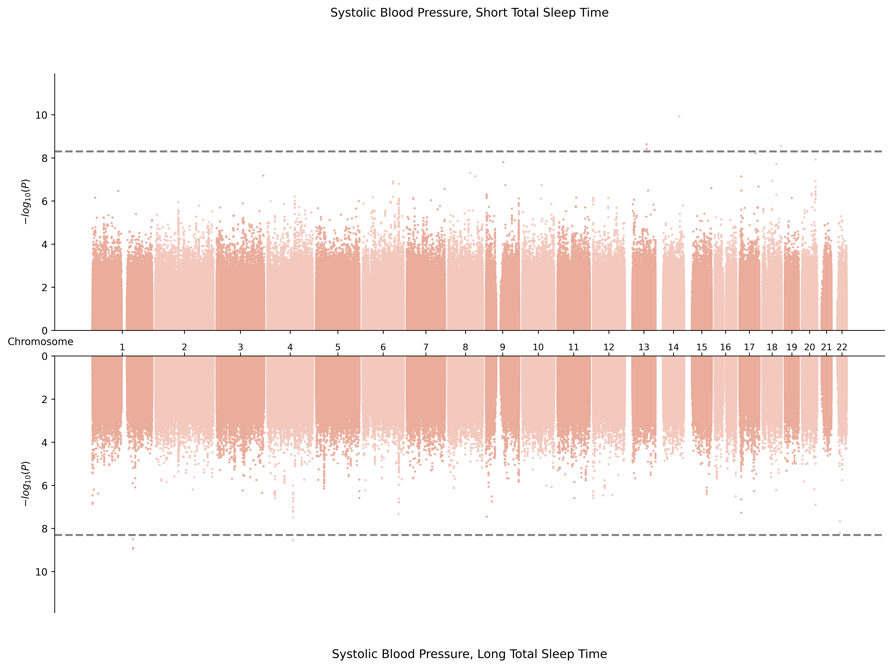

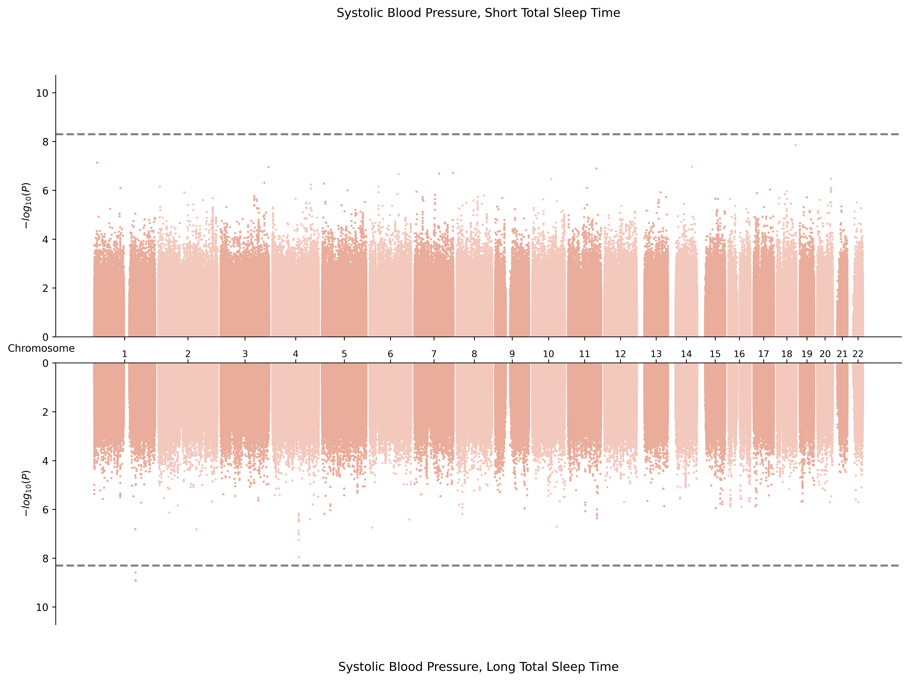

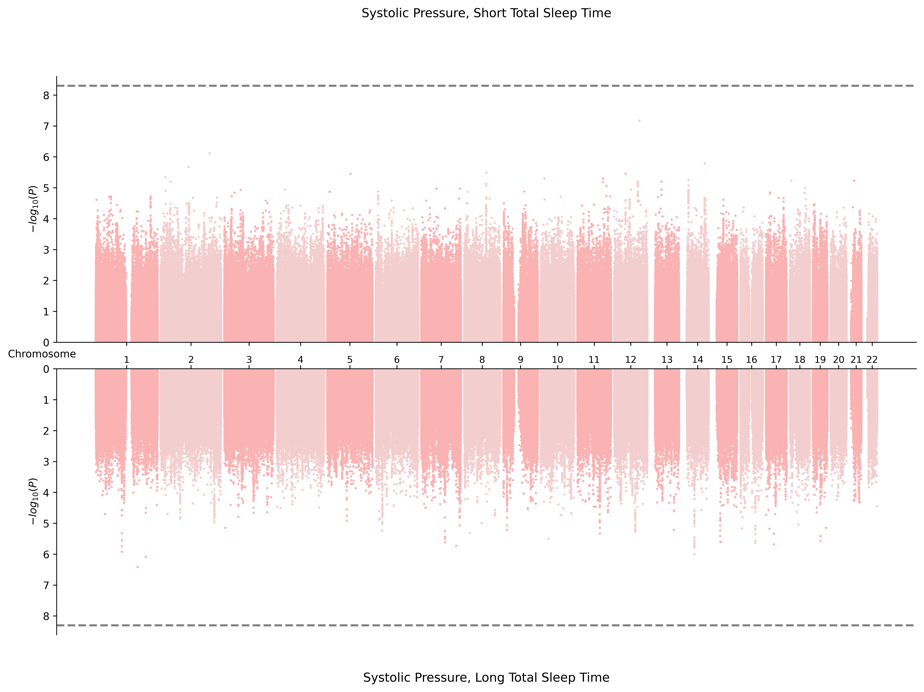
3. EAS-Specific Meta-Analysis

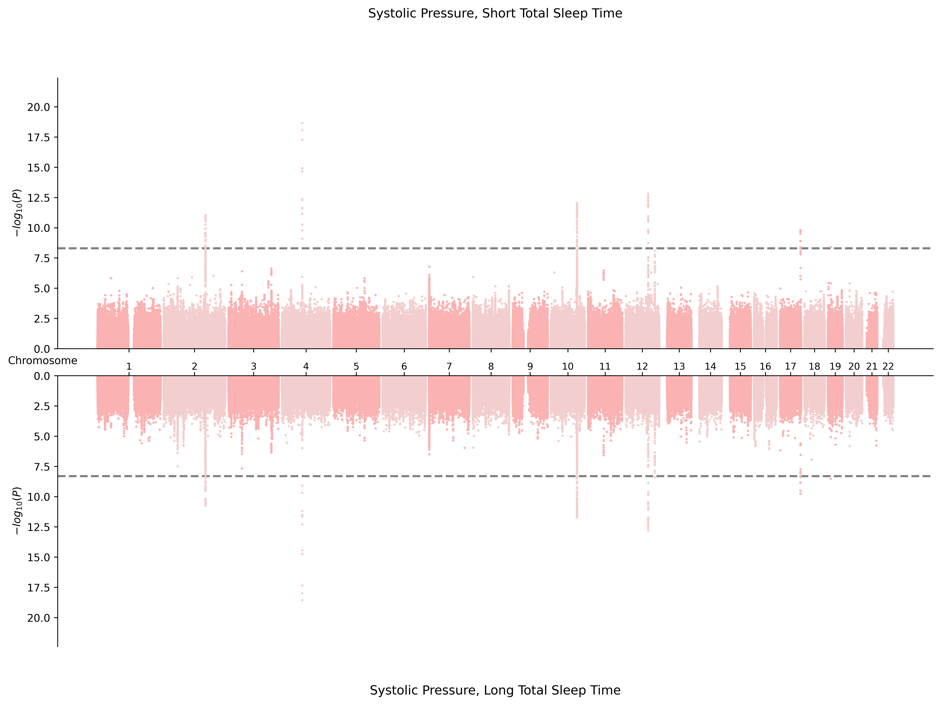

4. EUR-Specific Meta-Analysis

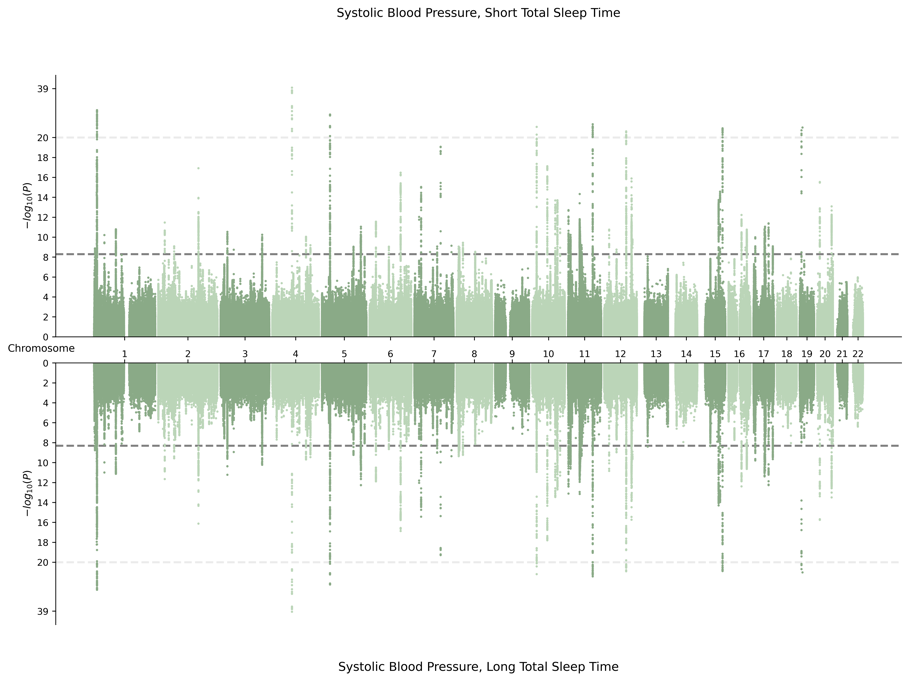

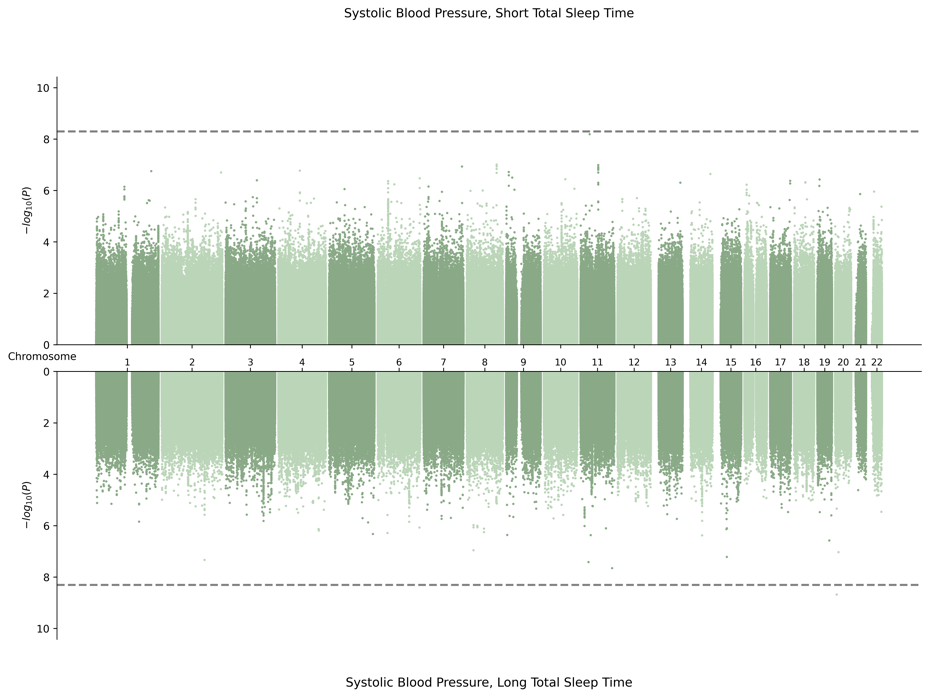

5. HIS-Specific Meta-Analysis

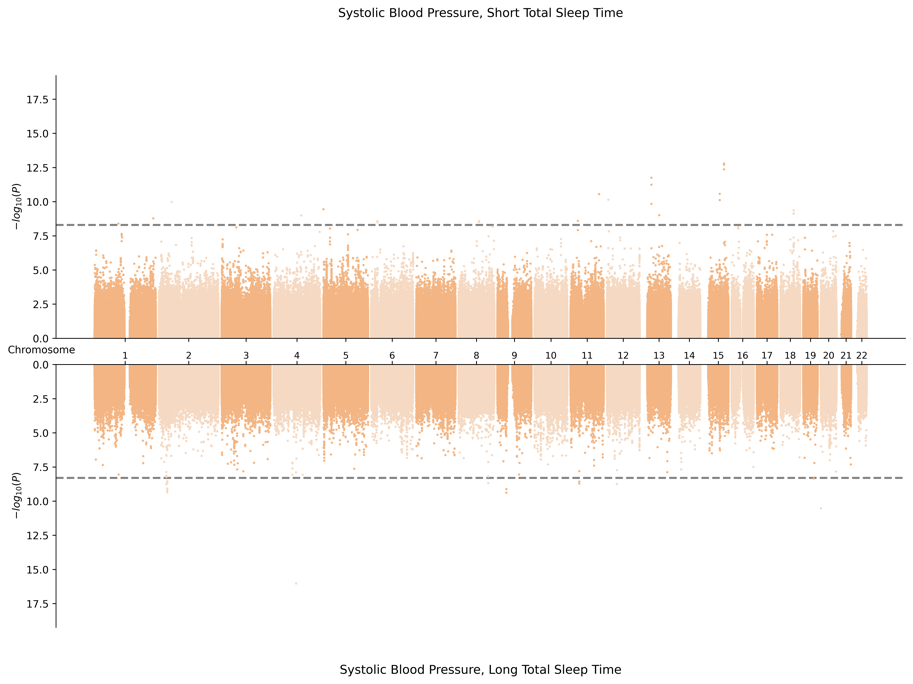

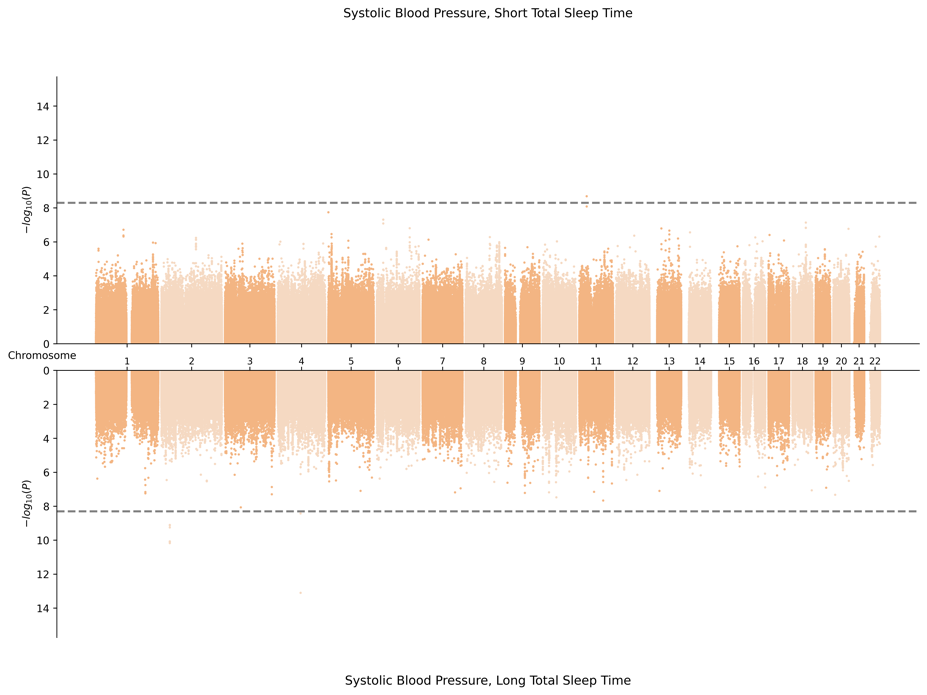

6. SAS-Specific Meta-Analysis

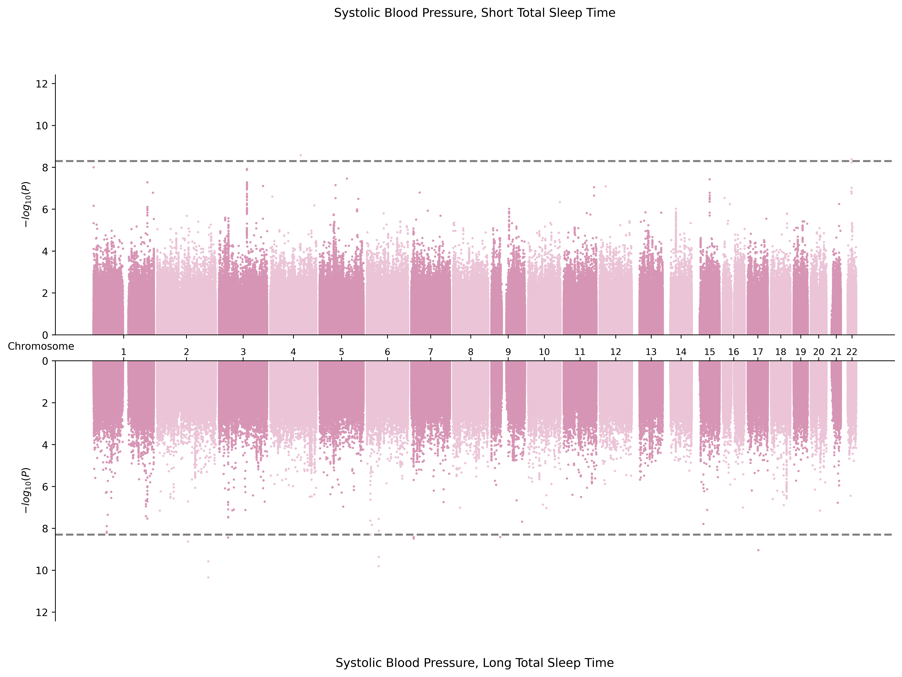

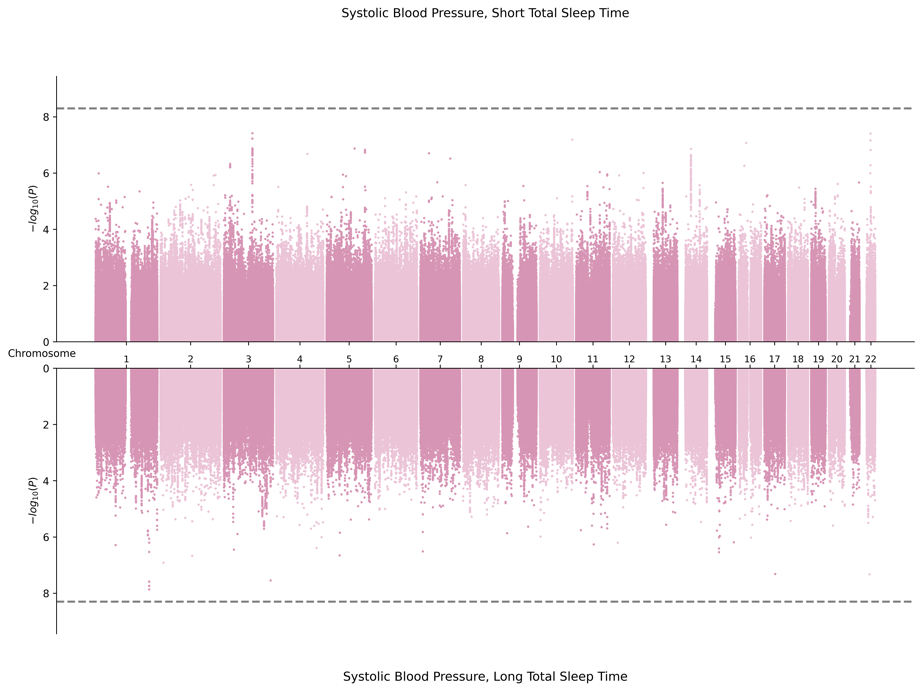

**F.** **Miami Plots for Diastolic Blood Pressure in Female Sex**

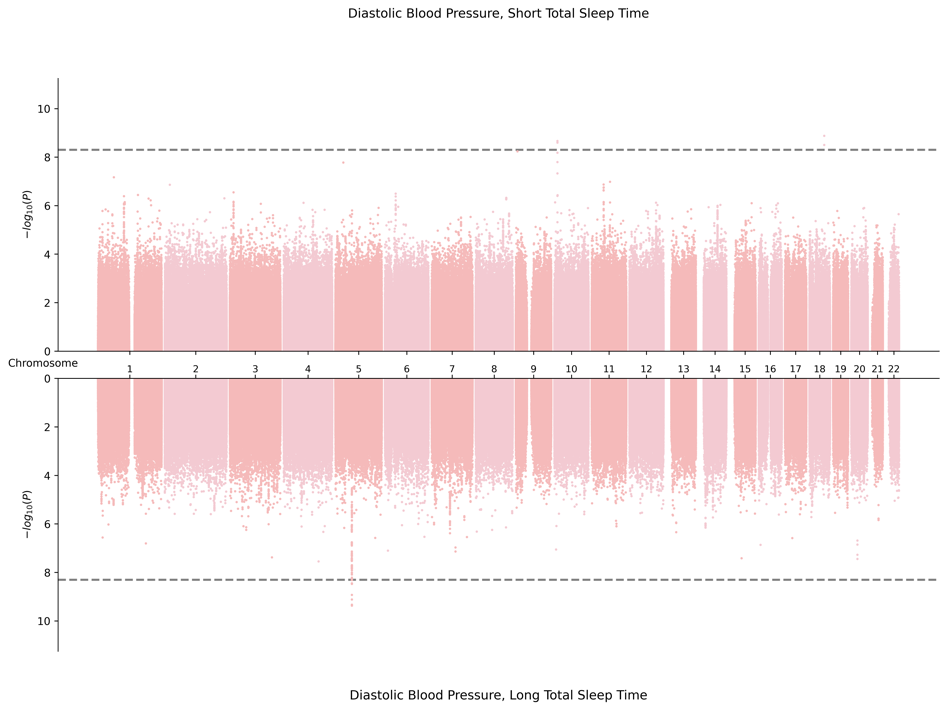
1. Cross Population Meta-Analysis

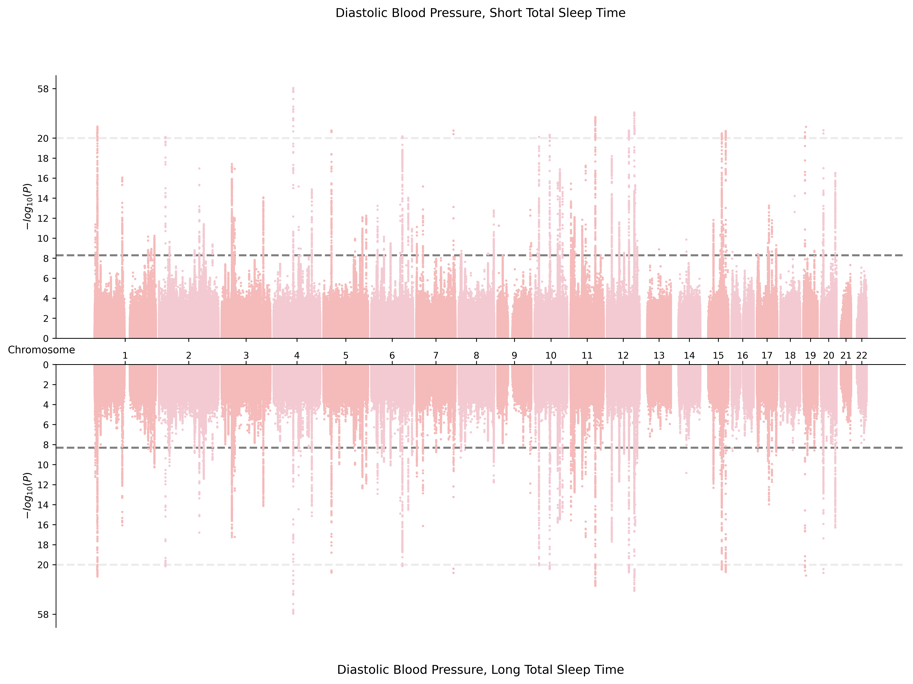

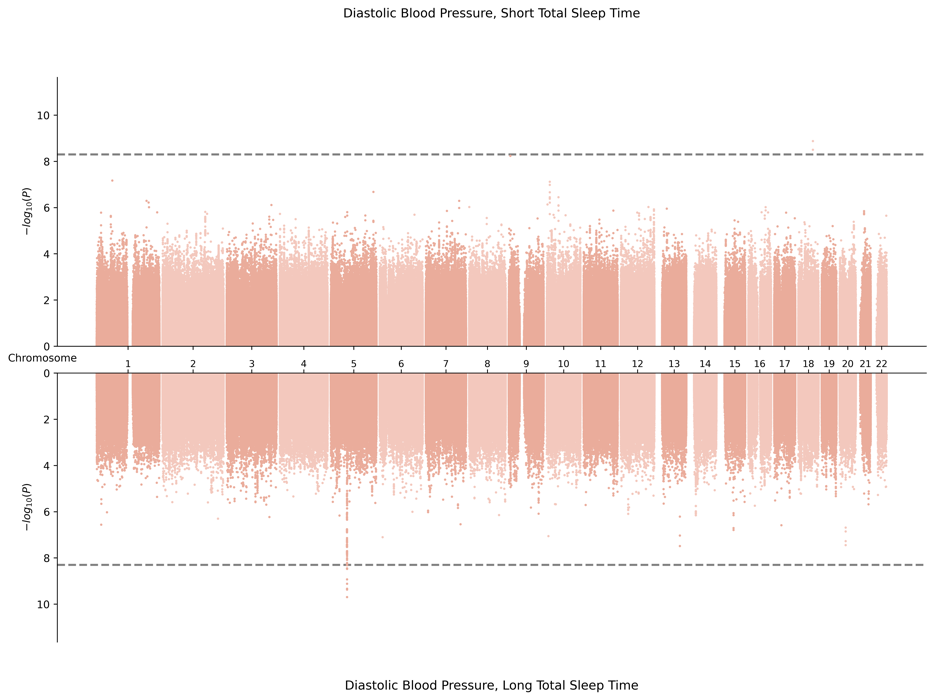
2. AFR-Specific Meta-Analysis

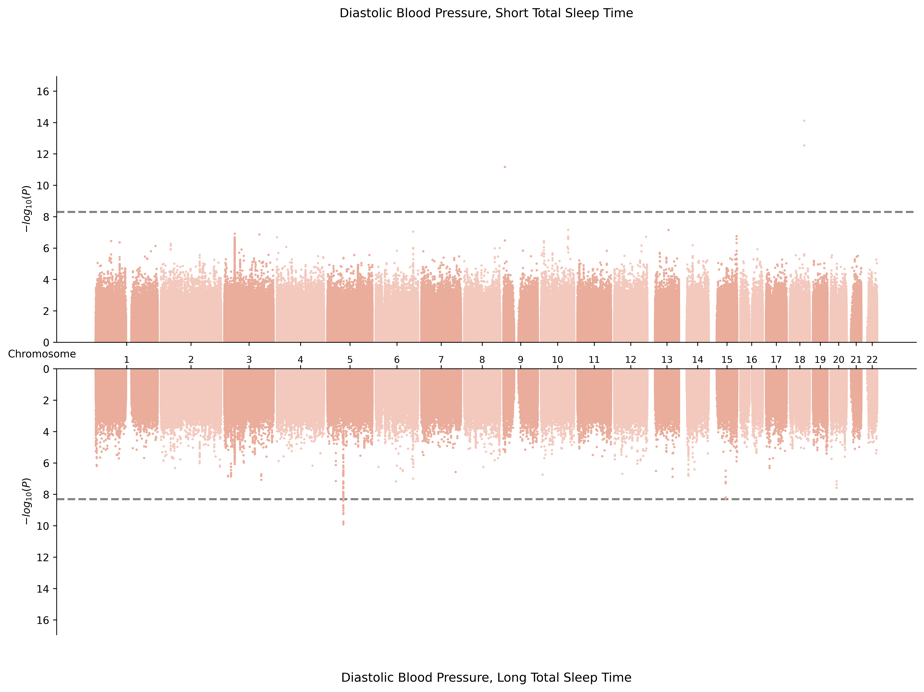

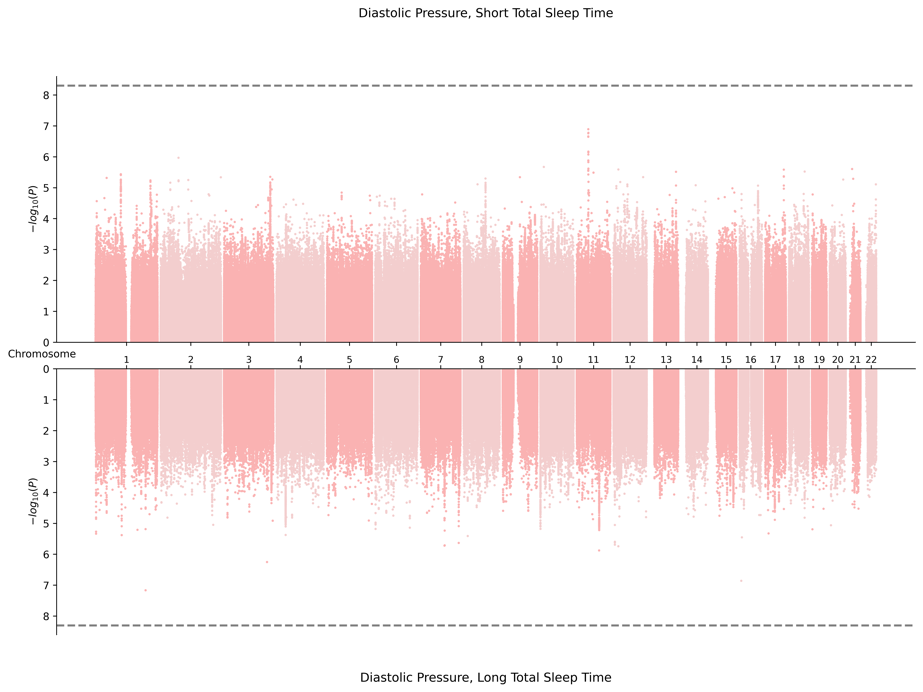
3. EAS-Specific Meta-Analysis

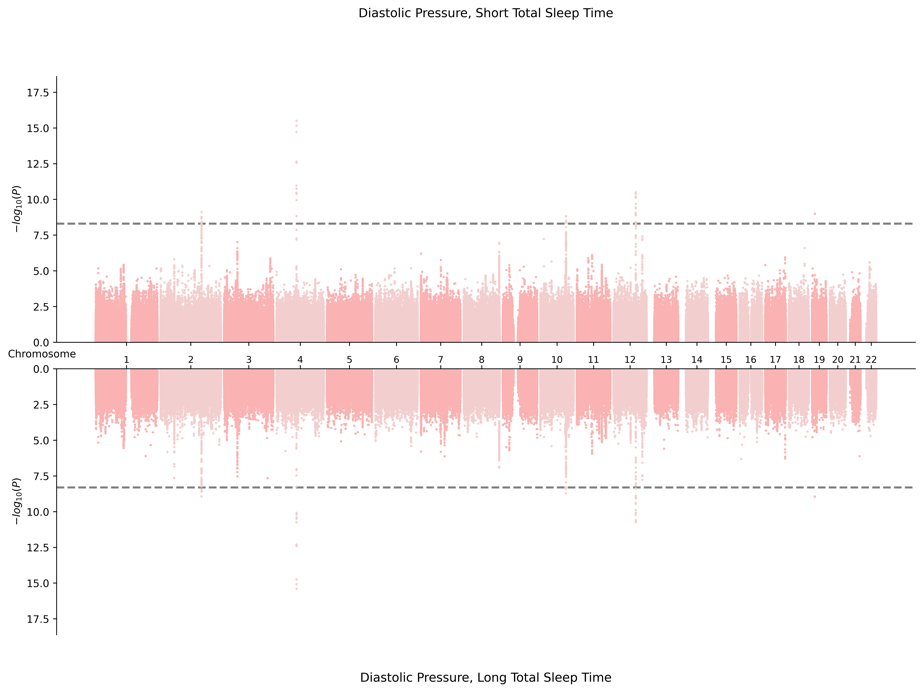

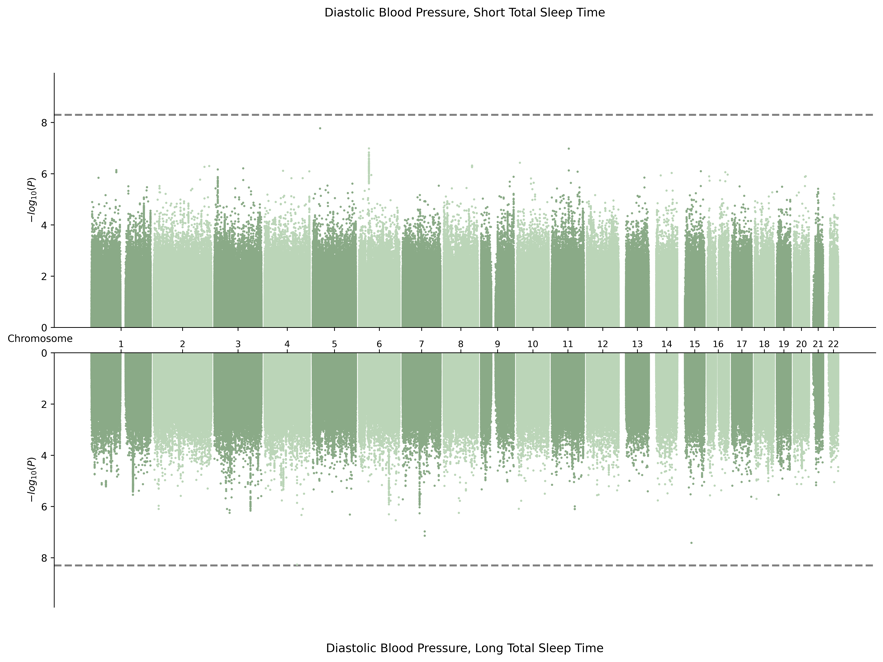
4. EUR-Specific Meta-Analysis

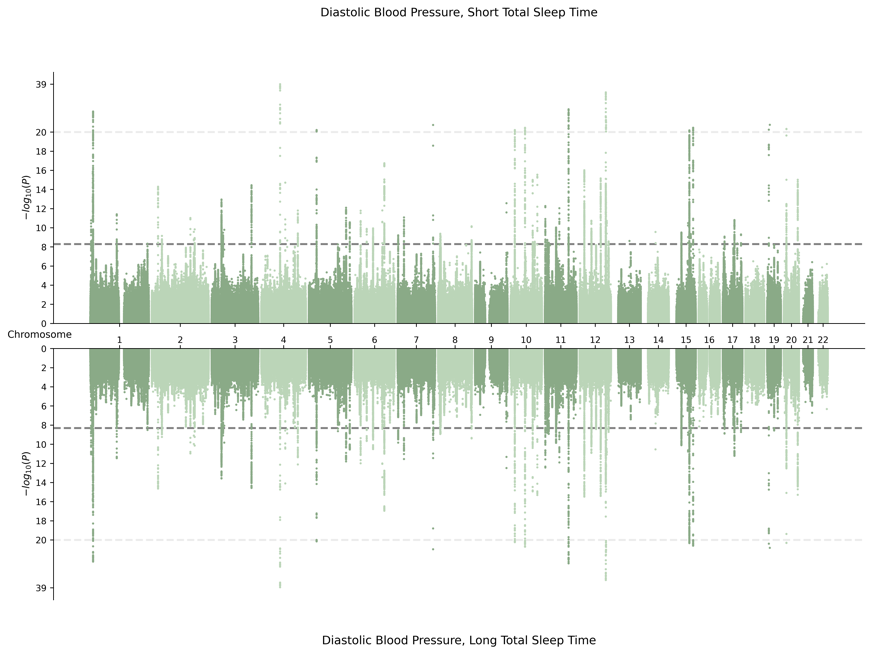

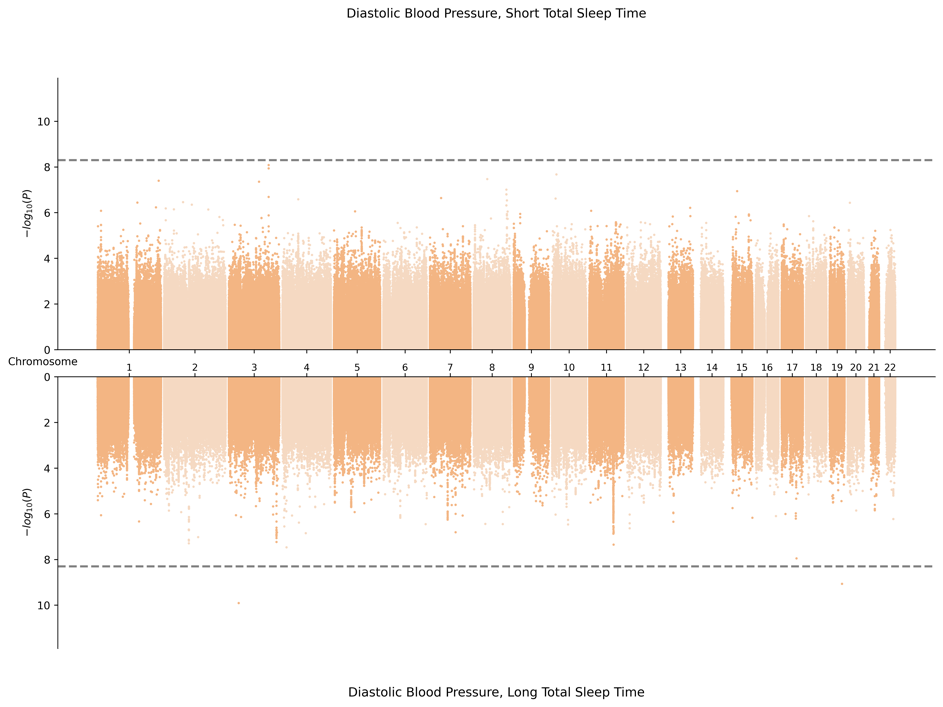
5. HIS-Specific Meta-Analysis

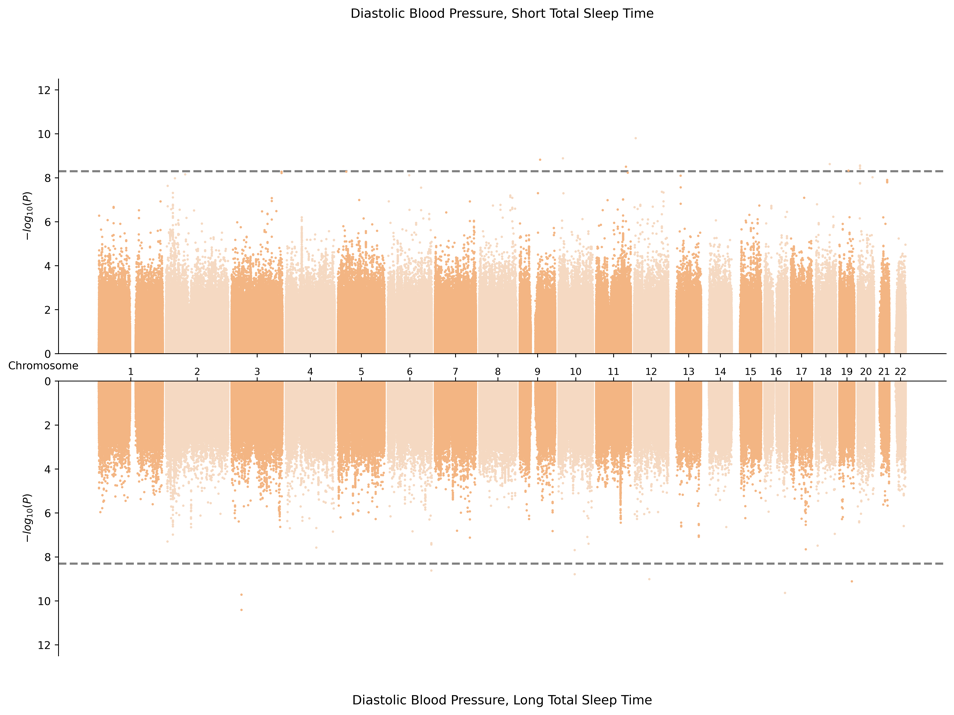

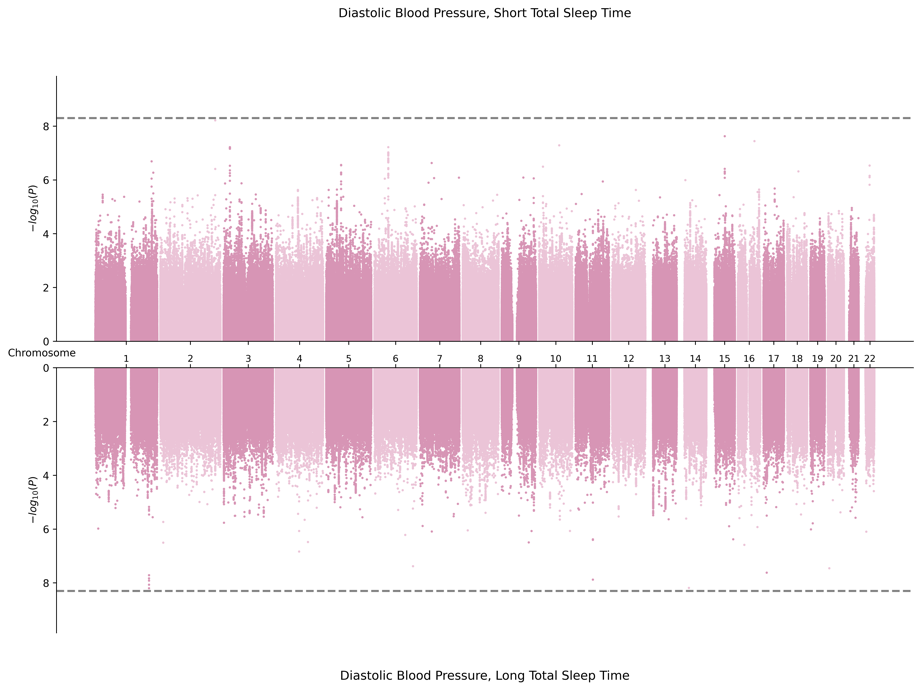
6. SAS-Specific Meta-Analysis

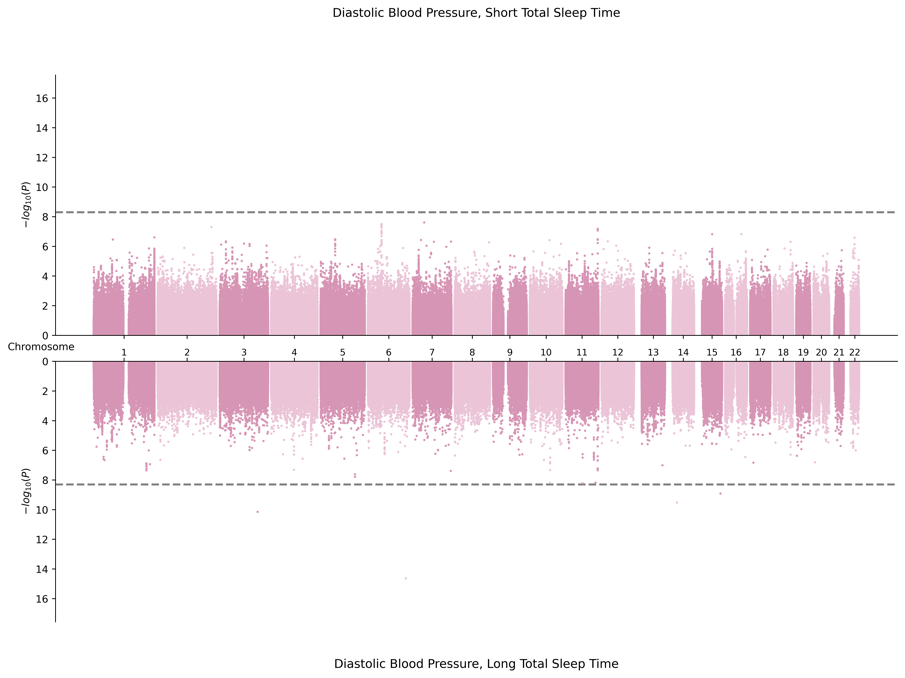

**G.** **Miami Plots for Pulse Pressure in Male Sex**

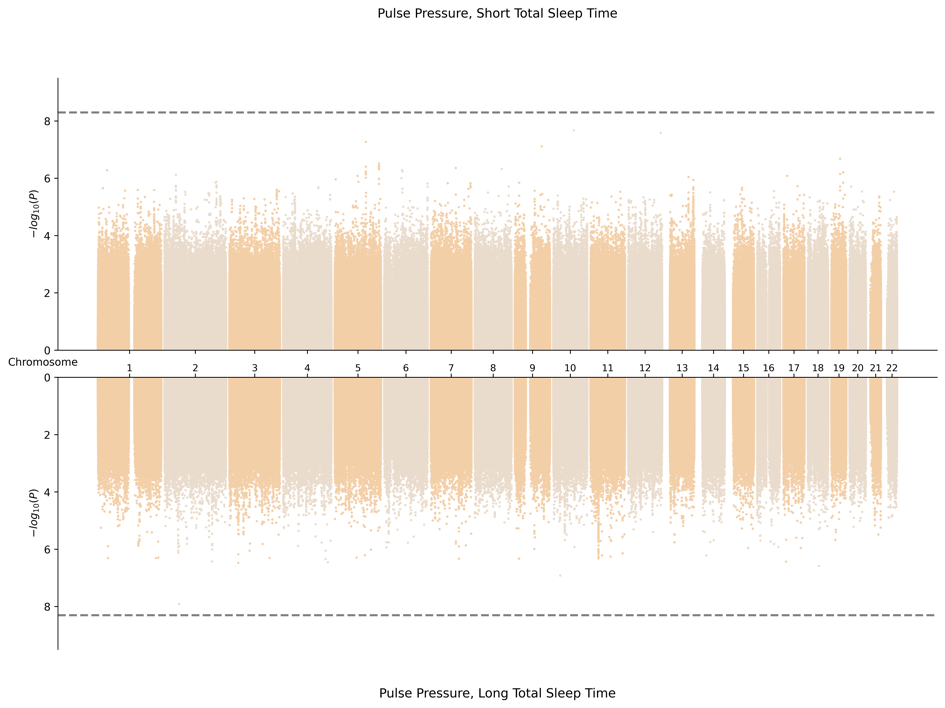
1. Cross Population Meta-Analysis

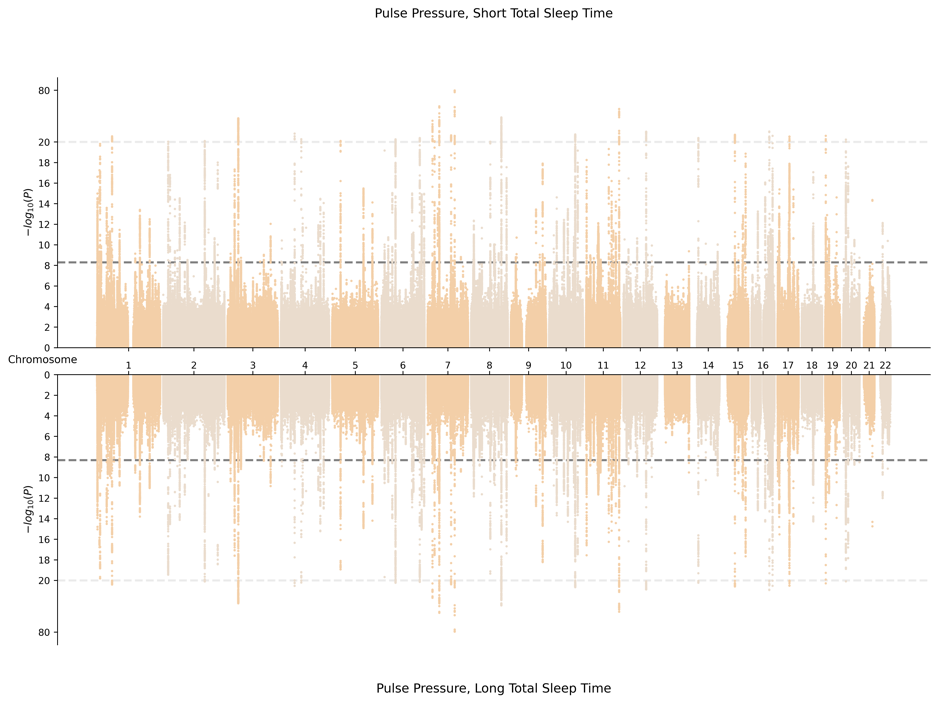

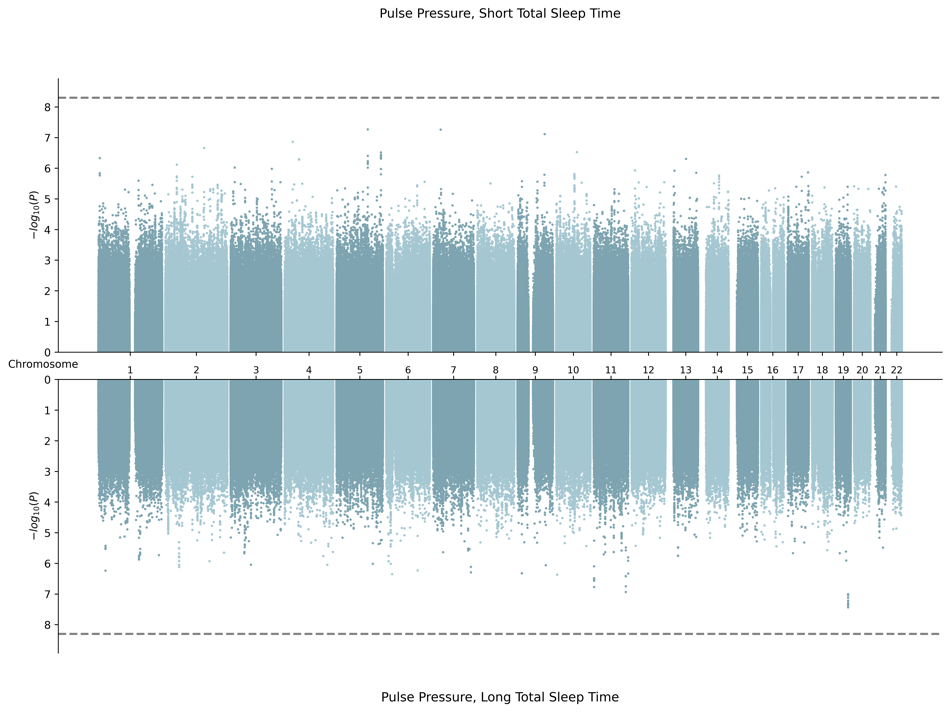
2. AFR-Specific Meta-Analysis

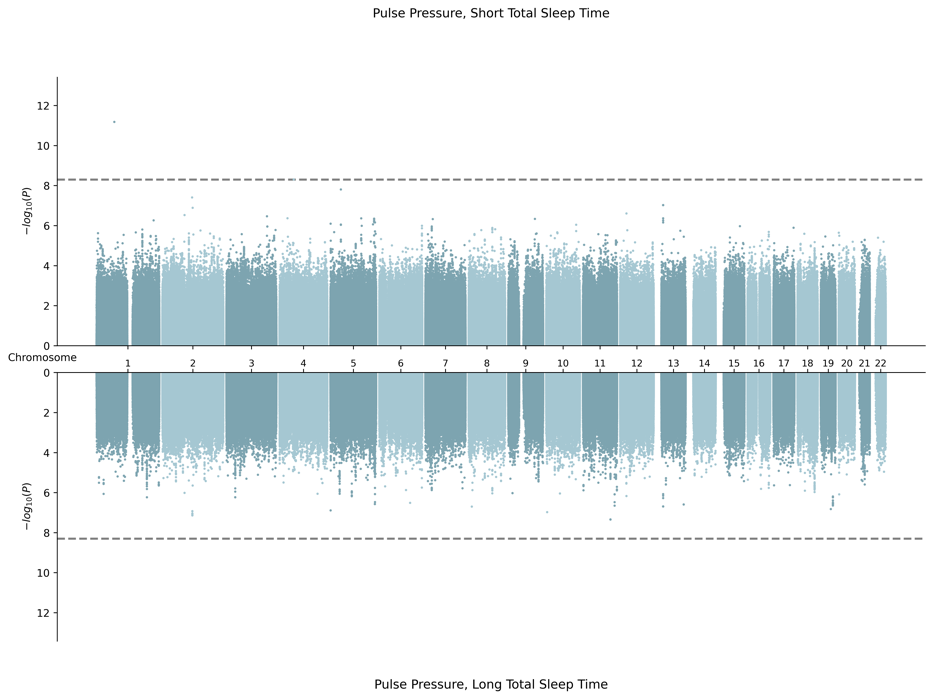

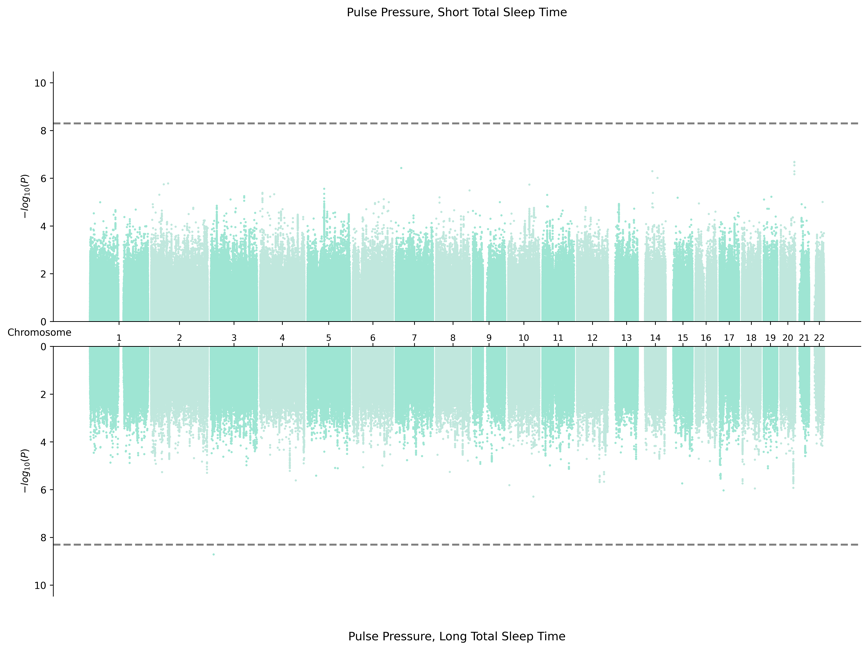
3. EAS-Specific Meta-Analysis

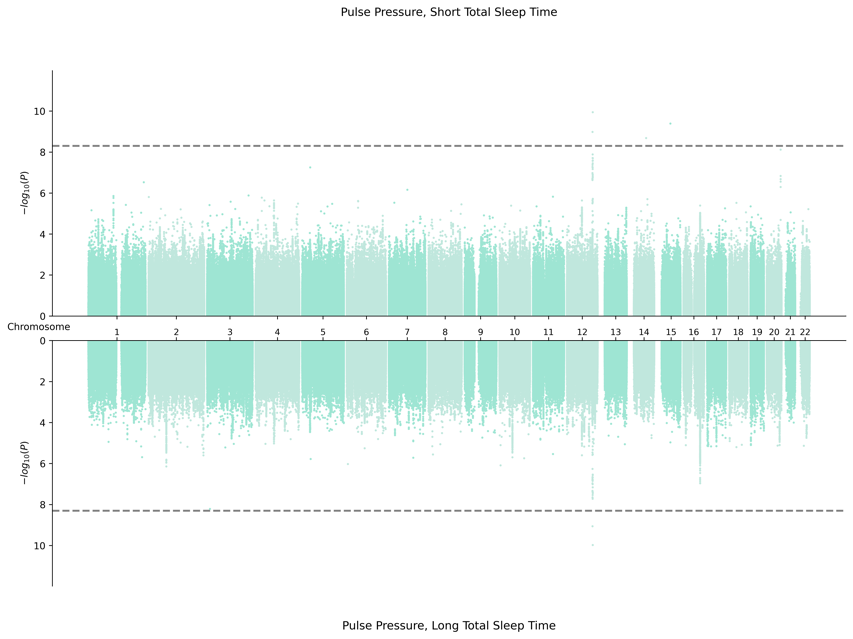

4. EUR Meta-Analysis

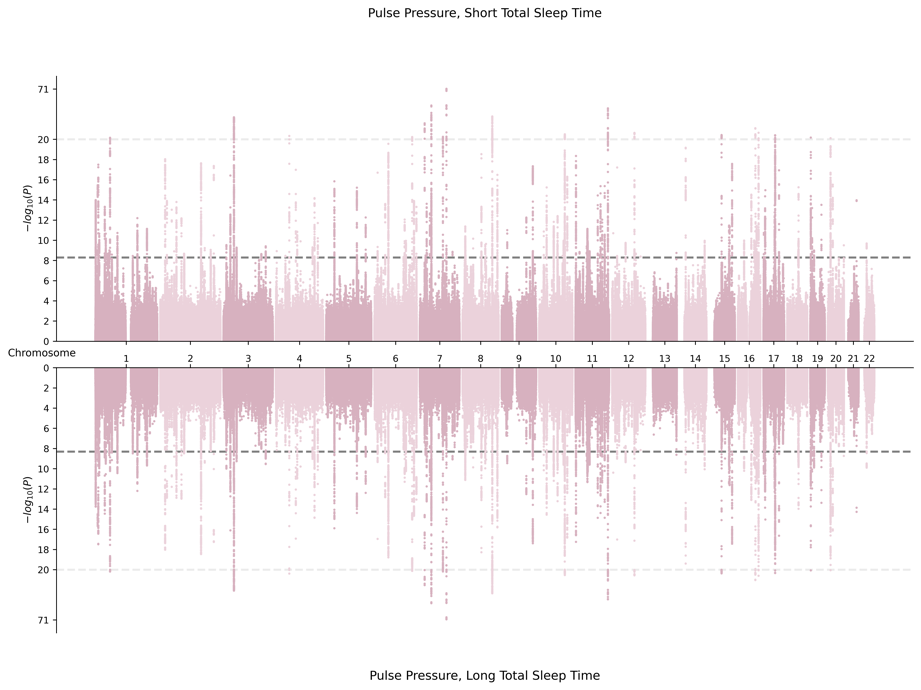

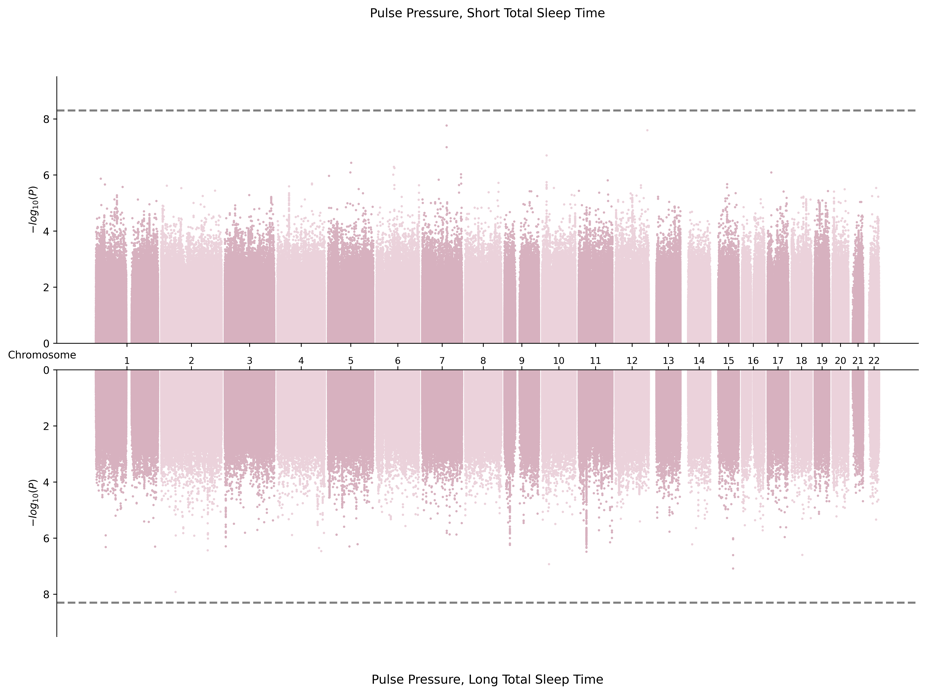

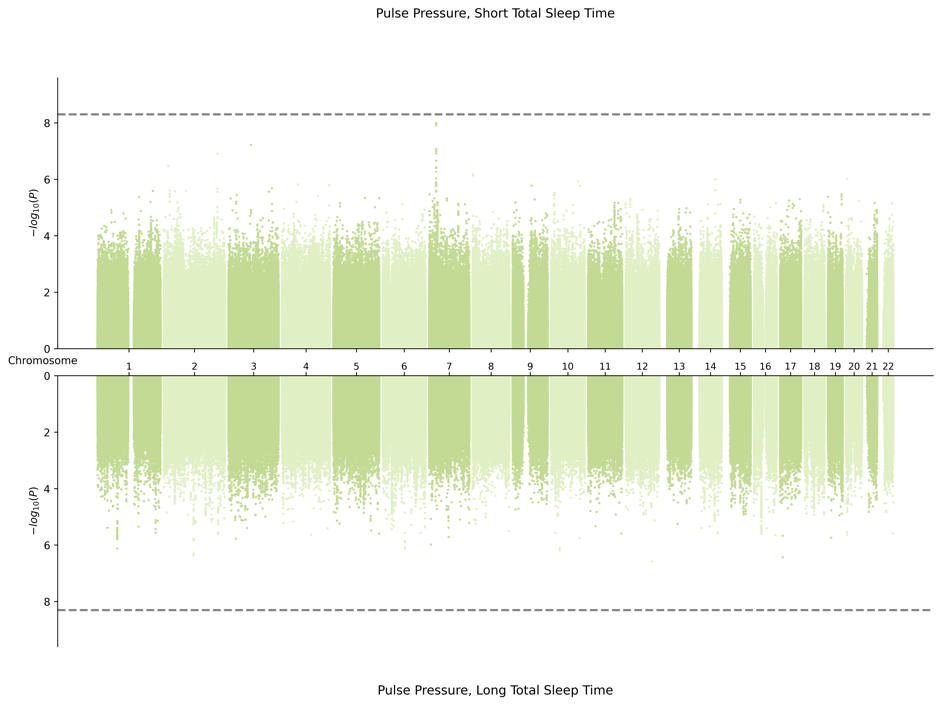
5. HIS-Specific Meta-Analysis

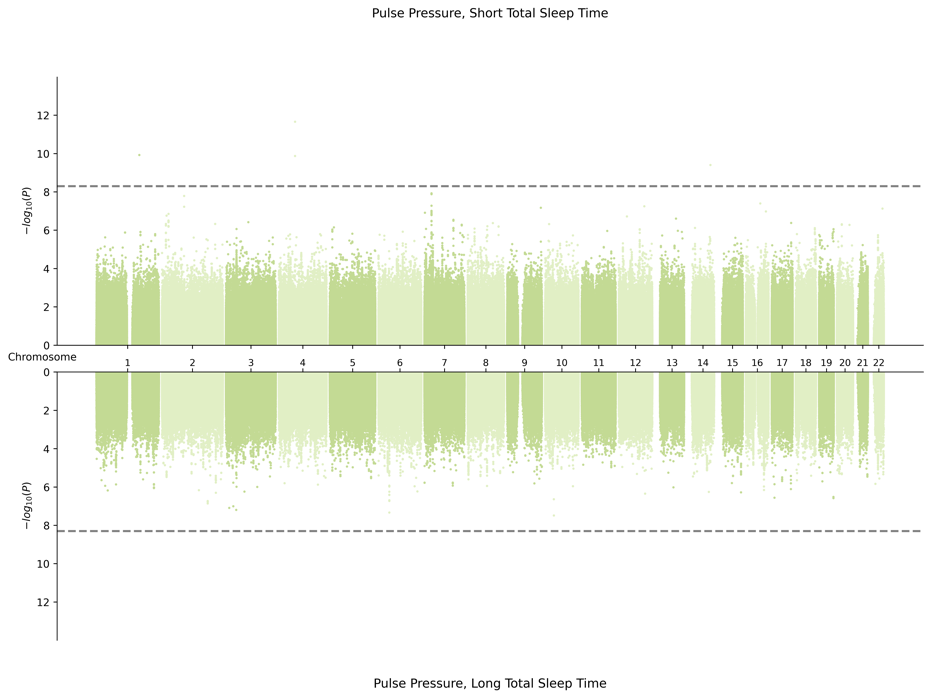

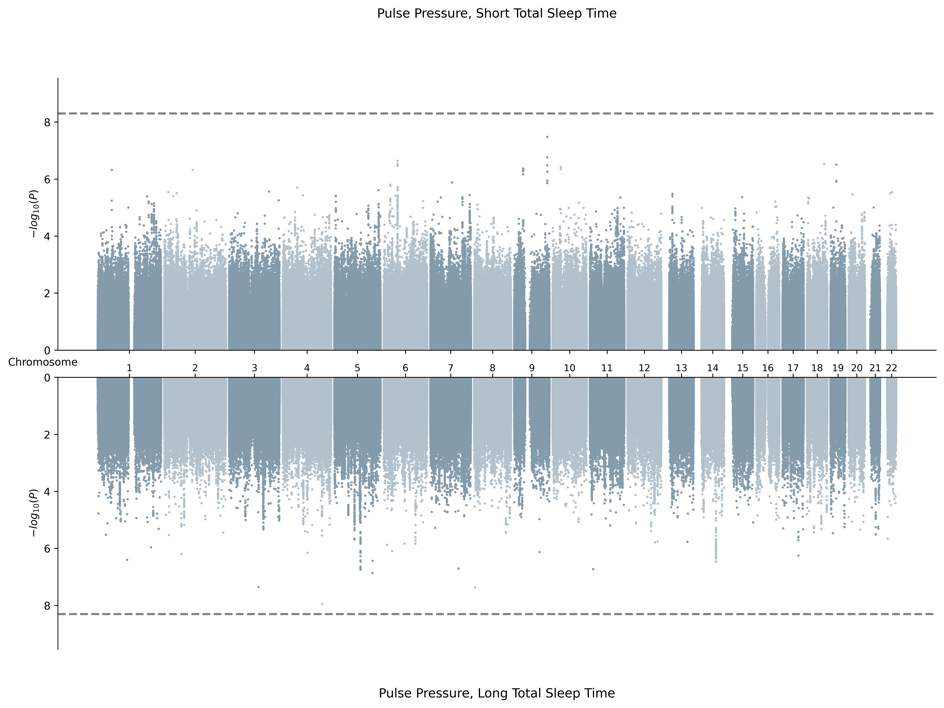
6. SAS-Specific Meta-Analysis

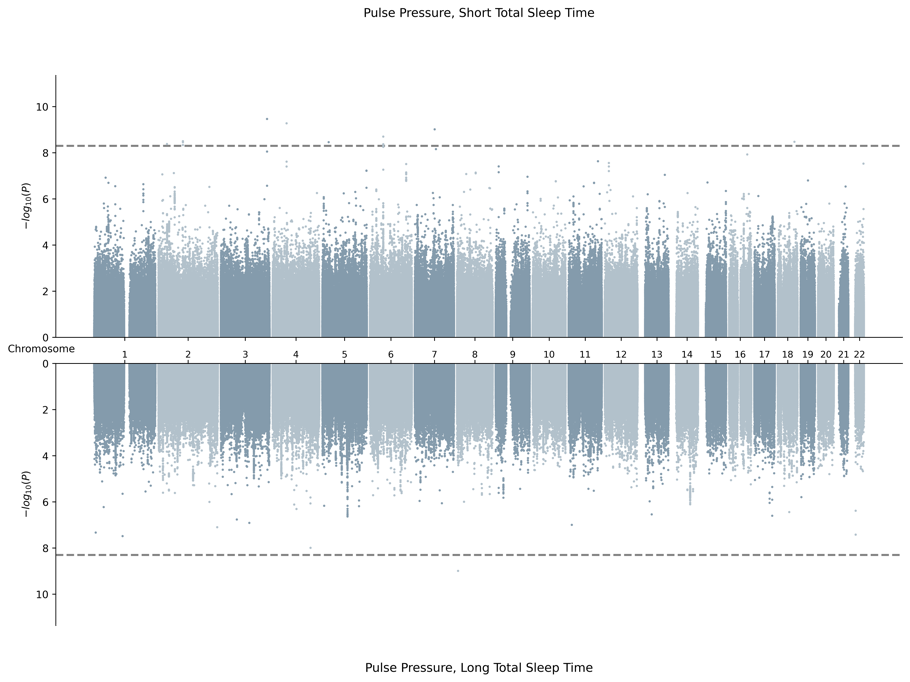

**H.** **Miami Plots for Systolic Blood Pressure in Male Sex**

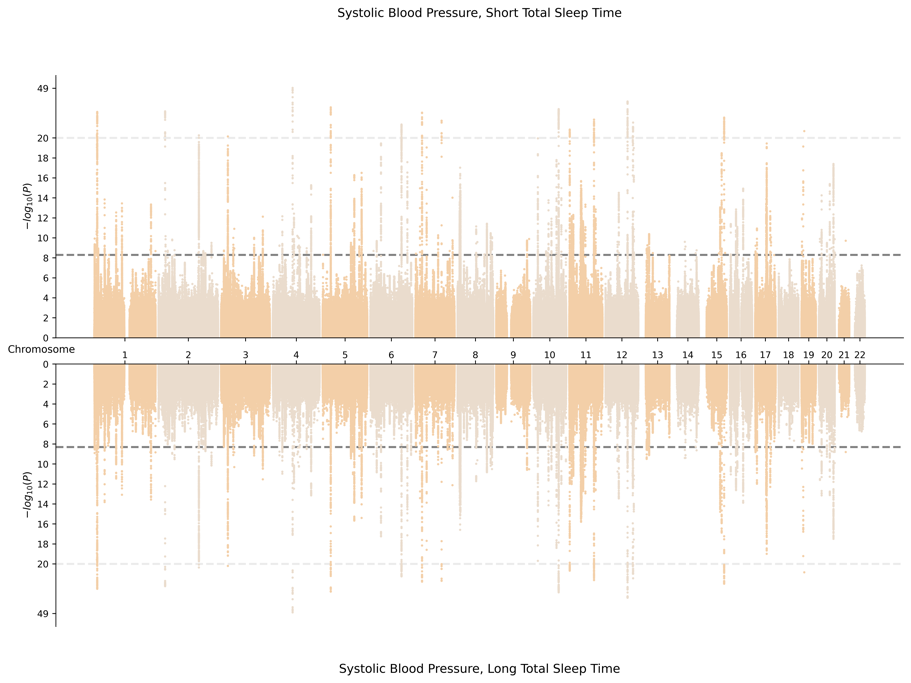

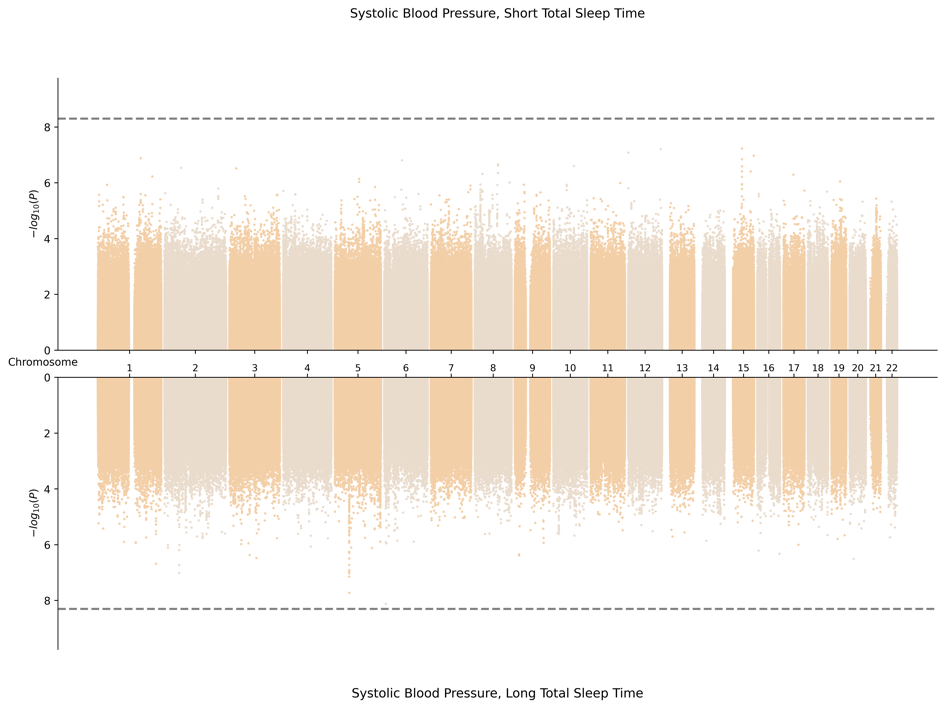
1. Cross Population Meta-Analysis

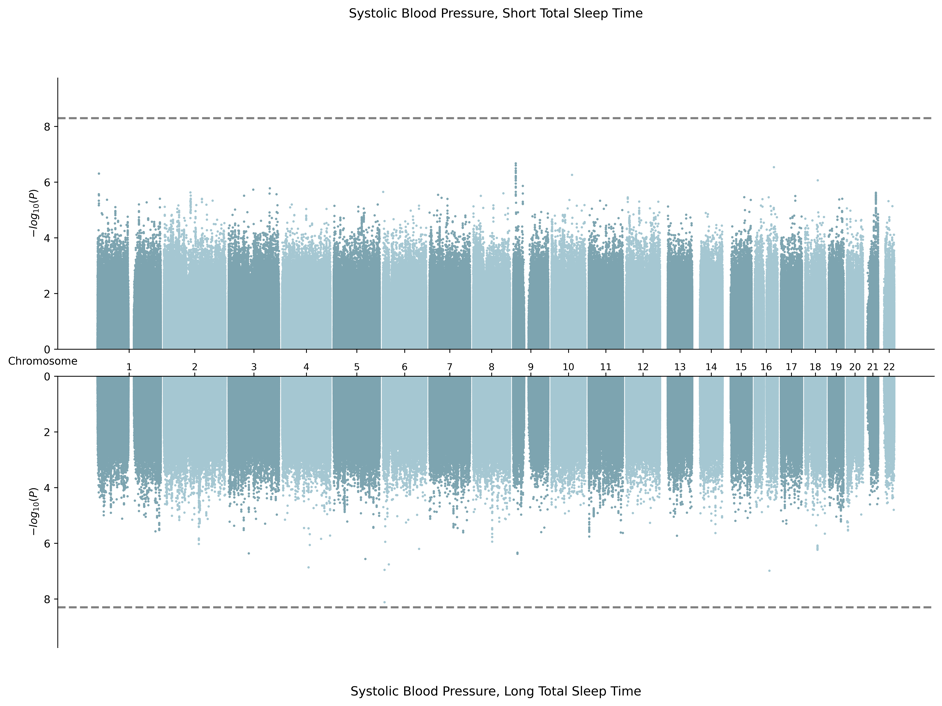
2. AFR-Specific Meta-Analysis

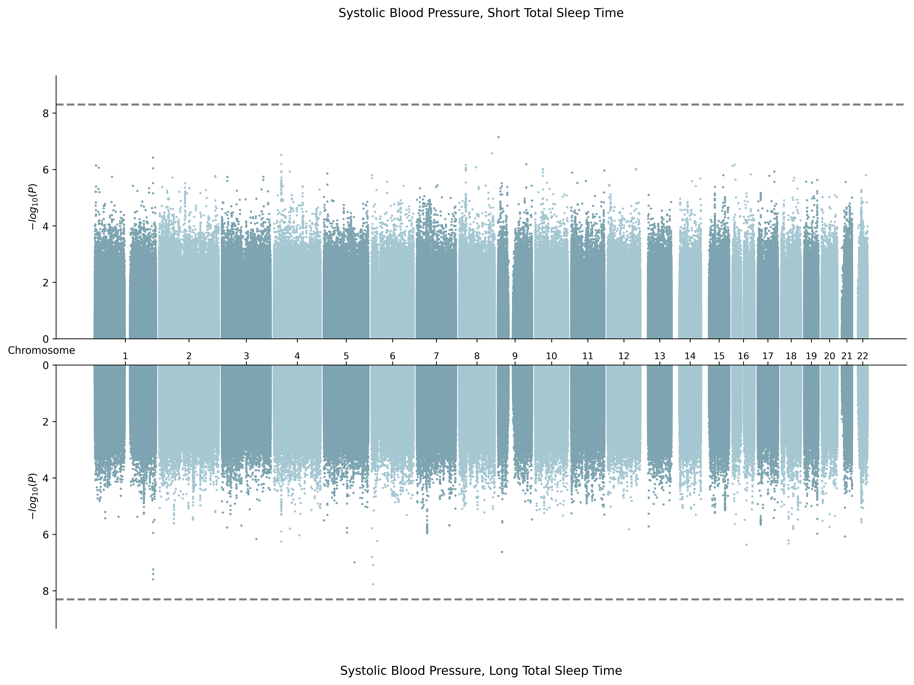

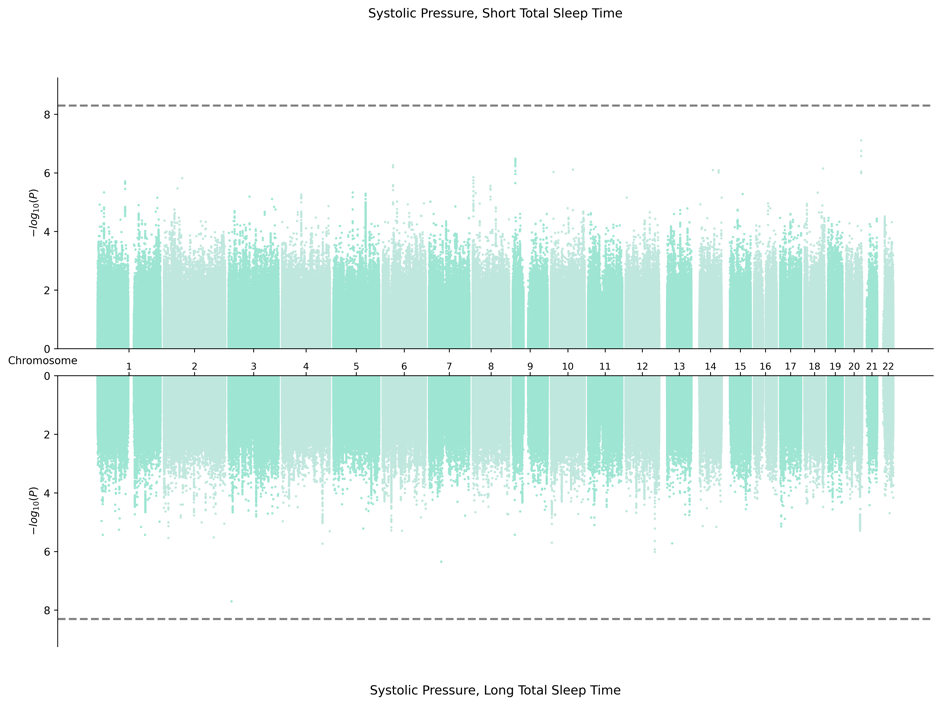
3. EAS-Specific Meta-Analysis

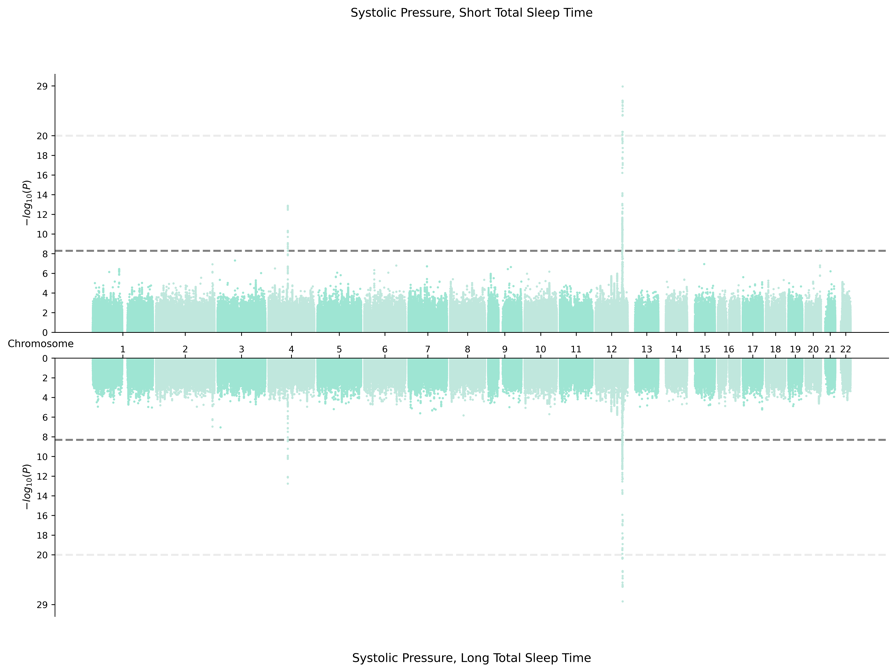

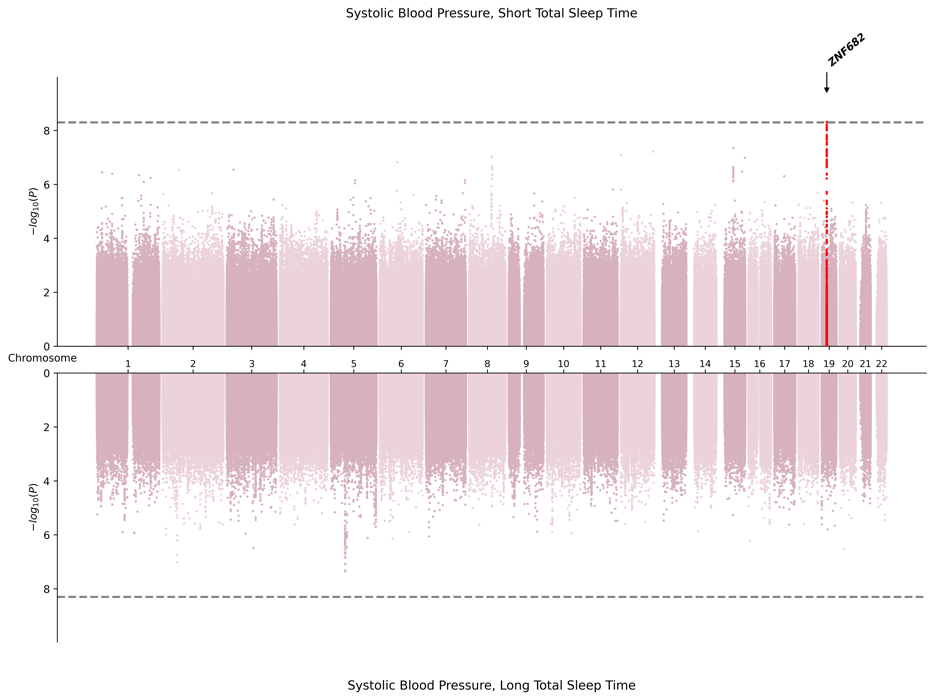
4. EUR-Specific Meta-Analysis

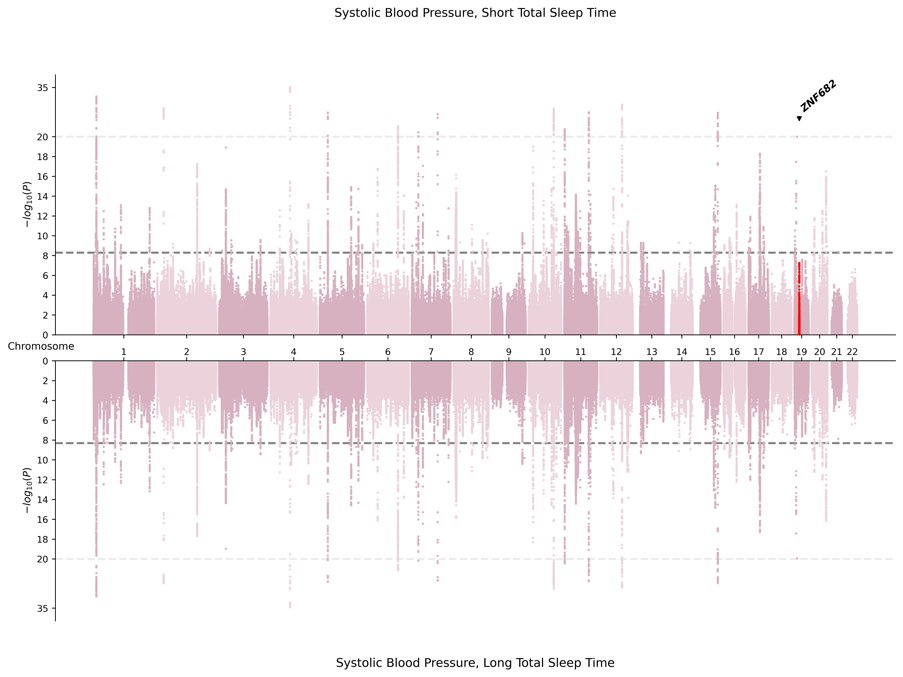

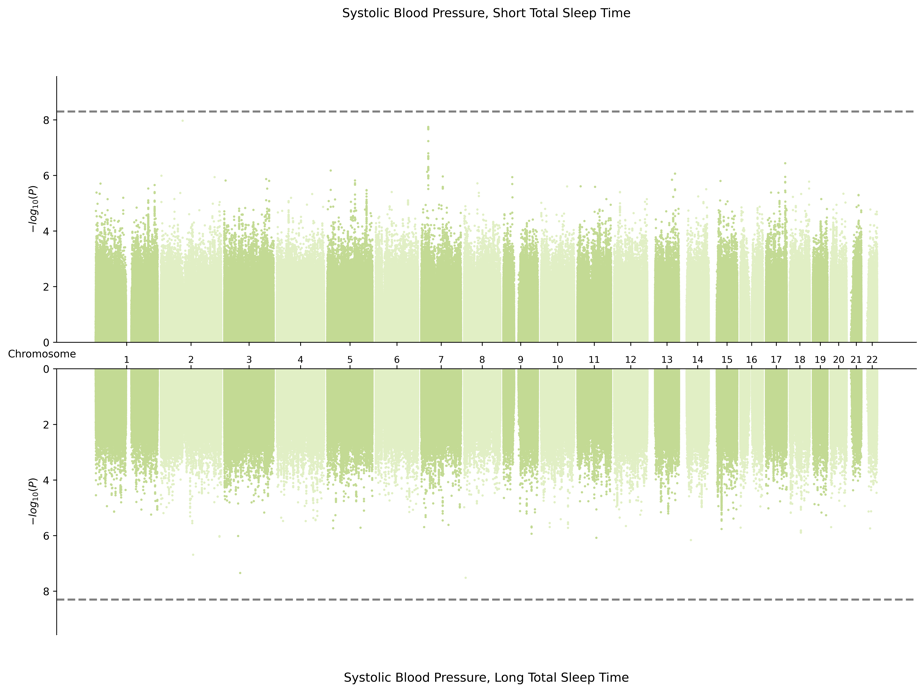
5. HIS-Specific Meta-Analysis

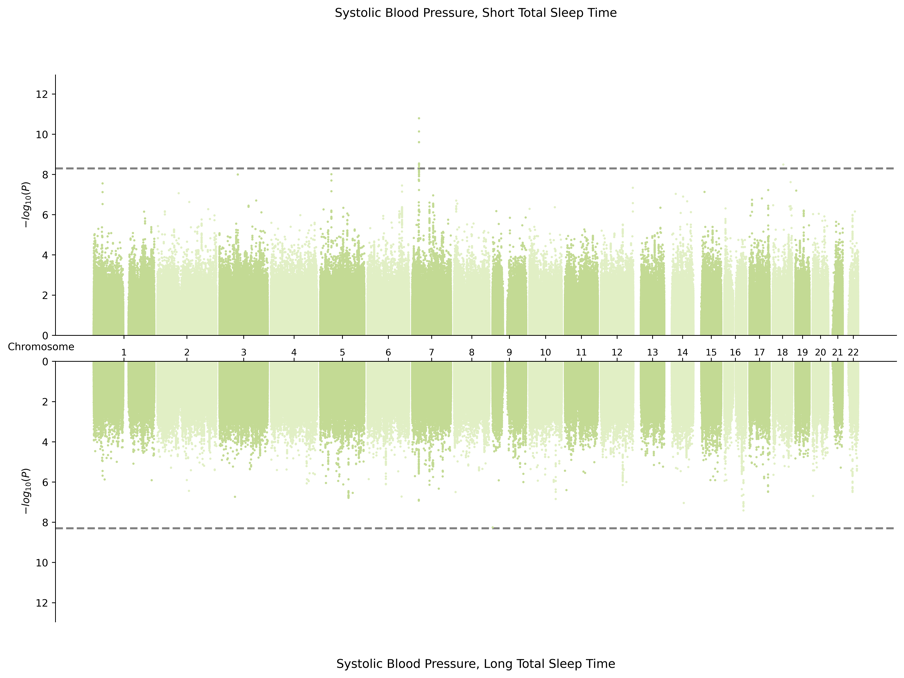

6. SAS-Specific Meta-Analysis

­­

**I.** **Miami Plots for Diastolic Blood Pressure in Male Sex**

1. Cross Population Meta-Analysis

2. AFR-Specific Meta-Analysis

3. EAS-Specific Meta-Analysis

4. EUR-Specific Meta-Analysis

5. HIS-Specific Meta-Analysis

6. SAS-Specific Meta-Analysis Results

**Supplementary Fig. S3**. **Forest Plots**

**A. Cross-Population Variants**

1. rs76458410 *(YWHAB*) was identified in combined sex meta-analysis by the 2df joint test for pulse pressure, found to be driven by interaction with STST.

2. rs1431999695 *(ALG10B)* was identified in female-specific meta-analysis by the 2df joint test for pulse pressure found to be driven by interaction with LTST.

3. rs34761985 (*ST6GAL2*) was identified in combined sex meta-analysis by the 2df joint test for systolic blood pressure, found to not be driven by interaction with STST.

4. rs34761985 (*ST6GAL2*) was identified in combined sex meta-analysis by the 2df joint test for systolic blood pressure, found to not be driven by interaction with LTST.

5. rs13032423 *(VRK2)* was identified in combined sex meta-analysis by the 2df joint test for systolic blood pressure, found to not be driven by interaction with LTST.

**B. AFR-Specific Variant**

1. rs533724062 *(BRINP3)* was identified in combined sex meta-analysis for pulse pressure, found to be driven by the interaction with LTST.

**C. EUR-Specific Variants**

1. rs752086677 (*KRTAP13-2*) was identified in combined sex meta-analysis for diastolic blood pressure by the 1df interaction test for interaction with STST.

2. rs772862932 *(ATP8A2)* was identified in combined sex meta-analysis for diastolic blood pressure by the 2df joint test, found to be driven by interaction with LTST.

3. rs764985249 *(EFNA5)* was identified in combined sex meta-analysis for pulse pressure by the 2df joint test, found to be driven by interaction with STST.

4. rs142966182 *(ALCAM)*, was identified in combined sex meta-analysis for diastolic blood pressure by the 2df joint test, found to be driven by interaction with LTST.

5. rs540041583 *(PAM*) was identified in combined sex meta-analysis for pulse pressure by the 2df joint test driven by interaction with LTST.

6. rs1035064 *(ZNF682*) was identified in male-specific meta-analysis for systolic blood pressure by the 1df interaction test for interaction with STST.

**D. HIS-Specific Variants**

1. rs14383772 *(MROH7)*, was identified in combined sex meta-analysis for systolic blood pressure by the 2df joint test driven by interaction with STST.

2. rs141117715 *(KCNJ3)*, was identified in combined sex meta-analysis for pulse pressure by the 2df joint test driven by interaction with STST.

3. rs17011282 *(ZNF385D),* was identified in combined sex meta-analysis for pulse pressure by the 2df joint test driven by interaction with STST.

4. rs542745170 *(WWOX)* was identified in combined sex meta-analysis for pulse pressure by the 2df joint test driven by the interaction with STST.

5. rs138288695 *(CRBN)* was identified in combined sex meta-analysis for diastolic blood pressure by the 2df joint test driven by interaction with LTST.

6. rs113952142 *(SDK1)* was identified in combined sex meta-analysis for systolic blood pressure by the 2df joint test driven by interaction with LTST.

7. rs111392401 *(JMJD1C)* was identified in combined sex meta-analysis for diastolic blood pressure by the 2df joint test driven by interaction with LTST.

8. rs59680540 *(PRMT6)* was identified in combined sex meta-analysis for pulse pressure by the 2df joint test, not driven by interaction with STST.

9. rs150586434 *(HTR1F)* was identified in combined sex meta-analysis for systolic blood pressure by the 2df joint test, not driven by interaction with STST.

**Supplementary Fig. S4**. **Phenome-wide Association Results From Common Metabolic Diseases Knowledge Portal**

**Note:** Below are p-values plotted and derived from (<https://md.hugeamp.org/>) from past GWAS studies.

**Supplementary Fig S5.** **Brain Imaging Association Results (p<5e-08) From Oxford Brain Imaging Genetics Server (BIG40)**

A. Phenome-wide Association Results for *rs13032423 (VRK2)*
B. Genome-wide Association Results for *rfMRI connectivity ICA100 edge 965*
